# Integrated analysis of high-throughput sequencing-based lncRNA-mediated ceRNA network in Hepatic Alveolar Echinococcosis

**DOI:** 10.1101/2024.07.02.24309861

**Authors:** Zhen Liu, Chang-zhen Shang, Jin-peng Wang, Zhi-gang Gai, Fu-cai Ma, Pan Xia, Yan Wang, Xiao Yang, Hai-hong Zhu

**Affiliations:** Department of Graduate School, Qinghai University, Xining, Qinghai Province, China; Department of General Surgery, Qinghai Provincial People’s Hospital, Xining, Qinghai Province, China; Department of Hepatobiliary Surgery, Sun Yet-Sen Memorial Hospital, Sun Yet-Sen University, Shanwei, Guangdong Province, China

**Keywords:** High-throughput sequencing, Hepatic Alveolar Echinococcosis, long non-coding RNA, competitive endogenous RNA network, lnc-PCBP1-AS1-miR20b-5p/CAPRIN2 axis

## Abstract

**Background:** Numerous studies have indicated that long non-coding RNAs (lncRNAs) can modulate the expression of target gene mRNAs by adsorbing microRNAs (miRNAs). The lncRNA–miRNA–mRNA ceRNA network has been theorized to play an indispensable role in many types of tumors, and has been garnering increasing attention. However, the role of the lncRNA-associated ceRNA regulatory network in Hepatic Alveolar Echinococcosis (HAE) remains unclear and requires further exploration.

**Methods:** In this study, high-throughput sequencing was performed on lesion tissues and adjacent tissues from three patients with Hepatic Alveolar Echinococcosis (HAE) to identify differentially expressed RNAs. We utilized Cytoscape (version 3.10.1) to construct the lncRNA-miRNA-mRNA ceRNA network based on the interactions from the miRcode, miRTarBase, miRDB, and TargetScan databases, and identified hub lncRNAs from within the ceRNA network. Through the use of the “clusterProfiler” package in R, we performed Gene Ontology (GO) and Kyoto Encyclopedia of Genes and Genomes (KEGG) pathway annotations for the DEGs (Differentially Expressed Genes) within the ceRNA network. Concurrently, we utilized these DEGs to construct a protein-protein interaction network (PPI). Finally, an analysis was conducted on the PCBP1-AS1-miR-20b-5p/CAPRIN2 axis within the ceRNA network.

**Results:** In HAE, a total of 979 differentially expressed lncRNAs (DELncRNAs) and 870 differentially expressed mRNAs (DEmRNAs) were identified. An HAE-specific ceRNA network comprising 11 lncRNAs, 21 miRNAs, and 56 mRNAs was established, and analysis of this network led to the construction of a sub-network associated with hub lncRNAs. GO and KEGG pathway analyses indicated that the HAE-specific ceRNA network is related to molecular functions and pathways associated with cancer. Subsequent experiments using qPCR and dual-luciferase assays validated the interactions between PCBP1-AS1 and miR-20b-5p, as well as between miR-20b-5p and CAPRIN2. Analysis of the target gene in relation to clinical characteristics of HAE patients suggested that the PCBP1-AS1-miR-20b-5p/CAPRIN2 axis may influence the development of HAE.

**Conclusion:** In this study, we described the gene regulation within the lncRNA-miRNA-mRNA ceRNA network during the development of Hepatic Alveolar Echinococcosis (HAE), which contributes to a deeper exploration of the molecular mechanisms underlying HAE. Additionally, we discovered that PCBP1-AS1 may regulate the expression of CAPRIN2 by adsorbing miR-20b-5p, affecting the onset and progression of HAE. PCBP1-AS1 could potentially serve as a useful target for the diagnosis and treatment of HAE.

## Introduction

Hepatic alveolar echinococcosis (HAE) is a zoonotic disease caused by the larval of the multilocular echinococcus, accounting for 5-10% of the total incidence of echinococcosis^[1]^. The growth of HAE is slow and the disease often remains asymptomatic for an extended period, approximately 5 to 15 years. Its growth pattern resembles that of a malignant condition, capable of infiltrating or invading surrounding tissues and organs, and it may even metastasize to distant organs^[2]^. Despite the advancements made in the diagnosis and treatment of HAE, the therapeutic outcomes and prognosis are still less than satisfactory^[3]^. Research conducted through BLAST sequence analysis of the Echinococcus genome has indicated that approximately one-third of the genes may be unique characteristics of the species and form the basis of its biological traits, which are specific to Echinococcosis. These genes could potentially aid in refining diagnostic methods and identifying novel therapeutic targets^[4]^. Therefore, this study preliminarily explores the biological mechanisms and pathogenesis of HAE at the genetic level, in an effort to identify new therapeutic targets to improve treatment outcomes and prognosis.

In recent years, advances in sequencing technologies have enabled more in-depth genomic research. It is now known that less than 2% of the human genome encodes proteins. Among the various types of non-coding RNAs (ncRNAs), long non-coding RNAs (lncRNAs) have attracted significant attention. These are defined as long chains of non-coding RNA that exceed 200 base pairs in length and do not have the function of encoding proteins^[5][6]^. However, through their diverse mechanisms, lncRNAs play functional roles in various cell types, and many lncRNAs possess both tumor-suppressive and oncogenic capabilities^[7][8]^. Additionally, microRNAs (miRNAs) are endogenous RNA molecules approximately 22 base pairs in length that can bind to target RNAs, promoting the degradation of mRNA or inhibiting protein translation^[9][10]^, they play a crucial role in regulating gene expression and are involved in numerous biological processes through their network of regulatory interactions^[11][12][13]^.

In 2011, Salmena and colleagues first proposed the competitive endogenous RNA (ceRNA) hypothesis. This hypothesis suggests that ceRNAs interact with target miRNAs through miRNA response elements (MREs), thereby regulating the transcriptome on a large scale, Simultaneously, this concept of ceRNAs and their networks offers a novel perspective on the intricate interactions of RNA molecules within the cell, enhancing our comprehension of gene regulation and the potential for targeted therapies in the treatment of diseases^[14]^. In recent years, a growing body of research has indicated that lncRNAs can act as ceRNAs, regulating the expression of target mRNAs by competitively sharing miRNAs. They play a significant role in various types of tumors, Such as in hepatocellular carcinoma^[15]^, pancreatic cancer^[16]^, gastric cancer^[17]^, and colorectal cancer^[18]^.However, to date, there have been no reports on the study of lncRNAs and ceRNA networks in HAE, and the expression profile of lncRNAs and the associated ceRNA regulatory networks in HAE require further clarification. Therefore, it is particularly important to further understand the role of lncRNAs in the development of HAE and to investigate the pathogenic mechanisms involving ceRNA networks in HAE.

In this study, we performed high-throughput sequencing on lesion tissues and adjacent tissues from three HAE patients to identify differentially expressed lncRNAs and mRNAs. Through comprehensive analysis, we constructed a ceRNA network based on the interactions between lncRNA-miRNA and miRNA-mRNA, which will help enhance our understanding of the pathogenesis of HAE, identify new therapeutic targets, improve treatment outcomes, and refine prognosis. Furthermore, our analysis of the Lnc-PCBP1-AS1-miR-20b-5p/CAPRIN2 axis revealed that overexpressed Lnc-PCBP1-AS1 may play a key role in the development and progression of HAE by adsorbing miR-20b-5p and thereby targeting the expression of CAPRIN2.

## Methods

### Patients and Samples

This study collected 13 cases of HAE patients who met the surgical resection criteria at Qinghai Provincial People’s Hospital from September 2021 to October 2023. The collected tissues included lesion center tissue, tissue <1cm around the lesion, and tissue >1cm around the lesion. The enrolled patients had no previous history of liver surgery and were not complicated with hepatic cystic echinococcosis, other parasitic diseases, liver benign or malignant tumors, liver abscess, liver metastatic cancer, or other space-occupying liver diseases. Both preoperative imaging and postoperative pathology diagnosed hepatic alveolar echinococcosis. After the specimens were removed, the required tissue samples were quickly cut and placed in RNA-free freezing tubes, then promptly stored in liquid nitrogen at −80°C for subsequent RNA extraction. This study was approved by the Ethics Committee of Qinghai Provincial People’s Hospital (approval number 2021-32). All participants voluntarily joined the project and signed an informed consent form. Table 1 presents the clinical data of the 13 HAE patients.

**Table 1.** Clinicopathological characteristics of 13 HAE patients.

| Clinicopathological characteristics | Patients (N = 13) |  |
| --- | --- | --- |
|  | N | % |
| **Age** |  |  |
| ≥40 | 7 | 53.8 |
| < 40 | 6 | 46.2 |
| **Gender** |  |  |
| Male | 5 | 38.5 |
| Female | 8 | 61.5 |
| **Lesion size** |  |  |
| ≥1040000 | 4 | 30.8 |
| < 1040000 | 9 | 69.2 |
| **Live vascular invasion** |  |  |
| Yes | 9 | 69.2 |
| No | 4 | 30.8 |
| **Multiple liver lesions** |  |  |
| Yes | 6 | 46.2 |
| No | 7 | 53.8 |
| **Distant metastasis** |  |  |
| Yes | 2 | 15.4 |
| No | 11 | 84.6 |

### RNA Extraction and Sequencing

Total RNA extraction was performed using TRIzol reagent (Thermo Scientific, USA). The RNA samples were assessed using 1% agarose gel electrophoresis, NanoPhotometer® spectrophotometer (Implen, CA, USA), Qubit® 3.0 Fluorometer (Implen, CA, USA), and Agilent 2100 RNA Nano 6000 Assay Kit (Implen, CA, USA). Three micrograms of total RNA from both the central lesion tissue and surrounding tissue of HAE patients were used as starting material for library construction. After library construction, initial quantification was done using Qubit 3.0, and the quality of the libraries was evaluated using the Agilent 2100, Bio-RAD CFX 96 real-time PCR instrument, and Bio-RAD KIT iQ SYBR GRN to ensure library quality. Clustering was followed by paired-end sequencing using the Illumina 4000 high-throughput sequencing platform (HiSeq/MiSeq). Uniquely mapped reads were used for gene read count and FPKM (Fragments Per Kilobase of transcript per Million mapped reads) calculation.

### Differentially Expressed Analysis

The R software package DESeq2 was utilized to calculate the differential expression of transcripts using a negative binomial distribution test. The cutoff criteria for identifying differentially expressed genes (DEGs) were set at a P-value <0.05 and a Fold Change(FC)>2/<0.5.

### Construction of a ceRNA Regulatory Network

The construction of the ceRNA network involved using the miRcode database (http://www.mircode.org/) to match differentially expressed lncRNAs (FC >2 or <0.5, *P* <0.05) with miRNAs. Based on the miRWalk database (http://mirwalk.umm.uni-heidelberg.de/), three bioinformatics algorithms—miRTarBase, miRDB, and TargetScan—were employed to predict the target mRNAs of miRNAs (FC >2 or <0.5, *P* <0.05). The predicted genes were then intersected with the genes obtained from sequencing. Following data integration, we constructed a comprehensive lncRNA-miRNA-mRNA regulatory network, which was visualized using Cytoscape (version v3.10.1).

Additionally, sub-networks were reconstructed using the BinGO plugin in Cytoscape^[19]^.

### Functional Enrichment and Protein–Protein Interaction Analysis

To gain a deeper understanding of the mechanisms by which differentially expressed mRNAs (DEmRNAs) contribute to the development of HAE, we utilized the “clusterProfiler” package in R software for functional annotation analysis. We also employed the ggplot2 package in R to create a bubble chart. *P* <0.05 was considered a threshold for significance. The protein-protein interaction (PPI) network was established using the STRING online search tool with a combined score of ≥0.4. The PPI network was visualized using Cytoscape (version v3.10.1).

### Real-Time Fluorescent Quantitative PCR (qRT-PCR)

We determined the relative expression levels of selected lncRNAs, miRNAs, and mRNAs using qRT-PCR. This study used GAPDH and U6 as internal references for qRT-PCR, and the 2^^-ΔΔCt^ method was employed to calculate the relative expression levels of the selected genes. The primer sequences for lncRNAs, mRNAs, and miRNAs, as well as the internal references, are listed in Table 2 and Table 3.

**Table 2.**
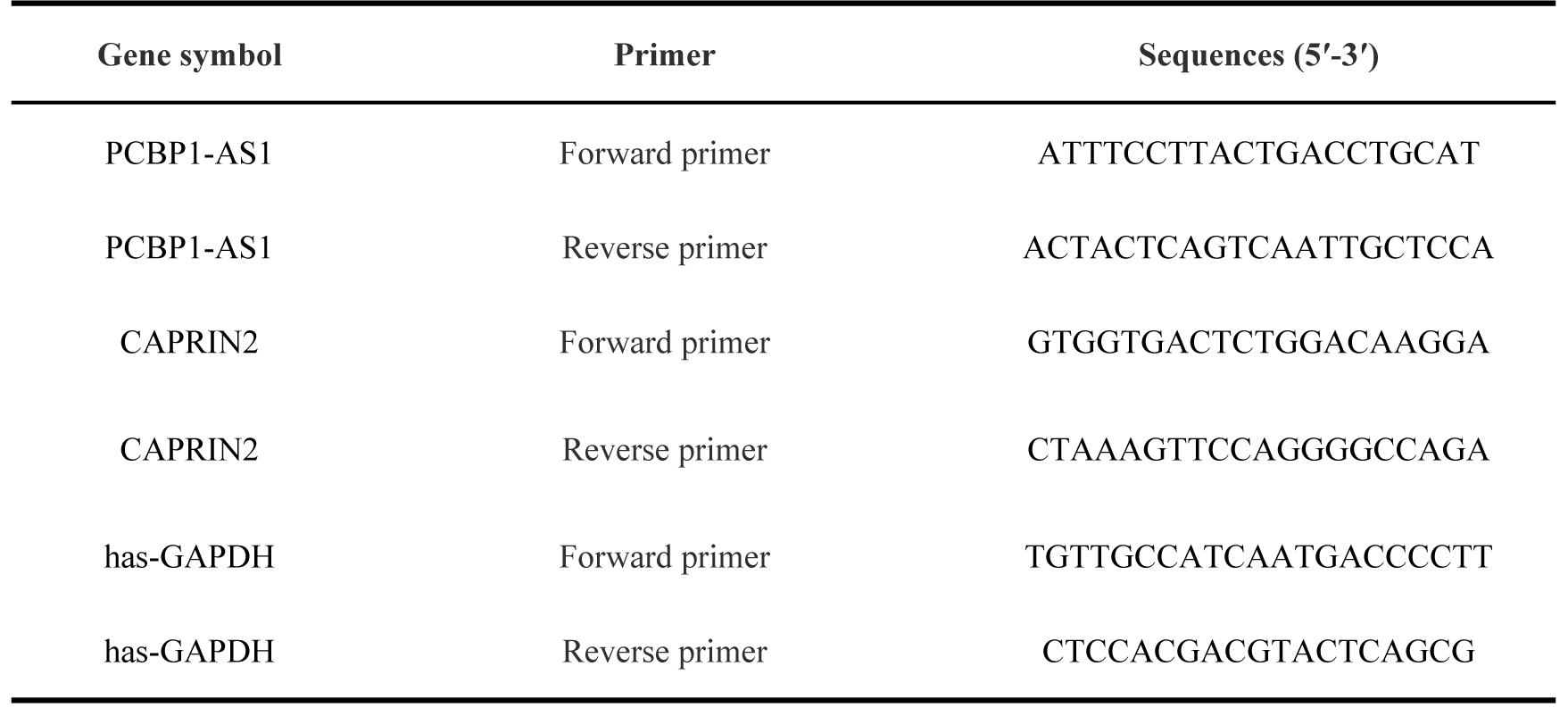
LncRNA and mRNA primer sequences.

**Table 3.**
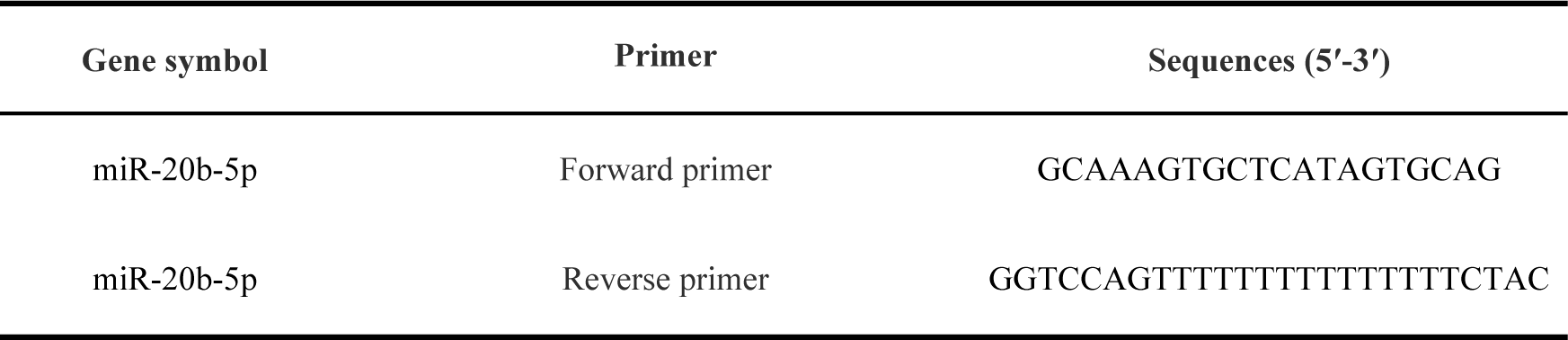
miRNA primer sequences.

### Cell culture and transfection

The 293T human embryonic kidney cell line was sourced from Promega, and the cells were cultured under conditions of DMEM medium supplemented with 10% FBS and 1% PS, at 37°C in a humidified atmosphere with 5% CO2. The overexpression plasmids for hsa-miR-20b-5p and the control plasmids were purchased from GeneCopoeia, while the wild-type and mutant plasmids for PCBP1-AS1 and CAPRIN2 were obtained from Universal Bio. Transfection of 293T human embryonic kidney cells was carried out using Lipofectamine 3000 reagent (Invitrogen, Shanghai, China), with 0.5μg of plasmid, 0.05μM of miRNA, and 1μL of P3000™ reagent. Subsequently, the Dual-Glo® Luciferase assay system was used to measure the luminescence of both firefly and Renilla luciferases. Luciferase activity was represented by the ratio of firefly to Renilla luciferase luminescence intensity. The experiment was conducted independently in triplicate.

### Statistical analysis

Data were statistically analyzed using SPSS 28.0 software (IBM, Armonk, NY, USA) or GraphPad Prism 9.0 (GraphPad Software, San Diego, CA, USA). Non-parametric data by group were analyzed using the Friedman rank-sum test, and experimental data are presented in the form of *M* (*P*_25_, *P*_75_). Student’s t-test or one-way analysis of variance (ANOVA) was used to analyze differences between two experimental groups or among more than two groups, respectively. Data were expressed as the mean ± standard deviation (SD). *P* <0.05 was considered statistically significant.

## Results

### Screening for the Coding Potential of LncRNAs

We sequenced lncRNAs from the central lesion tissue and surrounding tissue of three patients with human HAE. Since lncRNAs are RNAs that do not encode proteins, we predicted their coding potential to differentiate RNAs with encoding functions from those without encoding potential in the sequencing data. To distinguish lncRNAs, we utilized a comprehensive set of the most widely applied coding potential analysis methods, including: CPC analysis^[20]^, CNCI analysis ^[21]^, pfam protein domain analysis ^[22]^, and PLEK analysis ^[23]^. Specifically, CPC analysis identified 9,046 lncRNAs, CNCI analysis identified 35,119 lncRNAs, pfam protein domain analysis identified 37,943 lncRNAs, and PLEK analysis identified 2,602 lncRNAs. By integrating the predictions from these four methods, we obtained a total of 26,365 lncRNAs. (Figure 1)

**Figure 1:**
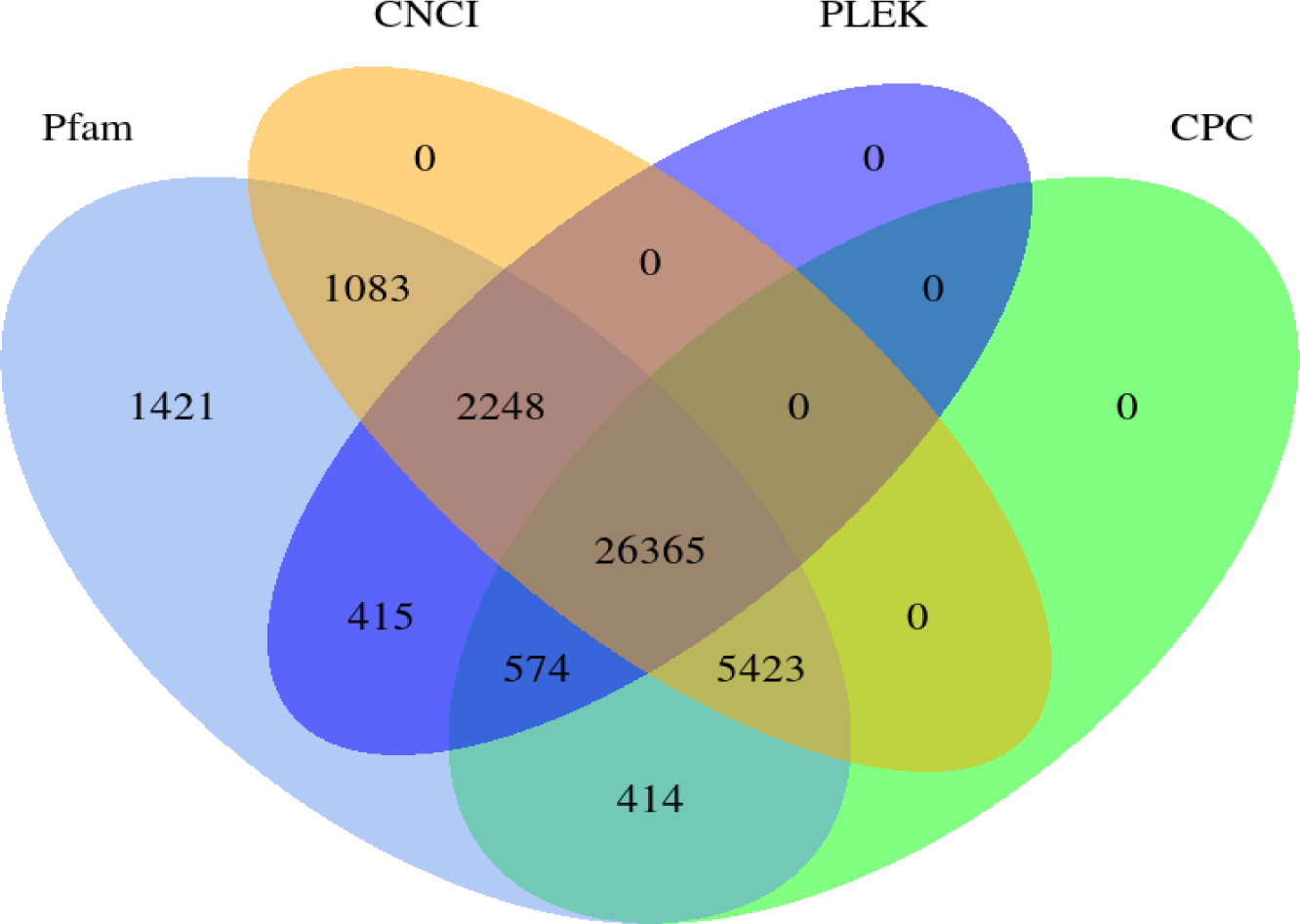
Venn diagram showing the results of four coding potential prediction methods.

### LncRNA expression profiles in HAE

To date, tens of thousands of lncRNA genes have been identified in humans and single-celled eukaryotic organisms. Based on the relative positional relationship between lncRNAs and protein-coding genes, lncRNAs are classified into intergenic lncRNA (lincRNA), intronic lncRNA, antisense lncRNA, sense lncRNA, and bidirectional lncRNA^[24]^. According to the lncRNA sequencing results, there are 9,022 intronic lncRNAs (denoted as ‘i’), 9,057 sense overlapping lncRNAs (denoted as ‘o’), 5,733 intergenic lncRNAs (denoted as ‘u’), and 2,553 antisense lncRNAs (denoted as ‘x’), with the majority containing two or more exons (Figure 2).

**Figure 2:**
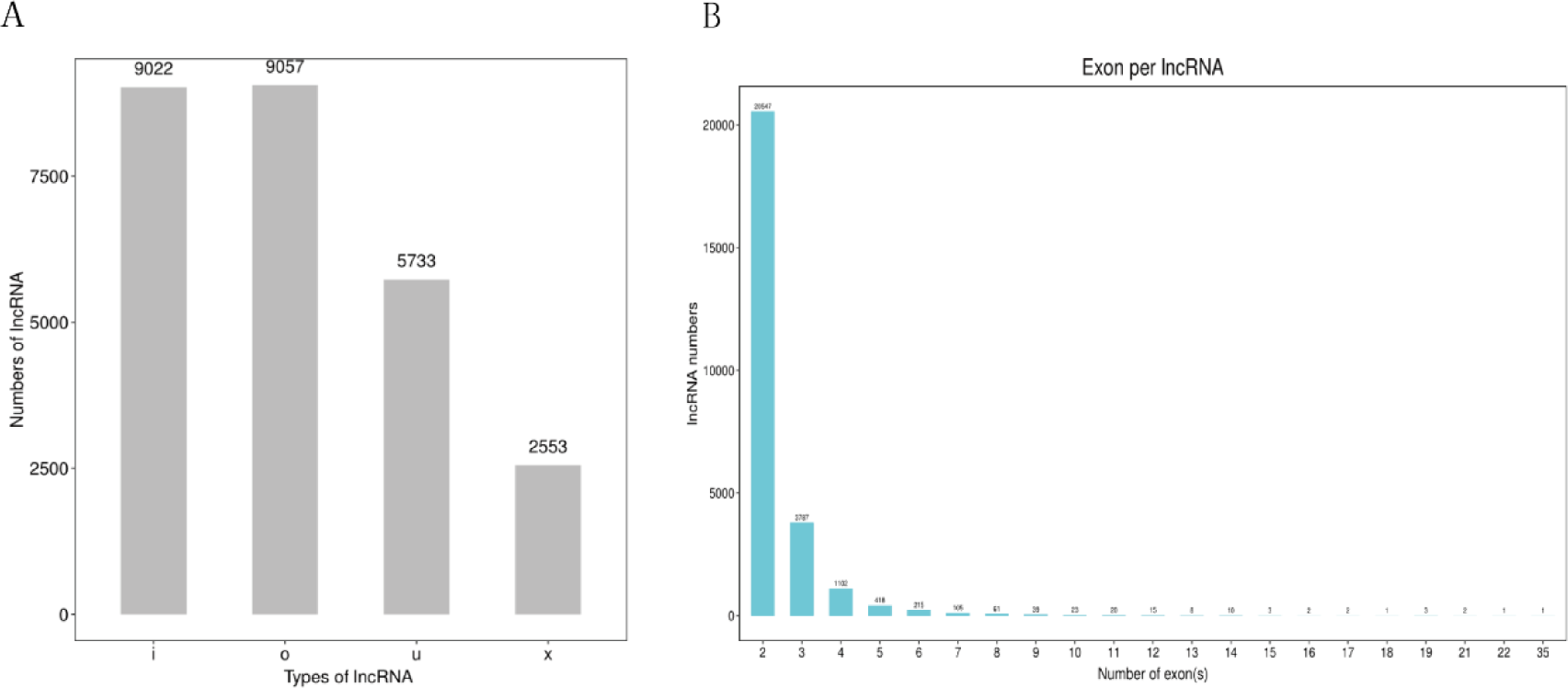
Characteristics of LncRNAs in HAE sequencing. (A)Distribution bar chart of lncRNA types. (B) Bar chart of the number of exons contained in lncRNA.

### The selection of DELncRNAs in HAE

To investigate the expression of LncRNAs in patients with HAE, high-throughput sequencing technology was employed, with a P-value <0.05 and a fold change (FC) >2 or FC <0.5 as the criteria for screening differentially expressed LncRNAs (DElncRNAs). A total of 979 DElncRNAs were identified (591 upregulated and 388 downregulated). These differentially expressed LncRNAs are displayed as a heatmap (Figure 3A) and a volcano plot (Figure 3B), they exhibit good discrimination and clustering between the central lesion tissue group and the surrounding lesion tissue group. The top 10 up-regulated and down-regulated DElncRNAs with their names, FoldChange, Log_2_FoldChange and P-Value are listed in Table 4. All DElncRNAs with their names, Fold Change, Log_2_FoldChange and P-Value are listed in Supplementary Table 1.

**Figure 3:**
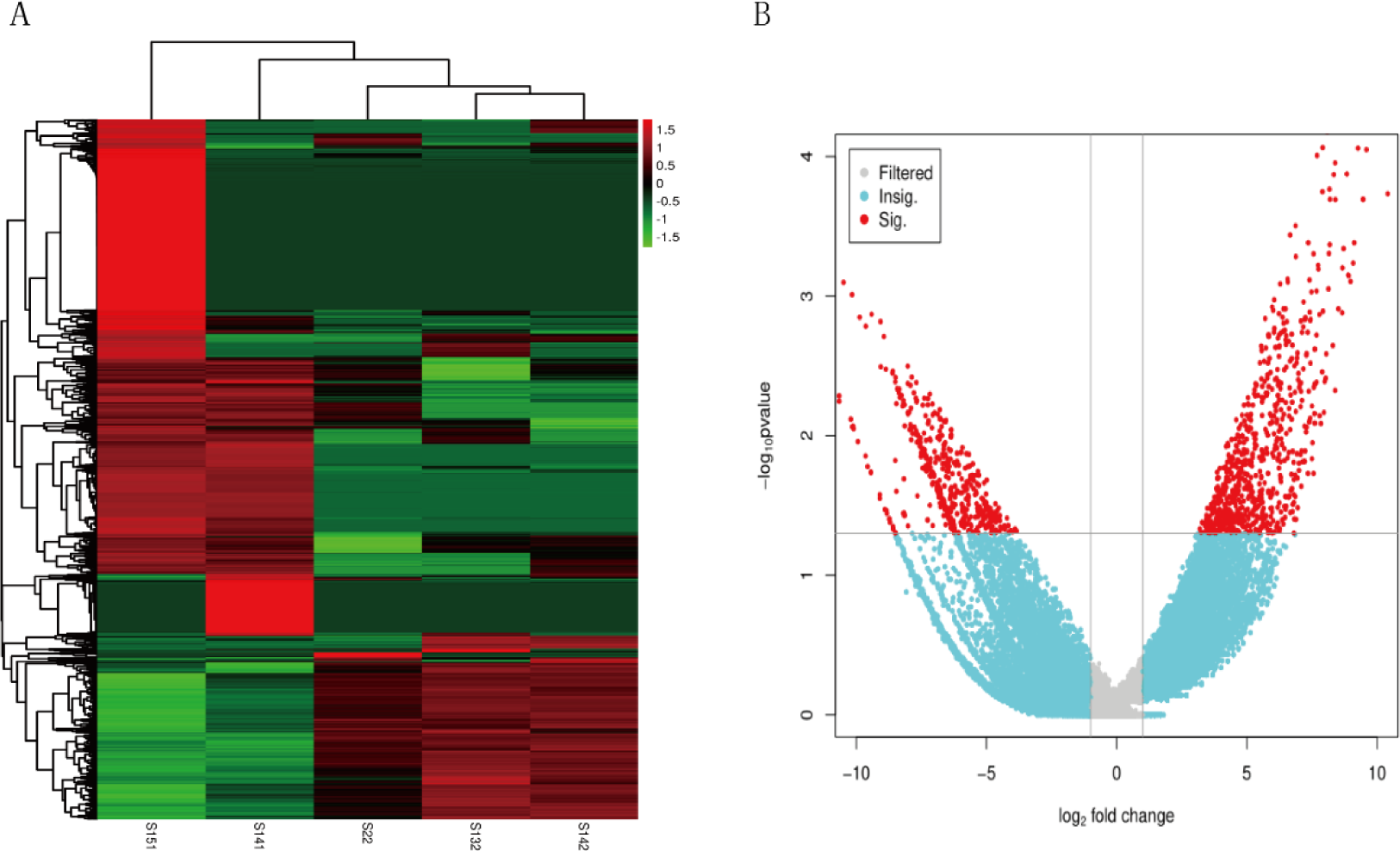
Identification of DELncRNAs in HAE sequencing from lesion tissue and surrounding tissue. (A) Heatmap of DElncRNAs. (B) Volcano plot of DElncRNAs. Red represents upregulated genes, blue represents downregulated genes, and gray represents genes without significant differences.

**Table 4.**
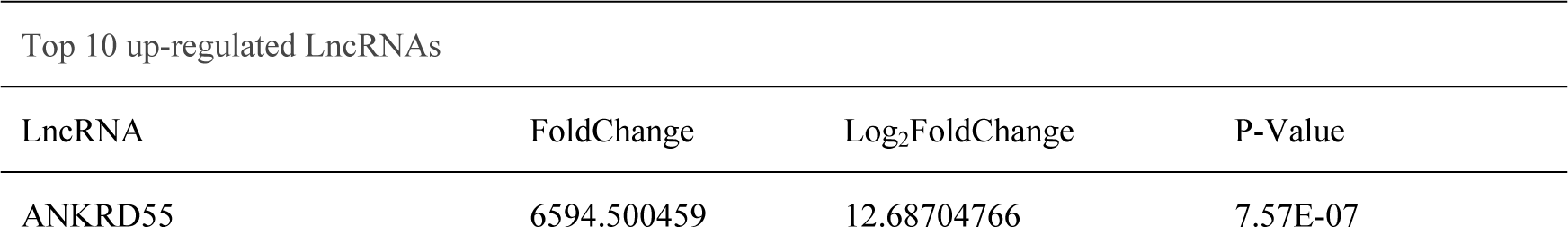
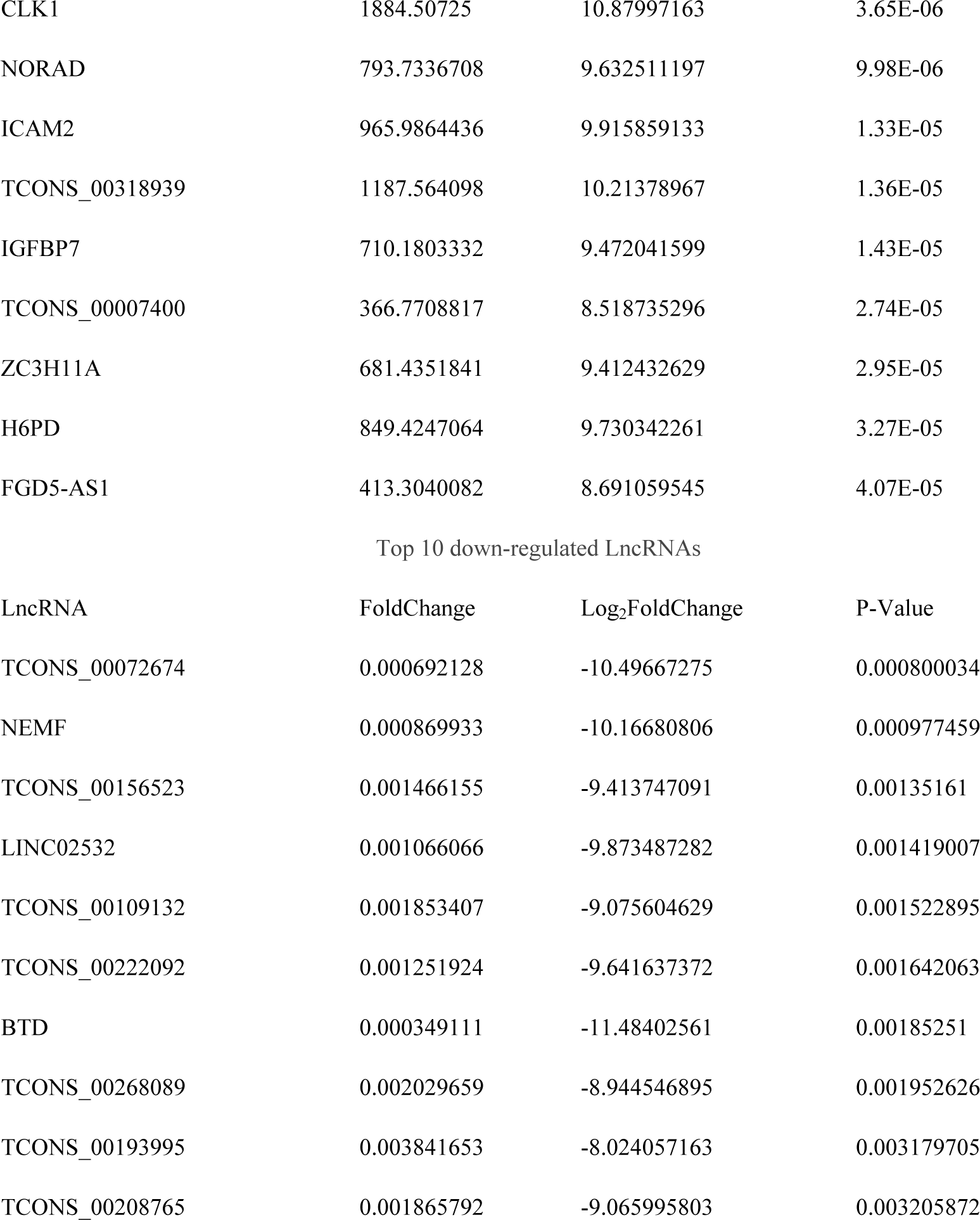
Top 10 up-regulated and down-regulated lncRNAs.

| Top 10 up-regulated LncRNAs |  |  |  |
| --- | --- | --- | --- |
| LncRNA | FoldChange | Log<sub>2</sub>FoldChange | P-Value |
| ANKRD55 | 6594.500459 | 12.68704766 | 7.57E-07 |
| CLK1 | 1884.50725 | 10.87997163 | 3.65E-06 |
| NORAD | 793.7336708 | 9.632511197 | 9.98E-06 |
| ICAM2 | 965.9864436 | 9.915859133 | 1.33E-05 |
| TCONS_00318939 | 1187.564098 | 10.21378967 | 1.36E-05 |
| IGFBP7 | 710.1803332 | 9.472041599 | 1.43E-05 |
| TCONS_00007400 | 366.7708817 | 8.518735296 | 2.74E-05 |
| ZC3H11A | 681.4351841 | 9.412432629 | 2.95E-05 |
| H6PD | 849.4247064 | 9.730342261 | 3.27E-05 |
| FGD5-AS1 | 413.3040082 | 8.691059545 | 4.07E-05 |

Top 10 down-regulated lncRNAs
| lncRNA | FoldChange | Log<sub>2</sub>FoldChange | P-Value |
| --- | --- | --- | --- |
| TCONS_00072674 | 0.000692128 | -10.49667275 | 0.000800034 |
| NEMF | 0.000869933 | -10.16680806 | 0.000977459 |
| TCONS_00156523 | 0.001466155 | -9.413747091 | 0.00135161 |
| LINC02532 | 0.001066066 | -9.873487282 | 0.001419007 |
| TCONS_00109132 | 0.001853407 | -9.075604629 | 0.001522895 |
| TCONS_00222092 | 0.001251924 | -9.641637372 | 0.001642063 |
| BTD | 0.000349111 | -11.48402561 | 0.00185251 |
| TCONS_00268089 | 0.002029659 | -8.944546895 | 0.001952626 |
| TCONS_00193995 | 0.003841653 | -8.024057163 | 0.003179705 |
| TCONS_00208765 | 0.001865792 | -9.065995803 | 0.003205872 |

### DEmRNAs in HAE

Most lncRNAs can act as miRNA sponges, indirectly regulating downstream target genes. To explore the potential “ceRNA” mechanism of lncRNAs in HAE and to reveal the differentially expressed mRNAs in the same tissues, our research group also conducted mRNA sequencing. A total of 870 mRNAs (743 upregulated and 127 downregulated) were identified as differentially expressed RNAs (DERNAs) in HAE (*P* <0.05, FC >2 / <0.5). These differentially expressed mRNAs are displayed as a heatmap (Figure 4A) and a volcano plot (Figure 4B). The top 10 up-regulated and down-regulated DEmRNAs with their names, FoldChange, Log_2_FoldChange and P-Value are listed in Table 5. All DEmRNAs with their names, Fold Change, Log_2_FoldChange and P-Value are listed in Supplementary Table 2.

**Figure 4:**
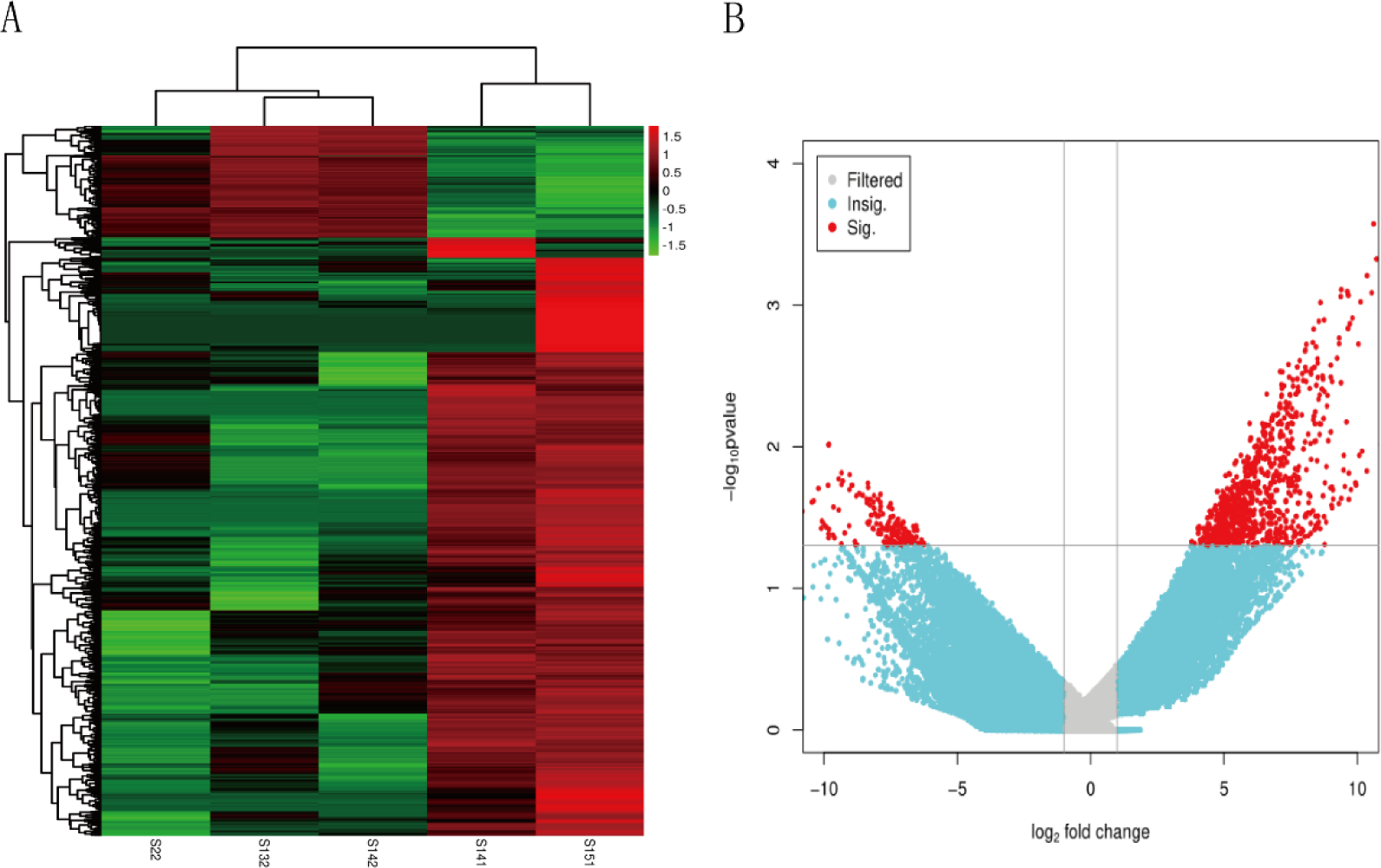
Identification of DEmRNAs in HAE sequencing from lesion tissue and surrounding tissue. (A) Heatmap of DEmRNAs. (B) Volcano plot of DEmRNAs. Red represents upregulated genes, blue represents downregulated genes, and gray represents genes without significant differences.

**Table 5.** Top 10 up-regulated and down-regulated mRNAs.

| Top 10 up-regulated mRNAs |  |  |  |
| --- | --- | --- | --- |
| mRNA | FoldChange | Log<sub>2</sub>FoldChange | P-Value |
| MIER1 | 6891.139911 | 12.75052693 | 5.86E-05 |
| NCK1 | 4520.924017 | 12.14240196 | 0.000119392 |
| METTL9 | 3199.851264 | 11.64378913 | 0.000214831 |
| GTF2H2C | 1561.441829 | 10.60866311 | 0.000267621 |
| IPMK | 1317.737516 | 10.36384731 | 0.000618708 |
| FCRL5 | 673.0462802 | 9.394561901 | 0.000778941 |
| SUB1 | 1486.820226 | 10.5380145 | 0.000819456 |
| MORC4 | 1114.003657 | 10.12153825 | 0.000947739 |
| CXorf40B | 899.6704652 | 9.813252852 | 0.00123812 |
| SPART | 429.6831405 | 8.747129361 | 0.0012714 |
| Top 10 down-regulated mRNAs |  |  |  |
| mRNA | FoldChange | Log<sub>2</sub>FoldChange | P-Value |
| CES2 | 0.001101447 | -9.826384333 | 0.009603027 |
| SURF4 | 0.001096504 | -9.832873188 | 0.00970853 |
| MDM4 | 0.00153438 | -9.348128419 | 0.015256378 |
| TPT1 | 0.001892148 | -9.045759259 | 0.015752727 |
| SYAP1 | 0.001429811 | -9.449960054 | 0.016911442 |
| SPPL2A | 0.001595592 | -9.291692341 | 0.017513725 |
| SLC7A2 | 0.000359791 | -11.44055405 | 0.017830314 |
| PCYOX1 | 0.003043029 | -8.36027622 | 0.018166061 |
| LDLR | 0.001535247 | -9.347313565 | 0.018465857 |
| WDR36 | 0.003073114 | -8.346082935 | 0.019419049 |

### Construction of the ceRNA Network in HAE

As previously mentioned, lncRNAs exert their functions primarily by adsorbing miRNAs. Therefore, based on high-throughput sequencing analysis, we constructed a comprehensive lncRNA-miRNA-mRNA network to study the roles of differentially expressed lncRNAs and mRNAs in HAE. Through the interactions of lncRNA-miRNA and miRNA-mRNA, we obtained a total of 11 lncRNAs, 21 miRNAs, and 56 mRNAs. The constructed lncRNA-miRNA-mRNA ceRNA regulatory network includes 88 nodes and 183 edges (Figure 5A). It is well known that lncRNAs and mRNAs have co-expression patterns in the ceRNA network. Thus, we selected a hub lncRNA (degree ≥16) and its connected miRNAs and mRNAs to reconstruct a sub-network. As shown in (Figure 5B), the PCBP1-AS1-miRNA-mRNA sub-network consists of 1 lncRNA node, 16 miRNA nodes, 51 mRNA nodes, and 91 edges. The DElncRNA and its matching miRNA and 21 miRNAs and their matching DEmRNAs can be found in (Table 6 and Table7). The degree of the ceRNA network were listed in Supplementary Table 3.

**Figure 5:**
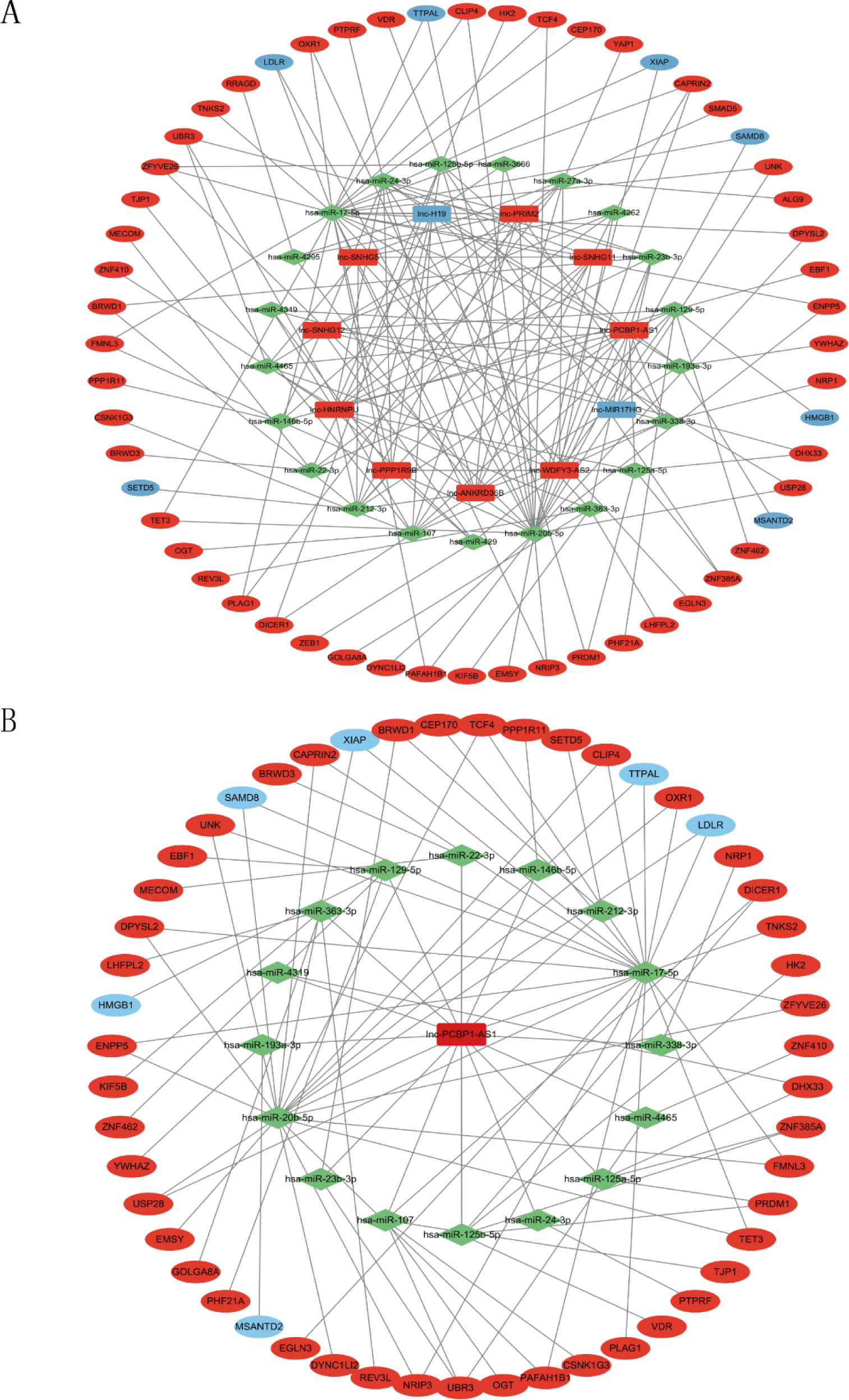
Construction of lncRNA-miRNA-mRNA network in HAE. (A) lncRNA-miRNA-mRNA ceRNA network in HAE. (B) The lncRNA PCBP1-AS1 sub-network. Red nodes indicate up-regulated RNAs while blue nodes indicate down-regulated RNAs. lncRNAs are denoted by rectangles, miRNAs by light green diamonds, and mRNAs by ellipses.

**Table 6.** DElncRNAs in the ceRNA network were targeted by miRNAs.

| DELncRNA | miRNA |
| --- | --- |
| lnc-H19 | hsa-miR-107,hsa-miR-193a-3p,hsa-miR-338-3p,hsa-miR-22-3p,hsa-miR-4295,hsa-miR-3666, hsa-miR-17-5p,hsa-miR-20b-5p,hsa-miR-212-3p,hsa-miR-24-3p |
| lnc-PRIM2 | hsa-miR-146b-5p, hsa-miR-27a-3p, hsa-miR-4465, hsa-miR-23b-3p, hsa-miR-107, hsa-miR-338-3p, hsa-miR-4295, hsa-miR-3666, hsa-miR-17-5p, hsa-miR-20b-5p, hsa-miR-212-3p, hsa-miR-24-3p |
| lnc-SNHG11 | hsa-miR-22-3p, hsa-miR-107, hsa-miR-17-5p, hsa-miR-20b-5p, hsa-miR-212-3p, hsa-miR-24-3p |
| lnc-PCBP1-AS1 | hsa-miR-363-3p, hsa-miR-129-5p, hsa-miR-125a-5p, hsa-miR-146b-5p, hsa-miR-193a-3p, hsa-miR-4465, hsa-miR-4319, hsa-miR-125b-5p, hsa-miR-23b-3p, hsa-miR-338-3p, hsa-miR-22-3p, hsa-miR-107, hsa-miR-17-5p, hsa-miR-20b-5p, hsa-miR-212-3p, hsa-miR-24-3p |
| lnc-WDFY3-AS2 | hsa-miR-4262, hsa-miR-27a-3p, hsa-miR-129-5p, hsa-miR-429, hsa-miR-4465, hsa-miR-23b-3p, hsa-miR-338-3p, hsa-miR-107, hsa-miR-24-3p |
| lnc-ANKRD36B | hsa-miR-193a-3p,hsa-miR-4295,hsa-miR-3666,hsa-miR-4262,hsa-miR-27a-3p,hsa-miR-129-5p,hsa-miR-338-3p,hsa-miR-4465,hsa-miR-107,hsa-miR-17-5p,hsa-miR-20b-5p,hsa-miR-212-3p, hsa-miR-24-3p |
| lnc-PPP1R9B | hsa-miR-125a-5p, hsa-miR-17-5p, hsa-miR-20b-5p, hsa-miR-212-3p, hsa-miR-24-3p, hsa-miR-4319, hsa-miR-125b-5p, hsa-miR-429 |
| lnc-SNHG12 | hsa-miR-146b-5p, hsa-miR-193a-3p, hsa-miR-429, hsa-miR-4262, hsa-miR-129-5p, hsa-miR-338-3p, hsa-miR-24-3p |
| lnc-SNHG5 | hsa-miR-363-3p, hsa-miR-27a-3p, hsa-miR-23b-3p, hsa-miR-4262, hsa-miR-4465, hsa-miR-212-3p |
| lnc-MIR17HG | hsa-miR-23b-3p |
| lnc-HNRNPU | hsa-miR-363-3p, hsa-miR-4262, hsa-miR-125a-5p, hsa-miR-27a-3p, hsa-miR-4319, hsa-miR-125b-5p, hsa-miR-129-5p, hsa-miR-429, hsa-miR-4465, hsa-miR-23b-3p, hsa-miR-212-3p, hsa-miR-24-3p |

**Table 7.** The 21 miRNAs with their target DEmRNAs in the ceRNA network.

| miRNA | DEmRNA |
| --- | --- |
| hsa-miR-107 | DICER1, OGT, CSNK1G3, TJP1, UBR3 |
| hsa-miR-125a-5p | DHX33, ZNF385A, PRDM1 |
| hsa-miR-146b-5p | PPP1R11 |
| hsa-miR-17-5p | USP28, TCF4, TET3, PAFAH1B1, FMNL3, XIAP, DPYSL2, ZFYVE26, LDLR, TTPAL, ENPP5, NRIP3, EGLN3 |
| hsa-miR-193a-3p | YWHAZ, MSANTD2 |
| hsa-miR-20b-5p | CAPRIN2, EMSY, SP28, TCF4, TET3, PAFAH1B1, FMNL3, DPYSL2, LDLR, TTPAL, ENPP5, NRIP3, SAMD8 |
| hsa-miR-212-3p | SETD5, BRWD1 |
| hsa-miR-24-3p | PTPRF |
| hsa-miR-27a-3p | YAP1, SMAD5, ALG9, PLAG1 |
| hsa-miR-338-3p | NRP1 |
| hsa-miR-363-3p | LHFPL2, KIF5B, GOLGA8A, REV3L |
| hsa-miR-4465 | UBR3, ZNF410 |
| hsa-miR-4319 | DHX33 |
| hsa-miR-125b-5p | ZNF385A, VDR, HK2, DICER1, PRDM1 |
| hsa-miR-23b-3p | BRWD1, UBR3 |
| hsa-miR-129-5p | EBF1, ZNF462, PHF21A, HMGB1 |
| hsa-miR-429 | ZEB1 |
| hsa-miR-22-3p | MECOM, BRWD3 |
| hsa-miR-4295 | RRAGD |
| hsa-miR-3666 | ZFYVE26 |
| hsa-miR-4262 | CAPRN2 |

### Functional Enrichment Analysis and PPI Network Construction

GO and KEGG enrichment analyses indicate that the DEmRNAs in the ceRNA network are significantly associated with the positive regulation of canonical Wnt signaling pathway, histone lysine methylation, and the positive regulation of the Wnt signaling pathway, among other biological processes (BP) (Figure 6A). The most enriched Cellular Component (CC) was the cellular microtubule (Figure 6B). The enrichment of Molecular Functions (MF) was primarily related to histone methyltransferase activity (H3-K9 specific), histone-lysine N-methyltransferase activity and microtubule binding (Figure 6C). DEmRNAs related to the ceRNA network were significantly enriched in 5 KEGG pathways, namely adherens junction, platinum drug resistance, insulin resistance, HIF-1 signaling pathway, and toxoplasmosis (Figure 6D). The top 10 results of the functional enrichment analysis are listed in the Supplementary Table 4. The enrichment analysis suggests that the HAE-specific ceRNA network may participate in the pathogenesis and development of HAE by regulating these biological processes and pathways.

**Figure 6:**
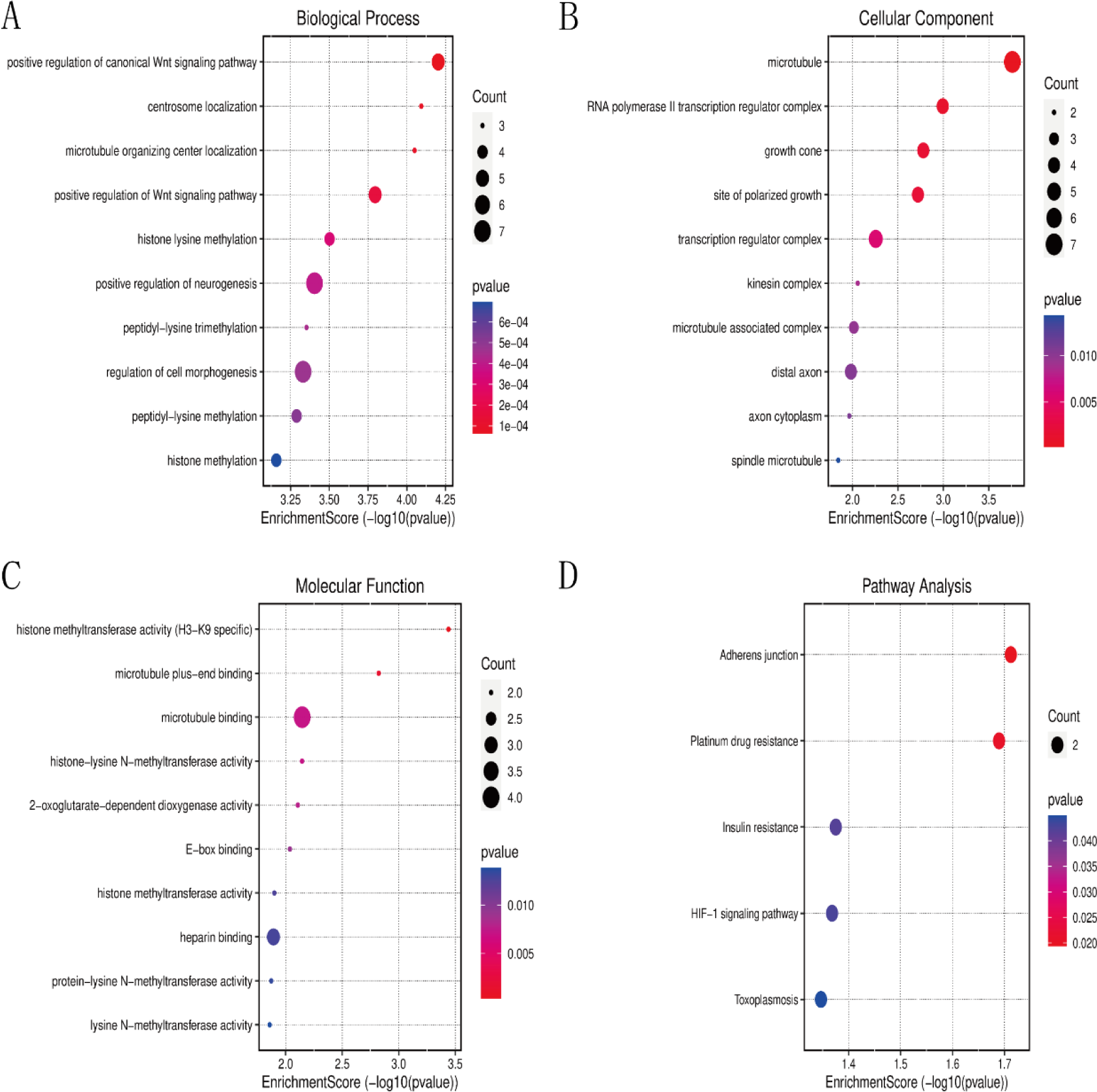
GO and KEGG enrichment analysis of DEmRNAs in the ceRNA network. (Top10). (A) Bubble Plot of BP. (B) Bubble Plot of CC. (C) Bubble Plot of MF. (D) Bubble Plot of KEGG.

Additionally, further analysis was conducted on the interacting mRNAs within the ceRNA network, leading to the construction of a Protein-Protein Interaction (PPI) network. The PPI network comprises 19 nodes and 17 edges (Figure 7). These mRNAs are likely to play a significant role in the disease occurrence and progression of HAE and serve as subjects for subsequent research. Notably, genes with high network degree and comprehensive scores within the network, such as YAP1, ZEB1, PAFAH1B1, TJP1 and YWHA, were found to be primarily enriched in biological processes like “ Regulation of developmental process” and “Positive regulation of developmental process”. This suggests that these genes are crucial for the development and progression of HAE.

**Figure 7:**
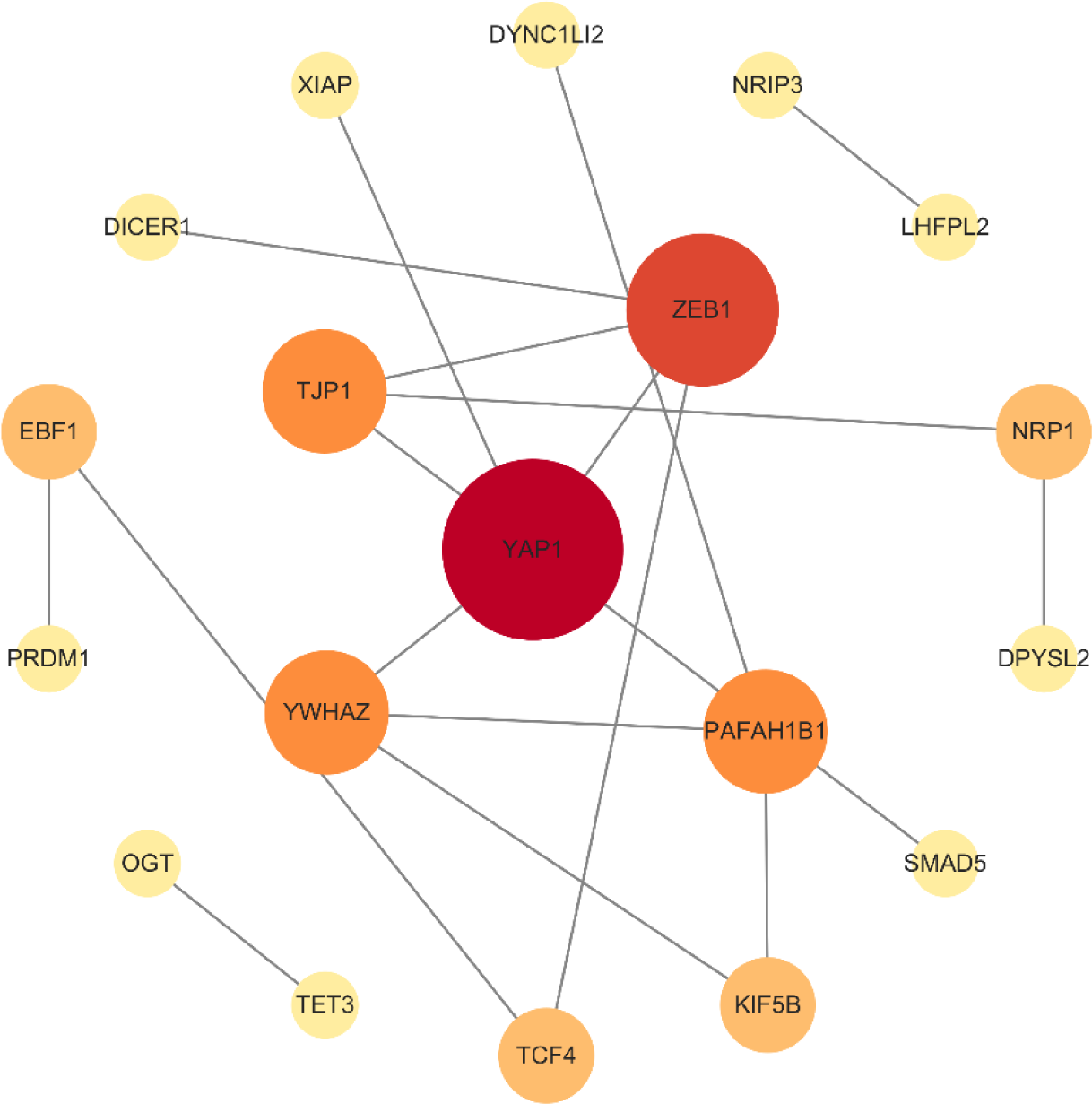
The PPI network of DEmRNAs within the ceRNA network. Circles represent DEmRNAs, lines represent interactions between encoded proteins, gradient from dark red to light yellow based on degree.

### Validation of screened lncRNA-PCBP1-AS1, miR-20b-5p and CAPRIN2 by qRT-PCR

We utilized qRT-PCR to analyze the expression levels of PCBP1-AS1, miR-20b-5p, and CAPRIN2 in 13 HAE lesion tissues, as well as in tissues less than 1cm and more than 1cm surrounding the lesion. The qRT-PCR results indicated that PCBP1-AS1 and CAPRIN2 were highly expressed (*P* <0.05) in the lesion tissues (Figure 8A and Figure 8B), which is consistent with the results from high-throughput sequencing; whereas miR-20b-5p exhibited low expression(*P* <0.05) in the HAE lesion tissues (Figure 8C).

**Figure 8:**
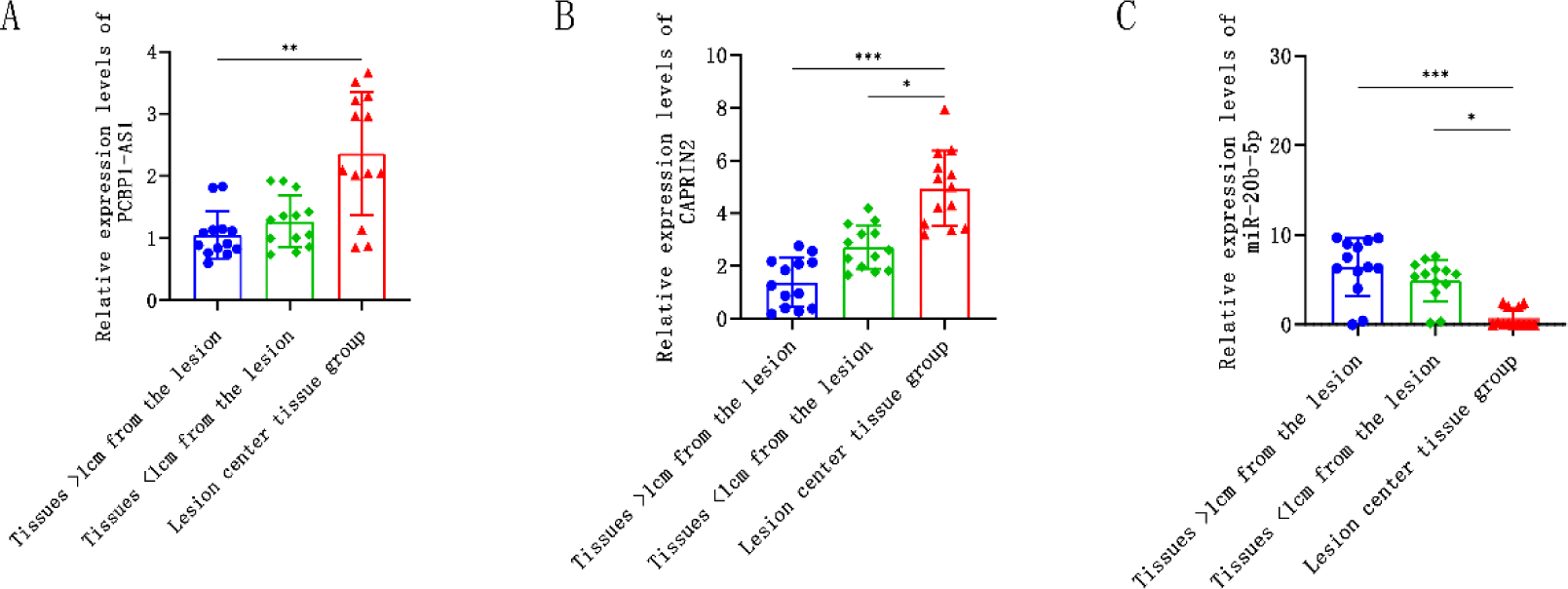
Expression of Candidate gene in HAE tissues assessed by qRT-PCR.(A) PCBP1-AS1 expression in HAE;(B) CAPRIN2 expression in HAE;(C) miR-20b-5p expression in HAE. The data are shown as the Median and interquartile range (IQR). n=13. \**P* <0.05, \*\**P* <0.01, \*\*\**P* <0.001.

### miR-20b-5p is directly regulated by PCBP1-AS1 and interacts with CAPRIN2

In the ceRNA network, there is a targeted binding relationship among PCBP1-AS1, miR-20b-5p, and CAPRIN2. The sequencing data show a consistent expression trend for PCBP1-AS1 and CAPRIN2, and qRT-PCR indicates that both PCBP1-AS1 and CAPRIN2 are overexpressed in the lesion tissues, while miR-20b-5p is underexpressed in the same tissues. Subsequently, we used the RNAhybrid (https://bibiserv.cebitec.uni-bielefeld.de/rnahybrid) and Targetscan databases(https://www.targetscan.org/) to predict potential binding sites between PCBP1-AS1 and miR-20b-5p, as well as between miR-20b-5p and CAPRIN2 (Figure 9A and Figure 9B), and calculated the minimum free energy of these binding sites using RNAhybrid (Figure 9C). In order to further confirm the predicted results, we transfected them into 293T cells. The results showed that miR-20b-5p could decrease the fluorescence values in cells transfected with PCBP1-AS1-WT and CAPRIN2-WT. In contrast, there was no significant change in fluorescence values in cells transfected with PCBP1-AS1-MUT and CAPRIN2-MUT (Figure 9D and Figure 9E). These results suggest that PCBP1-AS1 directly targets miR-20b-5p, and miR-20b-5p interacts with CAPRIN2, with miR-20b-5p expression being negatively correlated with that of PCBP1-AS1 and CAPRIN2.

**Figure 9:**
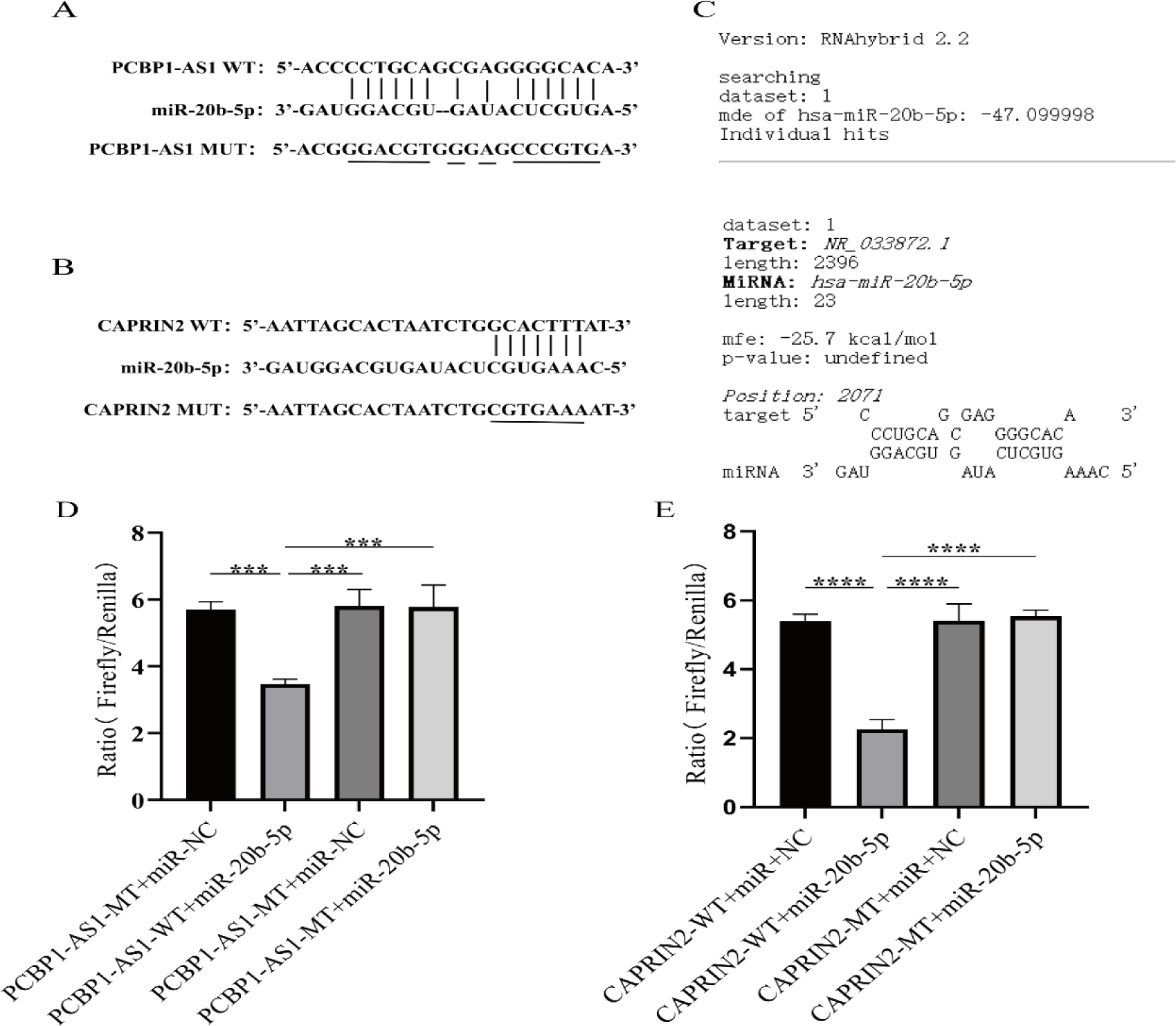
(A) The binding sites between miR-20b-5p and PCBP1-AS1;(B) The binding sites between miR-20b-5p and CAPRIN2;(C) The interaction between THAP9-AS1 and miR-484 and the minimum free energy (mfe) was predicted with RNAhybrid software;(D) Results of the dual-luciferase assay for miR-20b-5p and PCBP1-AS1;(E) Results of the dual-luciferase assay for miR-20b-5p and CAPRIN2. The data are shown as the Mean ± standard deviation. (n=3). \*\*\**P* <0.001, \*\*\*\**P* <0.0001.

### Analysis of CAPRIN2 expression in HAE lesions and Clinicopathological characteristics

Through qRT-PCR and dual-luciferase reporter assays, the targeted regulatory relationship among PCBP1-AS1, miR-20b-5p, and CAPRIN2 has been demonstrated. Specifically, PCBP1-AS1 can affect the expression of the CAPRIN2 gene by adsorbing miR-20b-5p. To further investigate the association between this regulatory relationship and the clinical characteristics of HAE patients, we compared the expression levels of CAPRIN2 with various clinical parameters of the patients. The analysis revealed that overexpression of CAPRIN2 was not significantly correlated with the patient’s sex, age, multiple intrahepatic lesions, or distant metastasis of HAE (*P* > 0.05), but it was significantly correlated with the size of HAE lesions and intrahepatic vascular invasion (*P* <0.05) (Figure 10), indicating that patients with overexpressed CAPRIN2 may have a faster disease progression rate than those with low CAPRIN2 expression. This could imply that the expression level of CAPRIN2 could serve as a potential biomarker for predicting the prognosis of HAE patients.

**Figure 10.**
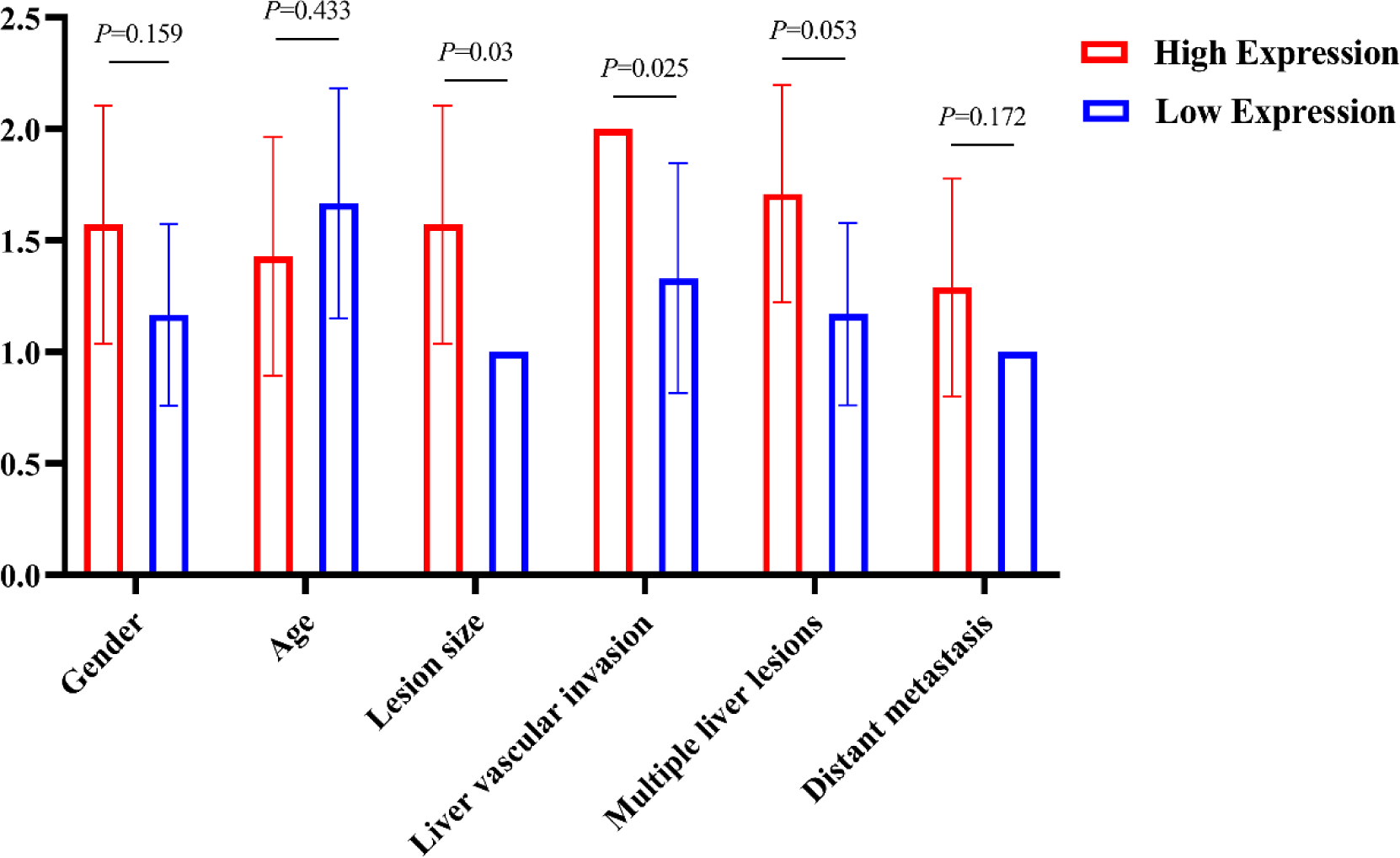
The expression of CAPRIN2 correlated significantly with Clinicopathological characteristics.

These findings suggest that the regulatory axis of PCBP1-AS1-miR-20b-5p/CAPRIN2 may play a key role in controlling the progression and vascular invasion of HAE, which is of great significance for understanding the molecular mechanisms of HAE and developing new therapeutic strategies. Future research can further explore the specific mechanisms of action of this molecular axis and assess its feasibility as a potential therapeutic target.

## Discussion

HAE is a zoonotic disease with severe clinical manifestations and poor prognosis. Currently, radical surgery combined with albendazole medication remains the most ideal treatment option for HAE patients^[25][26]^. In recent years, numerous studies have gradually revealed the ability of LncRNAs to regulate the expression of target genes through various pathways, playing a significant role in promoting the occurrence, development, and metastasis of tumors. Moreover, the ceRNA hypothesis has proposed an innovative regulatory mechanism, where lncRNAs act as ceRNAs and form extensive regulatory networks by competitively binding to endogenous miRNAs. The imbalance of such ceRNA regulatory networks can significantly affect the biological functions of an organism, leading to the onset of diseases. Additionally, multiple studies have confirmed that ceRNA network play a crucial role in the development of various tumors. However, in HAE, the ceRNA network mediated by lncRNAs has not yet been comprehensively studied, and many ceRNA molecules still await discovery and understanding.

In this study, we utilized high-throughput sequencing technology to precisely identify DElncRNAs and DEmRNAs in HAE lesion tissues and their adjacent tissues. We attempted to screen for ceRNA molecules with specific functions using the hierarchical action model of lncRNA-miRNA-mRNA. By integrating the interactions between miRNAs, DEmRNAs, and DElncRNAs, we constructed an HAE-characteristic ceRNA regulatory network, which includes 89 key nodes and 183 interacting edges. Furthermore, we identified hub LncRNAs and built a sub-network centered around PCBP1-AS1.

Furthermore, we conducted GO and KEGG enrichment analyses to delve deeper into the biological pathways and functions involved in the ceRNA network for DEmRNAs. The GO enrichment analysis revealed that DEmRNAs in the ceRNA network are primarily involved in the positive regulation of the Wnt signaling pathway and histone lysine methylation, key biological processes that may be significantly associated with the occurrence of HAE. At the molecular level, the DEmRNAs are mainly associated with histone methyltransferase activity (specifically H3-K9) and histone lysine N-methyltransferase activity. Notably, DNA methyltransferase 3A (DNMT3A) is related to histone methyltransferase activity, and DNA methyltransferases are enzymes responsible for methylating DNA in mammals, which can lead to gene silencing^[27]^. Moreover, a multitude of histone lysine methyltransferases catalyze various lysine methylation events, decorating the core histones, and their aberrant expression is frequently observed in cancers, developmental disorders, and other pathologies^[28]^. In simple terms, HAE may be related to abnormalities in the intracellular epigenetic regulatory mechanisms, which affect gene expression and cellular function.

The KEGG pathway enrichment analysis results further revealed the roles of DEmRNAs involved in the ceRNA network across multiple biological pathways, such as adherens junction, platinum drug resistance, insulin resistance, HIF-1 signaling pathway, and toxoplasmosis. The enrichment of these pathways suggests that the pathogenesis of HAE may involve several complex biological processes. It is particularly noteworthy that adherens junctions play a critical role in maintaining the integrity of migrating cell groups, facilitating cell coordination, and cell rearrangement^[29]^. They are also implicated in the collective invasion of cancer cells^[30]^, and adherens junctions mediate adhesive forces between adjacent cells^[31]^, initiating key intracellular signals that regulate major cellular functions, including proliferation, differentiation, and migration^[32]^. Interestingly, the pathogenesis of HAE may be related to the disruption of Kupffer cells^[33]^and metabolic pathways^[34]^, indicating a significant association between adherens junctions and HAE.

To systematically analyze the relationships and functions of DEmRNAs involved in the ceRNA network in HAE, we constructed a PPI network. Through this analysis, we identified several genes with high comprehensive scores, such as YAP1, ZEB1, PAFAH1B1, TJP1 and YWHAZ, which are primarily enriched in biological processes related to the regulation of developmental processes. Simultaneously, we observed that YAP1 and YWHAZ have a higher number of nodes in the PPI network, which typically indicates that DEmRNAs with more nodes play a more central role within the network. Research has indicated that YAP1 is one of the most critical effectors of the Hippo pathway and interacts with other pro-oncogenic pathways, promoting cancer development in various ways, including the promotion of malignant phenotypes, expansion of cancer stem cells, and enhancement of cancer cell drug resistance^[35][36]^. Additionally, YWHAZ has been closely associated with the progression of and as a therapeutic target for various types of cancer, including hepatocellular carcinoma^[37]^, gastric cancer^[38]^, and pancreatic cancer^[39]^. All in all, these genes are highly relevant to the occurrence and progression of related cancers and merit further research to clarify their roles in HAE.

Compelling evidence has shown that lncRNAs can compete for binding sites on miRNAs, thereby regulating the expression of target mRNAs. Within the ceRNA network, we discovered that Lnc-PCBP1-AS1 plays a pivotal role, which may influence variable targets through 16 lncRNA-associated miRNAs. Interestingly, a number of miRNAs, including miR-363^[40]^, miR-125a-5p^[41]^, miR-193a^[42]^, miR-4465^[43]^, miR-4319^[44]^, miR-125b-5p^[45]^, miR-22-3p^[46]^, miR-107^[47]^, miR-17-5p^[48]^, miR-20b-5p^[49]^, and miR-24-3p^[50]^, have all been associated with the initiation and progression of hepatocellular carcinoma. Furthermore, miR-20b-5p can be sponged by the lncRNA WWOX-AS1, which upregulates WWOX and thereby inhibits the progression of hepatocellular carcinoma^[51]^. The lncRNA MALAT1 can also function as a ceRNA, regulating TXNIP by sequestering miR-20b-5p^[52]^. Consequently, the predicted miRNAs can be utilized to identify lncRNAs that play a significant role in gene regulation in HAE, thereby enhancing our comprehension of the ceRNA network.

Within the ceRNA network, PCBP1, miR-20b-5p, and CAPRIN2 exhibit a high degree of intramodular connectivity, suggesting that they may play a pivotal role in regulating gene expression and biological functions. So, they have been selected as candidate genes for further research. It has been reported that PCBP1-AS1 is closely associated with the proliferation and metastasis of hepatocellular carcinoma^[53]^. Additionally, the dysregulation of miR-20b-5p has been confirmed in hepatocellular carcinoma, where it negatively regulates CPEB3, affecting the progression of the disease^[54]^. Concurrently, our research has discovered that PCBP1-AS1 shares miR-20b-5p response elements with CAPRIN2 and regulates the expression of CAPRIN2 by spongeing miR-20b-5p. Also,Experiments have validated that CAPRIN2 is indeed a genuine target of miR-20b-5p.Besides, the target gene CAPRIN2 plays a crucial role in the Wnt signaling pathway^[55]^, abnormal Wnt signaling is the basis for a variety of human diseases^[56]^, and dysregulation of the Wnt signaling pathway has been proven to be associated with a range of cancers^[57]^. Moreover, numerous preclinical experiments have indicated that inhibiting Wnt signaling can prevent the growth and survival of cancer cells^[58]^. The aforementioned GO enrichment results indicate that DEmRNAs involved in the ceRNA network in HAE are primarily engaged in the regulation of the Wnt signaling pathway. This suggests that the PCBP1-AS1-miR-20b-5p/CAPRIN2 axis may play a key role in HAE through the Wnt signaling pathway. Concurrently, we have analyzed the target gene CAPRIN2 in relation to clinical characteristics of patients and found that overexpression of CAPRIN2 is significantly associated with the size of lesions and the extent of vascular invasion in the liver in HAE. This suggests that PCBP1-AS1 might control CAPRIN2 expression in HAE by binding miR-20b-5p, affecting the HAE start and spread. However, the specific mechanisms by which the PCBP1-AS1-miR-20b-5p/CAPRIN2 axis modulates the development of HAE require further experimental validation. Considering that PCBP1-AS1 is a hub lncRNA in the ceRNA network, it could potentially serve as a target for HAE diagnosis and treatment. Overall, our research enhances the understanding of the regulatory mechanisms of the PCBP1-AS1-miR-20b-5p/CAPRIN2 axis in HAE, which may provide valuable insights to advance HAE research.

In conclusion, we have constructed a ceRNA network using lncRNA-miRNA and miRNA-mRNA interactions in HAE, laying the groundwork for further investigation into the regulatory mechanisms of HAE. However, this study still has some limitations, such as a lack of extensive experiments and a small sample size. Subsequently, we will expand our collection of clinical data based on these findings and conduct a more detailed analysis of the pathogenesis of HAE to clarify the roles of these differentially expressed genes within the ceRNA regulatory network. Equally, we will also focus on how the lnc-PCBP1-AS1-miR-20b-5p/CAPRIN2 axis regulates the progression of HAE, its functional and mechanistic aspects, which could lay the foundation for the diagnosis and treatment of HAE.

## Author contributions

All authors contributed to the article and approved the submitted version.

## Data availability

The experimental data has been deposited in the Gene Expression Omnibus (GEO) database under the accession number GSE261626.

