## Supplemental Data 1 for "Integrated analysis of high-throughput sequencing-based lncRNA-mediated ceRNA network in Hepatic Alveolar Echinococcosis"

Supplementary Table 1 All differentially expressed LncRNAs

| DELncRNAs | Fold Change | log2FoldChange | P-value |
| --- | --- | --- | --- |
| ASH1L-AS1 | 0.009262943 | -6.754313614 | 0.026910127 |
| CARMN | 472.4428004 | 8.88399586 | 0.000708197 |
| CASC15 | 23.94021746 | 4.581364352 | 0.011790669 |
| CCDC18-AS1 | 32.8647057 | 5.038467161 | 0.037844618 |
| CFAP58-DT | 56.45219971 | 5.818957893 | 0.049519927 |
| DHRS4-AS1 | 0.005395195 | -7.53410927 | 0.012038371 |
| DLEU2 | 0.003865589 | -8.015096043 | 0.006970118 |
| ELOA-AS1 | 0.02057074 | -5.603262518 | 0.043231617 |
| FAM198B-AS1 | 46.01981124 | 5.524183161 | 0.030562823 |
| FAM30A | 170.4393649 | 7.413114771 | 0.001305256 |
| FGD5-AS1 | 413.3040082 | 8.691059545 | 4.07E-05 |
| FTX | 33.62700911 | 5.071548562 | 0.006018204 |
| GAS5 | 0.013191302 | -6.244269202 | 0.049765996 |
| GCC2-AS1 | 0.003472513 | -8.169804204 | 0.035667132 |
| GMDS-DT | 205.2108265 | 7.680963036 | 0.004137238 |
| H19 | 0.018763189 | -5.735951158 | 0.02167632 |
| HCG27 | 18.81464559 | 4.233784208 | 0.049832111 |
| IFNG-AS1 | 16.74629173 | 4.065769758 | 0.036532403 |
| JPX | 0.018054225 | -5.791519704 | 0.028287157 |
| KCNQ1OT1 | 0.004968554 | -7.652958212 | 0.026930264 |
| LHFPL3-AS2 | 148.9802353 | 7.218977136 | 0.015313765 |
| LINC00115 | 99.59169256 | 6.6379535 | 0.004543642 |
| LINC00273 | 145.2239238 | 7.182135328 | 0.001101291 |
| LINC00342 | 70.98912691 | 6.149526165 | 0.043767268 |
| LINC00609 | 75.84174985 | 6.244920346 | 0.03022004 |
| LINC00847 | 165.8500055 | 7.37373525 | 0.004800032 |
| LINC00894 | 184.001548 | 7.523574093 | 0.011228308 |
| LINC00909 | 82.11017206 | 6.359489053 | 0.040104732 |
| LINC00927 | 0.008741662 | -6.837876743 | 0.027488408 |
| LINC00963 | 0.05012009 | -4.318467174 | 0.043996095 |
| LINC00997 | 0.003527496 | -8.147140011 | 0.037079123 |
| LINC01000 | 0.010762578 | -6.537832438 | 0.017896794 |
| LINC01050 | 20.62855828 | 4.366571091 | 0.015163635 |
| LINC01128 | 76.90994351 | 6.265098228 | 0.046080924 |
| LINC01138 | 137.2034488 | 7.100172936 | 0.012906901 |
| LINC01146 | 0.009040538 | -6.789375716 | 0.030343742 |
| LINC01181 | 68.98256866 | 6.108159945 | 0.006189199 |
| LINC01224 | 0.011377871 | -6.457625575 | 0.036055823 |
| LINC01278 | 28.41462431 | 4.828561735 | 0.039324056 |
| LINC01341 | 0.01070243 | -6.54591776 | 0.015665822 |
| LINC01348 | 0.027515571 | -5.183607921 | 0.028048602 |
| LINC01480 | 68.65328461 | 6.10125684 | 0.002337689 |
| LINC01481 | 0.011401728 | -6.45460367 | 0.0378121 |
| LINC01558 | 70.02981439 | 6.129897359 | 0.044940117 |
| LINC01614 | 59.9448277 | 5.905563372 | 0.00352363 |
| LINC01705 | 149.0362505 | 7.219519474 | 0.001476576 |
| LINC02015 | 0.002144737 | -8.864983325 | 0.003341887 |
| LINC02154 | 23.64339423 | 4.563365258 | 0.037611554 |
| LINC02331 | 74.88864714 | 6.226675122 | 0.04779759 |
| LINC02345 | 59.63206467 | 5.898016384 | 0.042922694 |
| LINC02362 | 214.2439295 | 7.743110517 | 0.00063924 |
| LINC02384 | 40.95935131 | 5.356120962 | 0.033312937 |
| LINC02470 | 480.5222832 | 8.908459524 | 0.000714181 |
| LINC02532 | 0.001066066 | -9.873487282 | 0.001419007 |
| LINC02577 | 228.8609033 | 7.838327215 | 0.008148109 |
| LINC-PINT | 26.17642243 | 4.71019603 | 0.010188852 |
| AACS | 52.64858757 | 5.718322923 | 0.008986333 |
| AAED1 | 49.01246626 | 5.615076839 | 0.013093875 |
| AAMP | 29.56285503 | 4.885713699 | 0.012616484 |
| ABCA5 | 21.61242525 | 4.433789069 | 0.029576639 |
| ABCB1 | 17.52477773 | 4.131324241 | 0.026337303 |
| ABHD6 | 34.58247215 | 5.111969099 | 0.008712893 |
| ABT1 | 0.017801081 | -5.811891346 | 0.042488374 |
| ACR | 25.9014382 | 4.694960302 | 0.042115383 |
| ACRBP | 40.72061269 | 5.347687364 | 0.01933934 |
| ACSBG1 | 70.80898709 | 6.145860574 | 0.010923969 |
| ACTR2 | 0.029082634 | -5.10369826 | 0.038661731 |
| ADAM30 | 0.016331455 | -5.936202856 | 0.034843909 |
| ADAMTS4 | 25.78572404 | 4.68850065 | 0.021682382 |
| ADH4 | 172.27627 | 7.428580183 | 0.00750105 |
| ADIPOQ | 18.42362852 | 4.203485322 | 0.017457738 |
| ADNP2 | 122.9374106 | 6.941780193 | 0.020125413 |
| AGAP5 | 12.50441866 | 3.644366082 | 0.031029719 |
| AGBL2 | 128.9924316 | 7.01114261 | 0.004503763 |
| AKAP6 | 100.8506007 | 6.656075867 | 0.009008852 |
| AKIRIN1 | 0.040123019 | -4.639426039 | 0.035642324 |
| AKR7A2 | 27.73156662 | 4.793457215 | 0.041210657 |
| ALG5 | 21.57715412 | 4.43143269 | 0.014187856 |
| ALOX12 | 0.004122964 | -7.922102377 | 0.008314352 |
| ANKRD12 | 13.57205622 | 3.762567406 | 0.027446567 |
| ANKRD36B | 258.1842466 | 8.012257166 | 0.003950314 |
| ANKRD44 | 47.4466717 | 5.568234983 | 0.002462619 |
| ANKRD52 | 33.05195634 | 5.046663762 | 0.028086864 |
| ANKRD55 | 6594.500459 | 12.68704766 | 7.57E-07 |
| ANXA2R | 98.45107209 | 6.621335011 | 0.007247527 |
| AP3S1 | 0.010663599 | -6.55116174 | 0.019665518 |
| APC | 24.73072886 | 4.628232854 | 0.034408432 |
| APIP | 112.3425496 | 6.81176064 | 0.026436838 |
| APOL5 | 15.02351277 | 3.909150276 | 0.027987132 |
| ARF6 | 67.15187681 | 6.069355817 | 0.048694268 |
| ARHGAP15 | 40.56512936 | 5.342168184 | 0.048213824 |
| ARHGAP17 | 18.82614103 | 4.234665402 | 0.047405536 |
| ARHGAP18 | 41.73023099 | 5.383021001 | 0.037659692 |
| ARHGAP44 | 79.14328338 | 6.306395014 | 0.035172491 |
| ARID1B | 28.73042858 | 4.844507609 | 0.016130217 |
| ARL10 | 34.99487011 | 5.129071548 | 0.044404712 |
| ARL15 | 20.69097107 | 4.37092945 | 0.04667361 |
| ARL5A | 0.022413488 | -5.479489038 | 0.048994236 |
| ARL5C | 28.31215197 | 4.823349506 | 0.02026322 |
| ARL6IP1 | 0.029459882 | -5.085104554 | 0.04338917 |
| ARX | 42.99267296 | 5.426018904 | 0.005102727 |
| ASB1 | 162.023091 | 7.340055625 | 0.018673152 |
| ATP5F1B | 367.1255637 | 8.520129765 | 0.001238116 |
| ATP6V1C1 | 72.46181127 | 6.179148963 | 0.032672224 |
| ATP6V1D | 181.2461695 | 7.501806695 | 0.014592424 |
| B3GALNT2 | 25.79358507 | 4.688940403 | 0.010175181 |
| B3GNT10 | 71.09760576 | 6.151729072 | 0.02573333 |
| BABAM2 | 20.04422008 | 4.325114378 | 0.014387876 |
| BACE1 | 0.006922838 | -7.174420751 | 0.019339471 |
| BAMBI | 32.06982899 | 5.003144756 | 0.013175376 |
| BAZ1A | 0.006153453 | -7.34438818 | 0.007183458 |
| BBS10 | 85.12055847 | 6.411435711 | 0.00123516 |
| BIN3 | 28.17916748 | 4.816557084 | 0.043295142 |
| BIRC2 | 15.0976768 | 3.916254662 | 0.028220711 |
| BMP6 | 148.9802353 | 7.218977136 | 0.015313765 |
| BMS1 | 22.39821471 | 4.485311839 | 0.020193027 |
| BPNT1 | 32.11499739 | 5.005175274 | 0.009037921 |
| BRAF | 194.7130914 | 7.605206076 | 0.004264427 |
| BRCA1 | 79.9865217 | 6.321685011 | 0.004199614 |
| BRF1 | 26.24675185 | 4.714066989 | 0.011130633 |
| BTD | 0.000349111 | -11.48402561 | 0.00185251 |
| BTG4 | 65.80579676 | 6.04014277 | 0.001820852 |
| C11orf94 | 89.21606491 | 6.479231611 | 0.027266562 |
| C16orf45 | 20.96899555 | 4.390185851 | 0.036043978 |
| C16orf46 | 14.86546733 | 3.893892913 | 0.040347228 |
| C17orf62 | 125.3998736 | 6.970392084 | 0.003970454 |
| C18orf32 | 12.34098656 | 3.625385826 | 0.031216184 |
| C1QTNF3 | 0.01138594 | -6.456602823 | 0.038920372 |
| C1QTNF9 | 91.35256475 | 6.513373328 | 0.001919606 |
| C1R | 52.12707412 | 5.703960979 | 0.038756291 |
| C2CD4B | 556.1516667 | 9.11933456 | 5.09E-05 |
| C2orf74 | 22.74591441 | 4.507535529 | 0.026822888 |
| C3orf85 | 0.002187306 | -8.836628933 | 0.034392456 |
| C5orf60 | 14.35842266 | 3.843825366 | 0.029785768 |
| C9 | 11.61601289 | 3.538043054 | 0.042710483 |
| CARHSP1 | 24.41741317 | 4.609838461 | 0.01193893 |
| CASTOR3 | 16.75312382 | 4.066358223 | 0.045229564 |
| CCDC14 | 39.25395968 | 5.294766286 | 0.010461292 |
| CCDC173 | 62.68567744 | 5.970063945 | 0.0046176 |
| CCDC34 | 0.002389105 | -8.709313742 | 0.04148242 |
| CCDC6 | 22.86724147 | 4.515210435 | 0.030015348 |
| CCDC71L | 0.025517949 | -5.292343834 | 0.022714155 |
| CCDC89 | 290.299189 | 8.181396731 | 0.000428083 |
| CCDC91 | 27.99432712 | 4.807062598 | 0.02693742 |
| CCNB1IP1 | 64.18539241 | 6.004173095 | 0.001266241 |
| CCNJ | 39.44901367 | 5.301917324 | 0.011404789 |
| CCNL1 | 18.32449791 | 4.195701764 | 0.011953356 |
| CD163 | 45.09886318 | 5.495019162 | 0.026911259 |
| CD200R1 | 39.42520339 | 5.301046293 | 0.01240122 |
| CDAN1 | 1023.422661 | 9.999186368 | 6.31E-05 |
| CENPK | 541.2747685 | 9.080217328 | 0.000578353 |
| CEP120 | 15.06610227 | 3.913234323 | 0.049002585 |
| CEP97 | 0.010675039 | -6.54961489 | 0.037264472 |
| CFAP36 | 0.01616138 | -5.951305773 | 0.03100405 |
| CFAP46 | 111.2192404 | 6.797262579 | 0.01551344 |
| CGAS | 26.14478159 | 4.708451113 | 0.022729045 |
| CHFR | 144.0901084 | 7.170827489 | 0.003322626 |
| CHRNA1 | 25.63826757 | 4.680226874 | 0.028801248 |
| CIB2 | 134.463639 | 7.071072289 | 0.00151225 |
| CLASP2 | 46.74982377 | 5.546889021 | 0.019597112 |
| CLEC18B | 0.008751136 | -6.836314011 | 0.02484231 |
| CLEC2D | 71.26964867 | 6.155215907 | 0.012341184 |
| CLEC3B | 16.481892 | 4.042809957 | 0.02467838 |
| CLHC1 | 11.14140494 | 3.477859264 | 0.033089015 |
| CLK1 | 1884.50725 | 10.87997163 | 3.65E-06 |
| CLNS1A | 37.53310257 | 5.230091647 | 0.025208437 |
| CMAS | 95.09953349 | 6.571366359 | 0.000794398 |
| CNGB1 | 32.61662588 | 5.027535641 | 0.033665759 |
| CNNM1 | 178.4501726 | 7.479377487 | 0.00093377 |
| CNR2 | 20.05429248 | 4.325839163 | 0.037973675 |
| CNTNAP3B | 156.0093269 | 7.285488472 | 0.002650201 |
| COA5 | 48.15634586 | 5.589654018 | 0.005723477 |
| COG7 | 14.81212517 | 3.888706741 | 0.023664089 |
| COL1A1 | 272.6642759 | 8.090981883 | 5.99E-05 |
| COL21A1 | 0.009225107 | -6.760218593 | 0.02585502 |
| COL24A1 | 21.63455552 | 4.435265576 | 0.028046397 |
| COQ5 | 63.14920261 | 5.980692612 | 0.026307312 |
| CPEB2 | 48.0458467 | 5.586339818 | 0.02433269 |
| CPLX1 | 0.050819937 | -4.298461606 | 0.046537736 |
| CPNE1 | 154.5822406 | 7.272230773 | 0.009409586 |
| CPNE4 | 17.8758394 | 4.159939084 | 0.02244194 |
| CPOX | 19.62346374 | 4.294507809 | 0.013742802 |
| CTNNBIP1 | 30.69646771 | 4.940000747 | 0.014996547 |
| CUL2 | 0.007297794 | -7.098323882 | 0.008739199 |
| CYP19A1 | 16.55745562 | 4.049409086 | 0.041115489 |
| CYP7A1 | 37.99792127 | 5.247848591 | 0.009667182 |
| DAPK2 | 0.017498206 | -5.836649176 | 0.03500126 |
| DCAF16 | 17.92287433 | 4.163730119 | 0.037771683 |
| DIRC3 | 34.08857137 | 5.091216233 | 0.021714213 |
| DKK4 | 0.000617285 | -10.66177625 | 0.00517931 |
| DLK1 | 47.05358441 | 5.556232723 | 0.003920838 |
| DLX2 | 59.38724729 | 5.892081257 | 0.008146853 |
| DNAH8 | 0.006822729 | -7.195435308 | 0.01694438 |
| DNM1L | 75.62343894 | 6.240761552 | 0.013583855 |
| DOK6 | 30.37193461 | 4.924666902 | 0.021073324 |
| DUOXA1 | 21.92656252 | 4.45460775 | 0.016096148 |
| DUSP2 | 24.76447517 | 4.630200142 | 0.016875326 |
| ECD | 105.0447216 | 6.714859859 | 0.018939413 |
| EEF1A1 | 9.716934719 | 3.280501277 | 0.041725916 |
| EFCAB11 | 47.06870875 | 5.556696371 | 0.01508863 |
| EFCAB8 | 24.21776621 | 4.597993895 | 0.019628025 |
| EGLN2 | 0.005176343 | -7.593851024 | 0.010653253 |
| EIF2AK3 | 23.50993511 | 4.555198652 | 0.016426551 |
| EIF2S3 | 37.55878193 | 5.231078371 | 0.006050832 |
| EIF3B | 1199.881175 | 10.22867583 | 4.26E-05 |
| EMC1 | 0.016528058 | -5.918938938 | 0.040068137 |
| EN1 | 36.57846745 | 5.192922726 | 0.016443038 |
| EPHX2 | 31.97608933 | 4.998921603 | 0.048647371 |
| ERI1 | 0.019268767 | -5.697591945 | 0.029057177 |
| ERMN | 219.9325156 | 7.780917103 | 0.001358035 |
| ERP27 | 16.17930755 | 4.016077959 | 0.029390881 |
| ERP44 | 0.010084469 | -6.631721133 | 0.03047405 |
| ERRFI1 | 0.005902736 | -7.404400523 | 0.009604916 |
| ESCO1 | 19.21309157 | 4.264017776 | 0.021206507 |
| EXOSC6 | 20.13669966 | 4.331755345 | 0.036689684 |
| F13A1 | 65.10855721 | 6.024775264 | 0.014154531 |
| FAM109A | 20.26267149 | 4.340752491 | 0.030563744 |
| FAM126B | 27.09785287 | 4.760106637 | 0.036471395 |
| FAM204A | 175.1496542 | 7.45244433 | 0.001476436 |
| FAM217A | 0.018803609 | -5.732846585 | 0.021710027 |
| FAM25C | 105.5224756 | 6.721406506 | 0.007622633 |
| FAM53B | 131.6411468 | 7.04046669 | 0.004263788 |
| FAM72B | 18.76650684 | 4.23008823 | 0.028322628 |
| FASTKD1 | 25.07632872 | 4.648254242 | 0.026092028 |
| FBLN1 | 0.036751829 | -4.766040147 | 0.04447245 |
| FBXO28 | 0.002819302 | -8.470446069 | 0.00589624 |
| FBXO43 | 23.31408556 | 4.543129939 | 0.016449684 |
| FCGR2B | 14.18997776 | 3.826800423 | 0.023942142 |
| FCN2 | 18.76570091 | 4.230026272 | 0.018312864 |
| FCRL5 | 30.59813048 | 4.935371603 | 0.018794781 |
| FGL2 | 52.75576067 | 5.721256733 | 0.004945643 |
| FNBP4 | 26.05524778 | 4.703502069 | 0.049813647 |
| FOXD4L6 | 34.72101865 | 5.117737369 | 0.00707523 |
| FOXO1 | 0.009966399 | -6.648711938 | 0.012540758 |
| FRG2C | 107.5333757 | 6.748640696 | 0.002257283 |
| FRMD4A | 0.011977026 | -6.383586503 | 0.040781472 |
| FTCDNL1 | 15.10065641 | 3.916539359 | 0.030477349 |
| FUBP1 | 55.90783508 | 5.804978575 | 0.009937052 |
| FXYD6 | 29.15570234 | 4.865706172 | 0.044501566 |
| FZD1 | 40.07233845 | 5.324534796 | 0.010578813 |
| GAB4 | 112.0466296 | 6.807955442 | 0.049722656 |
| GALNT15 | 195.1591741 | 7.608507473 | 0.002409716 |
| GCNT1 | 71.11978048 | 6.152178966 | 0.007685355 |
| GGCT | 13.22410781 | 3.725098487 | 0.029835971 |
| GIP | 81.39787511 | 6.346919228 | 0.01144404 |
| GJA10 | 16.6097416 | 4.053957724 | 0.035394086 |
| GJA9 | 107.3994747 | 6.746843126 | 0.035051005 |
| GNG4 | 150.4470249 | 7.233111768 | 0.001460296 |
| GOLGA6L22 | 18.99629967 | 4.247646515 | 0.015459863 |
| GOLGA8A | 25.45268267 | 4.669745817 | 0.012952172 |
| GOLGA8O | 12.57067123 | 3.651989781 | 0.048709997 |
| GPC1 | 91.50187607 | 6.515729418 | 0.009632858 |
| GPR108 | 23.17924686 | 4.534761786 | 0.041596102 |
| GPR141 | 36.61127401 | 5.194216073 | 0.027058907 |
| GPR149 | 0.000894799 | -10.12614818 | 0.008533875 |
| GPR155 | 17.01543724 | 4.088772319 | 0.023126228 |
| GPR180 | 0.020563527 | -5.603768433 | 0.022611949 |
| GPR75 | 0.007837165 | -6.995452451 | 0.012984911 |
| GRAP | 15.50233907 | 3.954414008 | 0.017164459 |
| GRHPR | 10.88941001 | 3.444853885 | 0.049456776 |
| GRTP1 | 13.83628389 | 3.790384615 | 0.04850447 |
| GSDMC | 23.09845315 | 4.529724336 | 0.037035969 |
| GSDME | 37.31797677 | 5.221798866 | 0.014329653 |
| GSN | 0.016346291 | -5.934892843 | 0.025664695 |
| GSPT1 | 17.34599752 | 4.116530903 | 0.023576028 |
| GTF2A2 | 82.52434177 | 6.366747821 | 0.001424684 |
| GTF2H2 | 32.10386112 | 5.004674915 | 0.007125852 |
| GXYLT1 | 31.40330271 | 4.972844391 | 0.04857527 |
| H6PD | 849.4247064 | 9.730342261 | 3.27E-05 |
| HACL1 | 12.07043149 | 3.593405345 | 0.047146019 |
| HDAC2 | 271.3084132 | 8.083789975 | 6.65E-05 |
| HDAC3 | 106.4644275 | 6.734227659 | 0.005738516 |
| HECTD4 | 163.193633 | 7.350440962 | 0.00041537 |
| HIST1H2AH | 23.80294205 | 4.573067997 | 0.031915977 |
| HK2 | 53.58666111 | 5.743802022 | 0.0373989 |
| HMCN1 | 38.05273019 | 5.249928062 | 0.034188541 |
| HMGB2 | 128.2224997 | 7.002505629 | 0.015708868 |
| HNRNPU | 20.71883912 | 4.372871265 | 0.046859887 |
| HS3ST3B1 | 0.016865975 | -5.889740511 | 0.032798517 |
| HS3ST5 | 49.12427402 | 5.618364182 | 0.042477866 |
| HSCB | 79.12643202 | 6.306087799 | 0.021413424 |
| ICAM2 | 965.9864436 | 9.915859133 | 1.33E-05 |
| ICE2 | 0.006894959 | -7.180242256 | 0.005010592 |
| IDS | 0.011925522 | -6.389803806 | 0.024429938 |
| IGF2 | 0.008906471 | -6.810930401 | 0.014650692 |
| IGFBP7 | 710.1803332 | 9.472041599 | 1.43E-05 |
| IGIP | 20.24220917 | 4.339294845 | 0.021111196 |
| IGLL1 | 15.49455217 | 3.953689153 | 0.034586567 |
| IL17B | 0.025050865 | -5.318995773 | 0.022543996 |
| IL5RA | 0.033575586 | -4.896443632 | 0.036233961 |
| INAFM2 | 14.09990479 | 3.817613515 | 0.039133981 |
| INTS14 | 22.1302676 | 4.467948991 | 0.014521362 |
| INTS5 | 97.91969195 | 6.613527114 | 0.002117932 |
| IRF2BP2 | 0.023734134 | -5.396892773 | 0.047308213 |
| ITGA9 | 0.002590394 | -8.592612539 | 0.044495761 |
| JAKMIP3 | 35.76160275 | 5.160339491 | 0.007003852 |
| JCAD | 26.83926224 | 4.74627311 | 0.037390051 |
| KALRN | 121.1234986 | 6.920334972 | 0.002775877 |
| KANSL1 | 0.013613616 | -6.198805894 | 0.031135824 |
| KANSL3 | 94.58081328 | 6.563475642 | 0.014362182 |
| KARS | 28.41963358 | 4.828816049 | 0.044122035 |
| KATNBL1 | 17.25515264 | 4.10895533 | 0.046453097 |
| KBTBD2 | 260.2517803 | 8.023764222 | 0.003860769 |
| KCNE1B | 281.1366865 | 8.135127918 | 0.000885348 |
| KCNE4 | 0.008652207 | -6.852716154 | 0.025778959 |
| KCNJ2 | 71.70832902 | 6.164068795 | 0.010437795 |
| KDM4C | 0.00849284 | -6.879537197 | 0.010332451 |
| KDM7A | 22.24192407 | 4.47520969 | 0.026198348 |
| KDSR | 532.1505921 | 9.055690758 | 6.33E-05 |
| KIAA0100 | 15.20830751 | 3.926787703 | 0.038227446 |
| KIAA0408 | 91.23158482 | 6.511461474 | 0.002146169 |
| KIAA0825 | 22.29055981 | 4.478360944 | 0.038571124 |
| KIAA1324L | 65.52657652 | 6.034008254 | 0.00969501 |
| KLF8 | 15.27967436 | 3.933541892 | 0.045235473 |
| KLHL1 | 33.53600732 | 5.067639032 | 0.027197686 |
| KNOP1 | 11.68085721 | 3.546074247 | 0.046655396 |
| KRT81 | 121.8326908 | 6.928757486 | 0.013468065 |
| LAMA4 | 0.002768475 | -8.496693025 | 0.00382411 |
| LAMB3 | 458.4624145 | 8.840659654 | 0.000133468 |
| LCP2 | 135.9559015 | 7.086994967 | 0.023073938 |
| LIMS4 | 115.5987718 | 6.852982259 | 0.02717793 |
| LINC02210 | 64.1412712 | 6.003181041 | 0.040718552 |
| LIPI | 33.42802633 | 5.062986272 | 0.039238726 |
| LITAF | 16.09889559 | 4.008889816 | 0.043396409 |
| LMOD3 | 21.54509044 | 4.429287249 | 0.049683952 |
| LPAR3 | 20.50530524 | 4.357925315 | 0.042991035 |
| LPCAT1 | 101.9989163 | 6.672410014 | 0.035494421 |
| LPP | 14.09995359 | 3.817618509 | 0.03553277 |
| LRP2BP | 29.87562695 | 4.900897084 | 0.023901966 |
| LRRC31 | 0.011488447 | -6.443672432 | 0.036253351 |
| LY75 | 46.73387108 | 5.546396639 | 0.020079526 |
| LYZL1 | 62.7745509 | 5.972107897 | 0.025128319 |
| MAFB | 60.04988868 | 5.908089666 | 0.033107947 |
| MAMDC2 | 0.026259726 | -5.251004327 | 0.043774998 |
| MAP3K5 | 0.009234283 | -6.758784261 | 0.007561384 |
| MBNL2 | 14.74463352 | 3.88211806 | 0.023252009 |
| MBTPS1 | 30.78044713 | 4.943942284 | 0.015959672 |
| MCL1 | 0.006931994 | -7.172513868 | 0.016727978 |
| MCOLN2 | 32.02370583 | 5.001068363 | 0.019099545 |
| MECR | 204.9250954 | 7.67895286 | 0.001903295 |
| MEIS1 | 23.62739358 | 4.562388585 | 0.010516439 |
| MESD | 21.10327729 | 4.399395159 | 0.04445015 |
| METTL14 | 0.016896574 | -5.887125419 | 0.023148203 |
| METTL2B | 10.92652912 | 3.449763288 | 0.045410778 |
| MFSD14B | 37.09577452 | 5.213182958 | 0.029516609 |
| MFSD8 | 28.09706609 | 4.812347586 | 0.007122898 |
| MICU2 | 23.56471074 | 4.558556067 | 0.030843517 |
| MIF4GD | 102.377646 | 6.67775693 | 0.005092392 |
| MIP | 0.003515004 | -8.152257874 | 0.037272047 |
| MITD1 | 68.03213918 | 6.088144548 | 0.003957455 |
| MLKL | 22.54312973 | 4.494615918 | 0.045648211 |
| MOB3A | 38.46432153 | 5.265448956 | 0.028152243 |
| MRPL20 | 17.72840142 | 4.147990549 | 0.023904561 |
| MRPS5 | 55.58569012 | 5.796641621 | 0.003677288 |
| MS4A6A | 72.7878379 | 6.185625506 | 0.005140183 |
| MTIF2 | 39.70947296 | 5.311411308 | 0.032299937 |
| MTMR2 | 27.58662565 | 4.785897094 | 0.017022737 |
| MTRF1 | 174.0163329 | 7.443078911 | 0.003026922 |
| MTRR | 80.90655467 | 6.338184683 | 0.011637936 |
| MUC20 | 0.010877428 | -6.522518673 | 0.037095303 |
| MYLK3 | 22.13818509 | 4.468465048 | 0.025444367 |
| MYOM1 | 0.003542171 | -8.141150361 | 0.00848774 |
| MZT1 | 558.2122509 | 9.124669977 | 0.000415114 |
| N4BP2L2 | 0.004143918 | -7.914788799 | 0.004363206 |
| NAA35 | 16.08792011 | 4.007905917 | 0.025889346 |
| NABP1 | 20.92811677 | 4.38737059 | 0.038534493 |
| NAXD | 117.3820635 | 6.875068165 | 0.000522027 |
| NBPF14 | 10.92146941 | 3.449095068 | 0.039313496 |
| NCOA4 | 0.007418984 | -7.0745627 | 0.022211735 |
| NDRG1 | 11.76340632 | 3.556233976 | 0.048466692 |
| NDUFA10 | 90.3613366 | 6.497633706 | 0.005765774 |
| NEDD9 | 64.27152646 | 6.006107832 | 0.023973203 |
| NEK10 | 19.99471892 | 4.321547095 | 0.027203669 |
| NEMF | 0.000869933 | -10.16680806 | 0.000977459 |
| NETO2 | 17.86095876 | 4.15873762 | 0.024868066 |
| NFKBIA | 70.02900839 | 6.129880754 | 0.003884162 |
| NFS1 | 23.25963237 | 4.539756389 | 0.013059693 |
| NIPAL2 | 13.18348344 | 3.720659716 | 0.04973244 |
| NMBR | 48.63663402 | 5.603971483 | 0.029985826 |
| NOL8 | 10.59577256 | 3.405416876 | 0.044067421 |
| NOTCH4 | 173.1559109 | 7.435927827 | 0.001495691 |
| NPBWR1 | 0.037826157 | -4.724471963 | 0.048148482 |
| NPC2 | 13.88975401 | 3.795949144 | 0.042115415 |
| NPIPB12 | 0.017939165 | -5.800743462 | 0.022961668 |
| NPIPB13 | 16.23489069 | 4.021025766 | 0.045251513 |
| NPIPB3 | 13.43002482 | 3.747390066 | 0.032962836 |
| NPIPB4 | 293.4787 | 8.19711199 | 0.000202266 |
| NPIPB5 | 15.41019171 | 3.945812904 | 0.033551033 |
| NT5C | 31.55762135 | 4.979916561 | 0.023206271 |
| NUPR2 | 126.8939317 | 6.987479268 | 0.00358533 |
| NXN | 60.86973114 | 5.927653088 | 0.026954588 |
| OLA1 | 139.0894313 | 7.119868991 | 0.005579982 |
| OR4F16 | 315.2693776 | 8.300441236 | 0.002268329 |
| OR4F29 | 24.84677264 | 4.634986566 | 0.009896069 |
| OR9A2 | 19.90490759 | 4.315052269 | 0.042596574 |
| OSGEPL1 | 10.37085813 | 3.374463369 | 0.048998312 |
| OTP | 338.3479203 | 8.40236371 | 0.00020377 |
| PANK3 | 14.66895246 | 3.874693944 | 0.020117181 |
| PATJ | 0.012176807 | -6.359720321 | 0.009887595 |
| PAWR | 74.2599094 | 6.21451165 | 0.004078109 |
| PAX1 | 0.031073413 | -5.008175457 | 0.03044054 |
| PAX8 | 12.10514803 | 3.597548817 | 0.049387433 |
| PCDH10 | 33.15343206 | 5.051086322 | 0.048783236 |
| PCGF2 | 10.87951266 | 3.443542028 | 0.037178276 |
| PCM1 | 0.006298002 | -7.310890046 | 0.016594951 |
| PDE1C | 72.25976385 | 6.175120635 | 0.005859809 |
| PGRMC2 | 44.71675812 | 5.482743694 | 0.022102157 |
| PHF14 | 0.011246184 | -6.474420685 | 0.017626615 |
| PJVK | 28.28124342 | 4.821773646 | 0.013376146 |
| PLA2R1 | 86.8902008 | 6.441121579 | 0.01241463 |
| PLEKHD1 | 28.69302311 | 4.842628074 | 0.010219062 |
| PLEKHO2 | 0.013160079 | -6.247688041 | 0.04325125 |
| POC1B | 0.009890807 | -6.659696047 | 0.031374231 |
| POTEB3 | 62.40139987 | 5.963506489 | 0.007972465 |
| POTEH | 26.34785089 | 4.719613386 | 0.020440155 |
| POU2F3 | 0.020713145 | -5.59330956 | 0.047708549 |
| PPEF1 | 0.017431582 | -5.842152644 | 0.033409455 |
| PPP1R9B | 101.0845603 | 6.659418846 | 0.000365558 |
| PREP | 84.15638804 | 6.395000881 | 0.042701362 |
| PRIM2 | 75.26283671 | 6.233865761 | 0.009255934 |
| PROB1 | 63.11389217 | 5.97988569 | 0.013435878 |
| PROM1 | 30.77376817 | 4.943629204 | 0.02092126 |
| PROP1 | 205.0530524 | 7.679853409 | 0.000916722 |
| PRPF18 | 45.91106395 | 5.520769961 | 0.028735615 |
| PRSS27 | 68.46290896 | 6.097250688 | 0.011688267 |
| PRSS55 | 44.20464629 | 5.466126112 | 0.027383476 |
| PRTFDC1 | 23.04874569 | 4.526616336 | 0.016015698 |
| PSIP1 | 82.11017206 | 6.359489053 | 0.040104732 |
| PSMA8 | 38.37297039 | 5.262018541 | 0.042617815 |
| PSMG1 | 20.03472506 | 4.324430806 | 0.017812343 |
| PTK2 | 0.002350077 | -8.733076393 | 0.039426589 |
| PXYLP1 | 18.9319169 | 4.24274859 | 0.033451291 |
| RAB17 | 15.33563173 | 3.938815692 | 0.043292467 |
| RAB23 | 0.013505132 | -6.210348499 | 0.022904371 |
| RAB35 | 49.40459508 | 5.626573327 | 0.013681256 |
| RAP1B | 47.98947249 | 5.58464605 | 0.007171549 |
| RAP1GAP | 43.85320406 | 5.454610349 | 0.03184052 |
| RBM15 | 62.01358429 | 5.954512372 | 0.008017546 |
| RBP4 | 26.7460221 | 4.741252432 | 0.033355275 |
| REP15 | 16.20477926 | 4.018347464 | 0.041326257 |
| RGPD8 | 40.27834713 | 5.331932577 | 0.004241489 |
| RGS10 | 31.72909952 | 4.987734673 | 0.010588546 |
| RGS11 | 15.86643752 | 3.987906332 | 0.027399665 |
| RHEX | 19.0250365 | 4.249827316 | 0.030151745 |
| RICTOR | 0.005874188 | -7.411394779 | 0.012478132 |
| RIOK2 | 14.500643 | 3.85804497 | 0.035861557 |
| RNASE4 | 18.05342291 | 4.174200492 | 0.03820079 |
| RNASEH1 | 40.55229641 | 5.341711709 | 0.029191405 |
| RPIA | 89.49516657 | 6.483737863 | 0.00123425 |
| RPL13 | 0.005447587 | -7.520166897 | 0.014137906 |
| RPL17 | 29.5571491 | 4.885435218 | 0.011176532 |
| RPLP0 | 12.13267243 | 3.600825458 | 0.033488066 |
| RPS6KA6 | 47.30717333 | 5.563987055 | 0.012716469 |
| RUBCN | 24.96424068 | 4.641791121 | 0.047458403 |
| RWDD2B | 14.65561508 | 3.873381613 | 0.041637115 |
| SAG | 12.1203909 | 3.599364324 | 0.045370093 |
| SCAPER | 0.003426074 | -8.189228116 | 0.036045273 |
| SCARB2 | 48.68511069 | 5.605408717 | 0.034664699 |
| SEC63 | 143.9135274 | 7.169058397 | 0.001661368 |
| SEM1 | 0.002202112 | -8.826896122 | 0.036038886 |
| SERF1A | 157.4798428 | 7.299023367 | 0.002581767 |
| SERPINB9 | 105.6881071 | 6.723669232 | 0.002793576 |
| SERPINC1 | 20.09069563 | 4.328455612 | 0.043884107 |
| SETD7 | 0.010377443 | -6.590405175 | 0.011264514 |
| SETD9 | 0.018105 | -5.787468035 | 0.037298843 |
| SEZ6L2 | 29.85163019 | 4.899737814 | 0.007824389 |
| SGMS1 | 0.029271769 | -5.094346272 | 0.021794817 |
| SH3BGRL2 | 24.27988959 | 4.601689956 | 0.04905945 |
| SH3BGRL | 90.89236062 | 6.506087138 | 0.027363212 |
| SH3BP2 | 92.68566348 | 6.534274297 | 0.001523747 |
| SHC4 | 21.89042638 | 4.45222815 | 0.041229288 |
| SIDT2 | 18.49798658 | 4.209296344 | 0.01999191 |
| SIKE1 | 27.91865266 | 4.803157415 | 0.040569438 |
| SKIL | 19.17718836 | 4.261319312 | 0.039010936 |
| SLC1A7 | 69.33865723 | 6.115587993 | 0.006642487 |
| SLC22A1 | 0.019416338 | -5.68658505 | 0.039702063 |
| SLC22A25 | 14.83280528 | 3.89071957 | 0.040852122 |
| SLC25A17 | 28.02162817 | 4.808468879 | 0.036754232 |
| SLC25A29 | 28.43035804 | 4.829360363 | 0.032602456 |
| SLC2A14 | 19.50956409 | 4.286109639 | 0.011819129 |
| SLC32A1 | 46.56920218 | 5.541304261 | 0.038681952 |
| SLC35E1 | 115.4839839 | 6.851548973 | 0.019197869 |
| SLC39A6 | 84.05051273 | 6.393184715 | 0.001791906 |
| SLC45A4 | 55.00096653 | 5.781385066 | 0.013340997 |
| SLC6A18 | 41.52464523 | 5.375895937 | 0.004336604 |
| SLX1B | 0.009535178 | -6.712524476 | 0.028609205 |
| SMIM14 | 21.79514245 | 4.445934728 | 0.02910454 |
| SNAPC5 | 89.98088596 | 6.491546667 | 0.002062313 |
| SNRPG | 87.69715341 | 6.454458109 | 0.001756749 |
| SNRPN | 19.2218842 | 4.264677856 | 0.011364287 |
| SNX31 | 21.60462069 | 4.433267997 | 0.016498675 |
| SOX4 | 27.3764534 | 4.774863654 | 0.008342258 |
| SOX5 | 0.009008412 | -6.79451154 | 0.007585294 |
| SP110 | 0.009870376 | -6.662679246 | 0.014155803 |
| SPATA32 | 20.9528548 | 4.389074918 | 0.035272416 |
| SPATA9 | 94.4264609 | 6.561119294 | 0.000757135 |
| SPIRE1 | 75.49846789 | 6.238375463 | 0.001890572 |
| SRPK2 | 0.016401999 | -5.929984512 | 0.012165201 |
| SRY | 116.8385691 | 6.868372785 | 0.0003142 |
| SS18 | 0.012937509 | -6.27229628 | 0.045827101 |
| SSBP2 | 62.74478789 | 5.971423717 | 0.035178716 |
| STYK1 | 29.51171621 | 4.883215916 | 0.015258242 |
| SUPT16H | 62.40546763 | 5.963600531 | 0.001201425 |
| SUPT3H | 99.15938838 | 6.631677468 | 0.026950427 |
| SYNJ2BP | 17.36735809 | 4.118306404 | 0.044241699 |
| SYT2 | 12.13524762 | 3.601131641 | 0.043931873 |
| TAL2 | 13.24066104 | 3.726903246 | 0.045942206 |
| TAS2R1 | 11.1290309 | 3.476256065 | 0.043501744 |
| TBC1D28 | 14.22771328 | 3.830631901 | 0.04947391 |
| TCIRG1 | 188.8660922 | 7.561219902 | 0.00049837 |
| TERB1 | 35.36822098 | 5.144381748 | 0.024395176 |
| TFAM | 14.63334359 | 3.871187545 | 0.020360375 |
| TFPI | 75.15288006 | 6.231756488 | 0.001337041 |
| TFPI2 | 29.44410487 | 4.87990691 | 0.01348415 |
| THOC5 | 60.70208 | 5.923674047 | 0.037974893 |
| THSD1 | 85.08779725 | 6.41088034 | 0.001561485 |
| TIGD2 | 0.009618967 | -6.699902353 | 0.029382243 |
| TIMM10B | 20.03888123 | 4.32473006 | 0.02732941 |
| TIMM8B | 17.87809707 | 4.160121281 | 0.043034964 |
| TKTL2 | 11.69067522 | 3.547286353 | 0.044635428 |
| TM4SF19 | 67.40058946 | 6.074689304 | 0.006050926 |
| TMEM135 | 45.16346831 | 5.497084377 | 0.017994582 |
| TMEM167A | 168.4582885 | 7.396247603 | 0.00790574 |
| TMEM178A | 26.37283893 | 4.720980975 | 0.015366782 |
| TMEM212 | 0.0026268 | -8.57247776 | 0.046062364 |
| TMEM30B | 50.78787931 | 5.666412329 | 0.002937306 |
| TMEM71 | 40.93648528 | 5.355315336 | 0.040866886 |
| TOB1 | 37.33950698 | 5.222630974 | 0.013533306 |
| TRABD2A | 148.3628861 | 7.212986428 | 0.008542875 |
| TRAP1 | 21.85294421 | 4.449755759 | 0.024678913 |
| TRIM69 | 0.023983164 | -5.381834203 | 0.030760539 |
| TRMT11 | 189.0702778 | 7.562778777 | 0.018704322 |
| TRPC4 | 119.969058 | 6.906518549 | 0.003004147 |
| TRPC4AP | 78.72308374 | 6.29871483 | 0.019947174 |
| TRPM1 | 89.53891603 | 6.484442948 | 0.03364729 |
| TSPAN8 | 12.5065585 | 3.644612945 | 0.041009993 |
| TTC30B | 0.026511291 | -5.237249249 | 0.036880628 |
| TXNDC12 | 47.5156121 | 5.570329709 | 0.045364278 |
| TXNDC16 | 25.91211167 | 4.695554685 | 0.048834855 |
| U2AF1 | 72.60836584 | 6.182063878 | 0.014923514 |
| UBASH3A | 54.23229599 | 5.761080345 | 0.005124605 |
| UBQLN2 | 20.63368468 | 4.36692957 | 0.040073767 |
| UBTD2 | 32.70000919 | 5.031219136 | 0.024150801 |
| UPK3A | 17.6199221 | 4.139135641 | 0.031195003 |
| URGCP | 0.00775037 | -7.011519184 | 0.019570823 |
| USP45 | 0.00416771 | -7.906529516 | 0.008977996 |
| USP54 | 22.85764361 | 4.514604779 | 0.028623489 |
| VEPH1 | 57.33056555 | 5.841232606 | 0.001950895 |
| VEZF1 | 98.58450914 | 6.623289065 | 0.006372393 |
| VSIR | 23.48632598 | 4.553749142 | 0.037754527 |
| VSTM5 | 33.69093465 | 5.074288547 | 0.007530758 |
| WDHD1 | 0.036415002 | -4.779323266 | 0.036162961 |
| WDR4 | 0.013233687 | -6.239641117 | 0.048733102 |
| WNT2 | 47.24821474 | 5.562187913 | 0.027940792 |
| XPO1 | 91.02630559 | 6.508211623 | 0.002239171 |
| XRCC2 | 0.011410362 | -6.453511653 | 0.012492771 |
| YME1L1 | 28.00483282 | 4.807603911 | 0.017841396 |
| ZBED5 | 35.63625754 | 5.15527393 | 0.007702635 |
| ZBTB20 | 14.55766669 | 3.863707233 | 0.027023947 |
| ZBTB2 | 59.29062275 | 5.889732045 | 0.006969771 |
| ZBTB7A | 56.80501323 | 5.827946353 | 0.024901466 |
| ZC3H11A | 681.4351841 | 9.412432629 | 2.95E-05 |
| ZC3H8 | 35.75381208 | 5.160025165 | 0.008860717 |
| ZDHHC20 | 55.12296256 | 5.784581522 | 0.003878912 |
| ZFHX4 | 106.1132794 | 6.729461401 | 0.023942274 |
| ZFP36L2 | 36.26628134 | 5.180556917 | 0.015449602 |
| ZFP37 | 30.00915411 | 4.907330748 | 0.040643739 |
| ZFP69 | 45.99808091 | 5.523501766 | 0.044896128 |
| ZMAT3 | 0.062398437 | -4.002346306 | 0.048697046 |
| ZNF106 | 82.11017206 | 6.359489053 | 0.007567313 |
| ZNF169 | 48.5477577 | 5.601332758 | 0.004416159 |
| ZNF207 | 0.007535731 | -7.052036835 | 0.021693085 |
| ZNF273 | 26.93602087 | 4.751464839 | 0.018650099 |
| ZNF280B | 24.04390446 | 4.587599287 | 0.007698445 |
| ZNF320 | 24.00044306 | 4.584989134 | 0.048503039 |
| ZNF33B | 16.12117773 | 4.010885238 | 0.024550608 |
| ZNF391 | 0.004930426 | -7.664072002 | 0.005604648 |
| ZNF460 | 0.004213009 | -7.890933361 | 0.01068163 |
| ZNF562 | 47.51118351 | 5.57019524 | 0.014127243 |
| ZNF583 | 97.98077715 | 6.61442683 | 0.013236246 |
| ZNF727 | 18.981246 | 4.246502794 | 0.041098154 |
| ZNF746 | 15.34900041 | 3.9400728 | 0.047993714 |
| ZNF843 | 0.018742973 | -5.737506357 | 0.042821746 |
| ZNF91 | 38.9404979 | 5.283199426 | 0.004996094 |
| ZRANB1 | 0.006117945 | -7.352737202 | 0.016589417 |
| ZSCAN10 | 336.4810029 | 8.394381245 | 0.004721667 |
| ZSCAN2 | 39.32058176 | 5.297212762 | 0.030063976 |
| ZSWIM2 | 75.75325551 | 6.243235985 | 0.046965216 |
| MAGI2-AS3 | 161.1645043 | 7.332390222 | 0.001823115 |
| MALAT1 | 302.7197609 | 8.241839044 | 6.37E-05 |
| MANCR | 59.72941334 | 5.900369647 | 0.047477959 |
| MIR100HG | 55.15213708 | 5.785344884 | 0.004947142 |
| MIR122HG | 0.012602334 | -6.310165219 | 0.030357554 |
| MIR17HG | 0.012078758 | -6.371384022 | 0.032803029 |
| MIR3142HG | 100.8589153 | 6.656194804 | 0.002917094 |
| MIR3936HG | 0.005265866 | -7.569113358 | 0.011897779 |
| MSC-AS1 | 135.2709351 | 7.079708079 | 0.004190024 |
| NDUFA6-DT | 0.019531215 | -5.678074476 | 0.048186972 |
| NEAT1 | 0.035194419 | -4.828509513 | 0.034955803 |
| NORAD | 793.7336708 | 9.632511197 | 9.98E-06 |
| NR2F2-AS1 | 0.020053647 | -5.639991561 | 0.020622963 |
| NUP50-DT | 0.003848259 | -8.021578401 | 0.006814156 |
| NUTM2A-AS1 | 0.00799501 | -6.966684374 | 0.01550682 |
| NUTM2B-AS1 | 0.019647978 | -5.66947535 | 0.013588716 |
| OIP5-AS1 | 15.68707349 | 3.97150433 | 0.025122667 |
| PCAT1 | 134.0249905 | 7.066358223 | 0.013629464 |
| PCBP1-AS1 | 34.83060525 | 5.122283637 | 0.036333413 |
| PCED1B-AS1 | 55.32794082 | 5.789936326 | 0.041951249 |
| PLCG1-AS1 | 0.006089681 | -7.359417528 | 0.013732985 |
| PSMA3-AS1 | 0.003840952 | -8.024320492 | 0.004218466 |
| PURPL | 30.12750477 | 4.913009285 | 0.03696591 |
| PVT1 | 128.2011462 | 7.00226535 | 0.017763083 |
| RAB11B-AS1 | 0.007037035 | -7.150816622 | 0.019605025 |
| RAB30-AS1 | 0.00706348 | -7.145405241 | 0.010818263 |
| RFX3-AS1 | 0.018295312 | -5.772382209 | 0.021486518 |
| RRS1-AS1 | 0.022212483 | -5.492485489 | 0.023942916 |
| RUNX2-AS1 | 241.7612975 | 7.917439498 | 0.00128587 |
| SGO1-AS1 | 44.57581003 | 5.47818911 | 0.038654722 |
| SLC16A1-AS1 | 44.47506879 | 5.474924932 | 0.049479796 |
| SLC25A25-AS1 | 80.37877493 | 6.328742684 | 0.008951449 |
| SNHG11 | 194.7252038 | 7.605295818 | 0.008117496 |
| SNHG12 | 707.2788953 | 9.466135403 | 0.000203128 |
| SNHG1 | 12.98281543 | 3.698531372 | 0.044704832 |
| SNHG21 | 49.26610324 | 5.622523459 | 0.041731026 |
| SNHG3 | 26.98446432 | 4.754057143 | 0.030060869 |
| SNHG5 | 15.14237941 | 3.920520017 | 0.019989772 |
| STK32A-AS1 | 51.02000265 | 5.672991069 | 0.043748503 |
| SYNJ2-IT1 | 31.13366101 | 4.960403328 | 0.034239653 |
| TCONS_00002059 | 0.004814205 | -7.698486662 | 0.004172812 |
| TCONS_00002476 | 0.019158945 | -5.705838107 | 0.016694594 |
| TCONS_00002655 | 0.009645435 | -6.695937926 | 0.013592265 |
| TCONS_00002658 | 0.003429474 | -8.187796975 | 0.005594436 |
| TCONS_00002960 | 0.009946611 | -6.651579258 | 0.011681117 |
| TCONS_00004551 | 0.00486982 | -7.681915864 | 0.005705262 |
| TCONS_00004581 | 0.010939918 | -6.514254311 | 0.035088028 |
| TCONS_00005379 | 0.034157407 | -4.871657741 | 0.043610834 |
| TCONS_00007400 | 366.7708817 | 8.518735296 | 2.74E-05 |
| TCONS_00008585 | 0.03516904 | -4.829550253 | 0.04567624 |
| TCONS_00009190 | 0.001019257 | -9.938266467 | 0.011016467 |
| TCONS_00009371 | 0.025079192 | -5.317365338 | 0.049104408 |
| TCONS_00010866 | 0.006124921 | -7.351092979 | 0.014029585 |
| TCONS_00012765 | 0.040280145 | -4.633787311 | 0.041231493 |
| TCONS_00013324 | 0.024887266 | -5.328448422 | 0.01806484 |
| TCONS_00014300 | 0.024638399 | -5.342947668 | 0.034962187 |
| TCONS_00016636 | 0.01092174 | -6.516653414 | 0.034489367 |
| TCONS_00018272 | 0.004812789 | -7.698911151 | 0.009234169 |
| TCONS_00018298 | 0.014199507 | -6.138015398 | 0.049713435 |
| TCONS_00018650 | 0.010683151 | -6.548518939 | 0.016293991 |
| TCONS_00019104 | 0.010226147 | -6.611593515 | 0.0335912 |
| TCONS_00019185 | 0.005683962 | -7.458887489 | 0.012487643 |
| TCONS_00020250 | 0.016353773 | -5.934232645 | 0.04309413 |
| TCONS_00020973 | 0.01142132 | -6.452126839 | 0.020191515 |
| TCONS_00024216 | 0.006932114 | -7.172488832 | 0.031547907 |
| TCONS_00024266 | 0.011991354 | -6.381861676 | 0.012726115 |
| TCONS_00025003 | 0.00537168 | -7.540411037 | 0.011167326 |
| TCONS_00025228 | 0.028986251 | -5.108487426 | 0.038529459 |
| TCONS_00026935 | 0.038418373 | -4.702059767 | 0.03575842 |
| TCONS_00027205 | 10.21182409 | 3.352168686 | 0.042610773 |
| TCONS_00027802 | 72.94768877 | 6.188790364 | 0.001998832 |
| TCONS_00028082 | 0.005032038 | -7.634641565 | 0.009829399 |
| TCONS_00028577 | 0.030212447 | -5.048713156 | 0.038810517 |
| TCONS_00029244 | 51.57303251 | 5.688544974 | 0.021832859 |
| TCONS_00029559 | 17.83167419 | 4.156370257 | 0.032345607 |
| TCONS_00030821 | 79.61470943 | 6.314963099 | 0.001827413 |
| TCONS_00030829 | 72.37308464 | 6.177381358 | 0.003358386 |
| TCONS_00032268 | 0.004184845 | -7.900610028 | 0.006339002 |
| TCONS_00032401 | 0.011344289 | -6.461890046 | 0.018026663 |
| TCONS_00032866 | 0.011151881 | -6.486569149 | 0.037795927 |
| TCONS_00037493 | 0.034642142 | -4.851328038 | 0.033086123 |
| TCONS_00039334 | 0.008931565 | -6.806871245 | 0.027199234 |
| TCONS_00039503 | 117.2714991 | 6.873708622 | 0.00671036 |
| TCONS_00039896 | 11.76831195 | 3.556835489 | 0.049523123 |
| TCONS_00040365 | 78.512234 | 6.294845571 | 0.02033009 |
| TCONS_00040697 | 0.00968659 | -6.689795417 | 0.015794439 |
| TCONS_00040945 | 0.007000528 | -7.158320621 | 0.015493862 |
| TCONS_00042045 | 0.024917173 | -5.326715783 | 0.03811343 |
| TCONS_00045290 | 0.03514392 | -4.830581069 | 0.035794754 |
| TCONS_00045571 | 35.70684194 | 5.158128636 | 0.019948212 |
| TCONS_00047676 | 77.53470618 | 6.276770331 | 0.000817871 |
| TCONS_00050054 | 0.01604949 | -5.961328733 | 0.013175898 |
| TCONS_00050487 | 0.010025263 | -6.640216056 | 0.029516182 |
| TCONS_00050644 | 123.5769338 | 6.949265672 | 0.032818803 |
| TCONS_00053156 | 0.045055265 | -4.472160489 | 0.044667597 |
| TCONS_00055069 | 0.002362323 | -8.725578037 | 0.039429992 |
| TCONS_00059700 | 91.64554688 | 6.517992876 | 0.03197599 |
| TCONS_00061007 | 0.019373173 | -5.68979592 | 0.031087662 |
| TCONS_00061318 | 0.010710564 | -6.544821699 | 0.032622951 |
| TCONS_00062785 | 32.45697059 | 5.020456445 | 0.022988474 |
| TCONS_00063300 | 0.008958223 | -6.802571714 | 0.01064927 |
| TCONS_00064544 | 0.004587744 | -7.767999437 | 0.008647371 |
| TCONS_00070539 | 0.029075605 | -5.104046957 | 0.035487605 |
| TCONS_00070986 | 0.002559409 | -8.60997369 | 0.003561094 |
| TCONS_00071174 | 0.020901527 | -5.580247846 | 0.044347501 |
| TCONS_00071847 | 12.87427783 | 3.686419603 | 0.036019929 |
| TCONS_00072673 | 0.041331127 | -4.5966275 | 0.034564134 |
| TCONS_00072674 | 0.000692128 | -10.49667275 | 0.000800034 |
| TCONS_00075283 | 0.006229416 | -7.326687282 | 0.013958524 |
| TCONS_00075315 | 0.009982717 | -6.646351761 | 0.029054318 |
| TCONS_00076237 | 0.023967756 | -5.38276137 | 0.025913829 |
| TCONS_00077200 | 0.01130629 | -6.466730578 | 0.038659594 |
| TCONS_00080998 | 23.98796077 | 4.584238614 | 0.037605275 |
| TCONS_00081827 | 75.60675613 | 6.240443253 | 0.002328838 |
| TCONS_00082380 | 17.203738 | 4.10465016 | 0.046674621 |
| TCONS_00084921 | 0.010539433 | -6.568058942 | 0.007927001 |
| TCONS_00086209 | 0.01045621 | -6.579496219 | 0.015712768 |
| TCONS_00086219 | 0.005477874 | -7.512168256 | 0.01330593 |
| TCONS_00086227 | 0.006897448 | -7.17972163 | 0.010464249 |
| TCONS_00086434 | 79.64027358 | 6.315426272 | 0.010526503 |
| TCONS_00087298 | 0.014284603 | -6.129395201 | 0.013352573 |
| TCONS_00088960 | 0.008964121 | -6.801622241 | 0.020989773 |
| TCONS_00088970 | 0.006049469 | -7.368975764 | 0.012707263 |
| TCONS_00093631 | 0.040840254 | -4.61386436 | 0.049082307 |
| TCONS_00093632 | 0.057022058 | -4.13233609 | 0.048617466 |
| TCONS_00093636 | 0.066781134 | -3.904415595 | 0.046408346 |
| TCONS_00094706 | 0.013916613 | -6.167048058 | 0.025811763 |
| TCONS_00098164 | 0.003354692 | -8.219604179 | 0.006125279 |
| TCONS_00098890 | 0.010645411 | -6.553624594 | 0.00774587 |
| TCONS_00099567 | 44.31461111 | 5.469710548 | 0.014222159 |
| TCONS_00100089 | 19.90244246 | 4.314873586 | 0.042218677 |
| TCONS_00101639 | 0.010497794 | -6.573770037 | 0.013698102 |
| TCONS_00102215 | 0.006114951 | -7.353443455 | 0.017462055 |
| TCONS_00103953 | 13.06450583 | 3.70758065 | 0.049949347 |
| TCONS_00108136 | 0.013344375 | -6.227624451 | 0.045952012 |
| TCONS_00108405 | 0.003253665 | -8.263718645 | 0.005146658 |
| TCONS_00108671 | 0.022234105 | -5.491081835 | 0.019332761 |
| TCONS_00109132 | 0.001853407 | -9.075604629 | 0.001522895 |
| TCONS_00109133 | 0.00398824 | -7.970032017 | 0.00676065 |
| TCONS_00109135 | 0.005941889 | -7.394862554 | 0.005374385 |
| TCONS_00109136 | 0.004022095 | -7.957837022 | 0.009026865 |
| TCONS_00109672 | 0.030628133 | -5.02899877 | 0.045750249 |
| TCONS_00110056 | 0.012101036 | -6.368725616 | 0.042049862 |
| TCONS_00110898 | 0.00624389 | -7.323339193 | 0.010784031 |
| TCONS_00111180 | 0.008946844 | -6.804405434 | 0.010204459 |
| TCONS_00112700 | 18.17117553 | 4.183579848 | 0.033420252 |
| TCONS_00113072 | 0.024134859 | -5.372737812 | 0.022079023 |
| TCONS_00113743 | 0.007878144 | -6.987928419 | 0.006834515 |
| TCONS_00113745 | 0.006557313 | -7.252679438 | 0.005991981 |
| TCONS_00115175 | 33.49845729 | 5.066022751 | 0.009249519 |
| TCONS_00116160 | 0.024295448 | -5.363170178 | 0.019198987 |
| TCONS_00116432 | 0.017150863 | -5.865575047 | 0.040742515 |
| TCONS_00116564 | 21.45227624 | 4.423058831 | 0.048675278 |
| TCONS_00116566 | 408.1227722 | 8.672859401 | 0.000626748 |
| TCONS_00117826 | 86.36047626 | 6.432299294 | 0.032050695 |
| TCONS_00117958 | 0.009409432 | -6.731676722 | 0.008568209 |
| TCONS_00119021 | 0.026969735 | -5.212514845 | 0.0418633 |
| TCONS_00119694 | 108.4937336 | 6.761467908 | 0.012936129 |
| TCONS_00120507 | 77.73225076 | 6.280441385 | 0.019366543 |
| TCONS_00120739 | 0.01310593 | -6.253636429 | 0.044681064 |
| TCONS_00120781 | 68.85566336 | 6.105503415 | 0.012360468 |
| TCONS_00120782 | 68.85566336 | 6.105503415 | 0.012360468 |
| TCONS_00122023 | 0.060633614 | -4.043738371 | 0.048352654 |
| TCONS_00124716 | 17.15668549 | 4.10069896 | 0.016941077 |
| TCONS_00125072 | 0.035412904 | -4.819581032 | 0.049376685 |
| TCONS_00127145 | 0.004886077 | -7.677107792 | 0.011161806 |
| TCONS_00127273 | 0.009627847 | -6.698571136 | 0.029362079 |
| TCONS_00127322 | 0.059587299 | -4.068851336 | 0.047783749 |
| TCONS_00127452 | 0.023243709 | -5.427015897 | 0.038270541 |
| TCONS_00128810 | 112.5923174 | 6.814964581 | 0.039529487 |
| TCONS_00130505 | 0.009018381 | -6.792915802 | 0.006495334 |
| TCONS_00130524 | 0.027073364 | -5.206982028 | 0.034265546 |
| TCONS_00130671 | 0.013097504 | -6.25456427 | 0.042956019 |
| TCONS_00130677 | 0.012147103 | -6.363243919 | 0.0102502 |
| TCONS_00130679 | 0.009008747 | -6.794457788 | 0.00925258 |
| TCONS_00132114 | 63.16154393 | 5.980974532 | 0.049769921 |
| TCONS_00133826 | 0.014934307 | -6.065225853 | 0.016209211 |
| TCONS_00134204 | 0.008314303 | -6.910188957 | 0.024410355 |
| TCONS_00137497 | 0.035908274 | -4.799539881 | 0.040820733 |
| TCONS_00139094 | 0.03304153 | -4.919575705 | 0.042237253 |
| TCONS_00139095 | 0.015872923 | -5.977288354 | 0.043344037 |
| TCONS_00141275 | 0.004771481 | -7.711347253 | 0.005369118 |
| TCONS_00143015 | 0.007460701 | -7.066473154 | 0.018302265 |
| TCONS_00143034 | 0.009716086 | -6.685409025 | 0.01726491 |
| TCONS_00143452 | 239.8869448 | 7.906210835 | 8.66E-05 |
| TCONS_00143625 | 0.034056438 | -4.875928637 | 0.036000109 |
| TCONS_00143696 | 770.7761313 | 9.590168086 | 8.96E-05 |
| TCONS_00144135 | 0.018975933 | -5.71968539 | 0.016653908 |
| TCONS_00146104 | 25.50976978 | 4.672977974 | 0.024739155 |
| TCONS_00147552 | 0.039628237 | -4.657327422 | 0.040673298 |
| TCONS_00148845 | 0.012908252 | -6.275562549 | 0.043761124 |
| TCONS_00151403 | 28.34472353 | 4.825008292 | 0.009974992 |
| TCONS_00151811 | 0.003065708 | -8.349564169 | 0.004615737 |
| TCONS_00153329 | 0.014441543 | -6.113631246 | 0.049553683 |
| TCONS_00153339 | 0.002757594 | -8.502373967 | 0.049890384 |
| TCONS_00154854 | 0.037350578 | -4.742725637 | 0.038776197 |
| TCONS_00156198 | 0.020258379 | -5.625337438 | 0.019439806 |
| TCONS_00156360 | 86.85101458 | 6.440470797 | 0.003366609 |
| TCONS_00156523 | 0.001466155 | -9.413747091 | 0.00135161 |
| TCONS_00157874 | 0.023097476 | -5.436120988 | 0.026394499 |
| TCONS_00158813 | 42.71940242 | 5.416819561 | 0.028561817 |
| TCONS_00161237 | 0.008291413 | -6.914166288 | 0.023833853 |
| TCONS_00161246 | 0.031032814 | -5.010061689 | 0.044968775 |
| TCONS_00162879 | 22.3704225 | 4.483520599 | 0.020358462 |
| TCONS_00163756 | 0.008501182 | -6.878120834 | 0.007008713 |
| TCONS_00164577 | 0.008980842 | -6.79893359 | 0.024247352 |
| TCONS_00165499 | 0.035739992 | -4.806316895 | 0.037956836 |
| TCONS_00166199 | 15.59053356 | 3.962598398 | 0.044518603 |
| TCONS_00167080 | 0.013031664 | -6.26183484 | 0.045581687 |
| TCONS_00167412 | 61.38629553 | 5.939844705 | 0.007250662 |
| TCONS_00168986 | 32.05204324 | 5.002344423 | 0.01279111 |
| TCONS_00170302 | 0.025525726 | -5.291904179 | 0.019220284 |
| TCONS_00170472 | 0.012504214 | -6.321441767 | 0.042288887 |
| TCONS_00172565 | 0.0126836 | -6.300891896 | 0.043426363 |
| TCONS_00172587 | 0.003112479 | -8.327720021 | 0.005300385 |
| TCONS_00174216 | 32.76652316 | 5.034150692 | 0.043444317 |
| TCONS_00175378 | 0.018321634 | -5.770307995 | 0.013485555 |
| TCONS_00175479 | 0.016149451 | -5.952371089 | 0.020423518 |
| TCONS_00176881 | 0.028587574 | -5.128468001 | 0.039967738 |
| TCONS_00177564 | 20.20781001 | 4.336841075 | 0.039494421 |
| TCONS_00178481 | 0.010140337 | -6.623750626 | 0.011657895 |
| TCONS_00178582 | 0.01227753 | -6.347835808 | 0.023583415 |
| TCONS_00179222 | 0.012067308 | -6.372752306 | 0.038936737 |
| TCONS_00179342 | 0.006567506 | -7.250438735 | 0.017397975 |
| TCONS_00179343 | 0.021744667 | -5.523194584 | 0.015913641 |
| TCONS_00180312 | 0.029164031 | -5.099666049 | 0.030524286 |
| TCONS_00184174 | 0.008780272 | -6.83151858 | 0.027014754 |
| TCONS_00184501 | 88.59347986 | 6.469128621 | 0.038479029 |
| TCONS_00185280 | 0.008193619 | -6.931283508 | 0.014345301 |
| TCONS_00185281 | 0.007356309 | -7.086802235 | 0.02227485 |
| TCONS_00189308 | 0.016282944 | -5.940494668 | 0.03108496 |
| TCONS_00189377 | 0.012863505 | -6.280572433 | 0.041812114 |
| TCONS_00189719 | 0.006535561 | -7.257473179 | 0.005405764 |
| TCONS_00190184 | 0.012274465 | -6.34819603 | 0.026028835 |
| TCONS_00190267 | 0.012202852 | -6.356637763 | 0.009069462 |
| TCONS_00190444 | 0.03082425 | -5.01979041 | 0.030095716 |
| TCONS_00191838 | 29.80115585 | 4.897296382 | 0.006031355 |
| TCONS_00192704 | 0.01186016 | -6.39773273 | 0.042650354 |
| TCONS_00193631 | 0.015633807 | -5.999187084 | 0.041767962 |
| TCONS_00193995 | 0.003841653 | -8.024057163 | 0.003179705 |
| TCONS_00195270 | 0.032091277 | -4.961674994 | 0.037104782 |
| TCONS_00197081 | 0.011083288 | -6.495470235 | 0.016751357 |
| TCONS_00197089 | 0.004519782 | -7.78953125 | 0.004903704 |
| TCONS_00198431 | 0.043058655 | -4.537552946 | 0.039823331 |
| TCONS_00201500 | 0.008432287 | -6.889860268 | 0.023334689 |
| TCONS_00203075 | 0.017601164 | -5.828185366 | 0.037729621 |
| TCONS_00205800 | 0.008458185 | -6.88543609 | 0.014278576 |
| TCONS_00205873 | 0.03933149 | -4.668171356 | 0.037601515 |
| TCONS_00205875 | 0.012934125 | -6.272673773 | 0.04076298 |
| TCONS_00206640 | 0.019385345 | -5.688889749 | 0.047716974 |
| TCONS_00208764 | 0.005705746 | -7.453368643 | 0.006035191 |
| TCONS_00208765 | 0.001865792 | -9.065995803 | 0.003205872 |
| TCONS_00209653 | 0.005646079 | -7.468535035 | 0.013119039 |
| TCONS_00210188 | 0.001440009 | -9.439706543 | 0.018531948 |
| TCONS_00210189 | 0.015021429 | -6.05683417 | 0.018775164 |
| TCONS_00210330 | 0.006547582 | -7.25482217 | 0.016063123 |
| TCONS_00210331 | 0.030401107 | -5.039732316 | 0.032260756 |
| TCONS_00211853 | 16.73318269 | 4.06463997 | 0.034899612 |
| TCONS_00212298 | 0.012902537 | -6.276201455 | 0.044920181 |
| TCONS_00216030 | 14.08862413 | 3.816458822 | 0.048895185 |
| TCONS_00217063 | 24.88595049 | 4.637259585 | 0.036244128 |
| TCONS_00218770 | 23.54917358 | 4.557604527 | 0.022912232 |
| TCONS_00219281 | 0.002758953 | -8.501663475 | 0.015064805 |
| TCONS_00219282 | 0.010160465 | -6.620889692 | 0.022920199 |
| TCONS_00219292 | 0.005378858 | -7.538484331 | 0.010361847 |
| TCONS_00219293 | 0.012997464 | -6.265626044 | 0.03253931 |
| TCONS_00219297 | 0.010089534 | -6.630996582 | 0.007866887 |
| TCONS_00219628 | 35.82607088 | 5.162937924 | 0.023210864 |
| TCONS_00220124 | 0.012061463 | -6.3734513 | 0.035021897 |
| TCONS_00222092 | 0.001251924 | -9.641637372 | 0.001642063 |
| TCONS_00222529 | 9.148286907 | 3.193501612 | 0.046446975 |
| TCONS_00223051 | 0.007039666 | -7.150277214 | 0.014599441 |
| TCONS_00224435 | 73.82170084 | 6.205973072 | 0.005105134 |
| TCONS_00225120 | 0.007455467 | -7.067485603 | 0.00708478 |
| TCONS_00225564 | 0.022199363 | -5.493337934 | 0.020920199 |
| TCONS_00225568 | 12.03916731 | 3.589663707 | 0.042046811 |
| TCONS_00228204 | 0.001822464 | -9.0998939 | 0.028036284 |
| TCONS_00228220 | 0.00646775 | -7.27252033 | 0.014717975 |
| TCONS_00229504 | 0.014270844 | -6.13078556 | 0.024751464 |
| TCONS_00231337 | 0.014429843 | -6.1148006 | 0.025230648 |
| TCONS_00231480 | 0.011324851 | -6.464364153 | 0.039190066 |
| TCONS_00232005 | 0.001314125 | -9.571681535 | 0.01667399 |
| TCONS_00232011 | 0.013418363 | -6.219647533 | 0.048074288 |
| TCONS_00232391 | 264.3779308 | 8.046457941 | 0.002601568 |
| TCONS_00236502 | 0.055579015 | -4.169315921 | 0.04388992 |
| TCONS_00237879 | 0.011545586 | -6.436514815 | 0.01655227 |
| TCONS_00238600 | 17.52877901 | 4.131653602 | 0.037802294 |
| TCONS_00239811 | 0.020470179 | -5.6103325 | 0.047499079 |
| TCONS_00241223 | 0.004368181 | -7.838751738 | 0.009629352 |
| TCONS_00241835 | 0.017215719 | -5.860129715 | 0.038334579 |
| TCONS_00242568 | 0.021333686 | -5.550722924 | 0.019269416 |
| TCONS_00246943 | 0.007146201 | -7.128607781 | 0.018194874 |
| TCONS_00247051 | 0.028756096 | -5.119988364 | 0.037711221 |
| TCONS_00247434 | 123.4707531 | 6.948025536 | 0.002563272 |
| TCONS_00248439 | 0.009038457 | -6.789707783 | 0.026366638 |
| TCONS_00249839 | 0.040874168 | -4.612666817 | 0.038203844 |
| TCONS_00250681 | 0.006646096 | -7.233277236 | 0.016725577 |
| TCONS_00252671 | 335.2849012 | 8.389243706 | 0.000111085 |
| TCONS_00253916 | 0.008281688 | -6.915859436 | 0.023365276 |
| TCONS_00254775 | 0.016681822 | -5.905579301 | 0.039725908 |
| TCONS_00256957 | 0.020181635 | -5.630813138 | 0.016457003 |
| TCONS_00256963 | 0.002827444 | -8.466285661 | 0.025050239 |
| TCONS_00260180 | 0.011763477 | -6.409541609 | 0.028151554 |
| TCONS_00260523 | 0.056002004 | -4.158377744 | 0.04389967 |
| TCONS_00262533 | 0.0401962 | -4.636797076 | 0.047281124 |
| TCONS_00263206 | 0.043298409 | -4.529542185 | 0.033019718 |
| TCONS_00263242 | 0.014302912 | -6.12754727 | 0.015292811 |
| TCONS_00267412 | 29.28722266 | 4.872199482 | 0.029297726 |
| TCONS_00268089 | 0.002029659 | -8.944546895 | 0.001952626 |
| TCONS_00268464 | 37.07350088 | 5.212316451 | 0.013687016 |
| TCONS_00268476 | 77.99692903 | 6.285345417 | 0.0045711 |
| TCONS_00268509 | 17.00230985 | 4.087658852 | 0.038125672 |
| TCONS_00270139 | 76.16686949 | 6.251091696 | 0.002283708 |
| TCONS_00271524 | 27.8191681 | 4.798007373 | 0.013605602 |
| TCONS_00272306 | 0.008357996 | -6.902627149 | 0.021758905 |
| TCONS_00272653 | 0.009495934 | -6.718474415 | 0.031095875 |
| TCONS_00274218 | 60.18072576 | 5.9112296 | 0.027622199 |
| TCONS_00274275 | 0.020539597 | -5.605448313 | 0.038164405 |
| TCONS_00274276 | 0.011227764 | -6.476785491 | 0.017280826 |
| TCONS_00274287 | 48.35885153 | 5.595708078 | 0.035993177 |
| TCONS_00276445 | 0.03954866 | -4.660227388 | 0.030643962 |
| TCONS_00276447 | 0.009567046 | -6.707710793 | 0.009981689 |
| TCONS_00278273 | 20.2419644 | 4.339277399 | 0.034089038 |
| TCONS_00278525 | 0.013077921 | -6.256723014 | 0.015579471 |
| TCONS_00279336 | 37.78833725 | 5.239869134 | 0.042662431 |
| TCONS_00280675 | 25.62982961 | 4.679751982 | 0.018640982 |
| TCONS_00280793 | 0.012972625 | -6.268385764 | 0.046920986 |
| TCONS_00281475 | 15.72796472 | 3.975260085 | 0.047831891 |
| TCONS_00282731 | 0.010630147 | -6.555694688 | 0.015947633 |
| TCONS_00283107 | 0.007395207 | -7.079193744 | 0.005915775 |
| TCONS_00283917 | 0.007041744 | -7.149851523 | 0.016914694 |
| TCONS_00284046 | 0.012346177 | -6.339791785 | 0.046058916 |
| TCONS_00284135 | 0.030916677 | -5.015470936 | 0.026090954 |
| TCONS_00284507 | 0.010807105 | -6.53187608 | 0.033538597 |
| TCONS_00284516 | 0.009850252 | -6.665623614 | 0.028462911 |
| TCONS_00284517 | 0.00616037 | -7.342767323 | 0.013768861 |
| TCONS_00285036 | 0.031547827 | -4.98631557 | 0.02090832 |
| TCONS_00285325 | 0.003501431 | -8.157839724 | 0.005999696 |
| TCONS_00287030 | 0.01839087 | -5.764866455 | 0.033720771 |
| TCONS_00287463 | 0.024368723 | -5.358825549 | 0.033062909 |
| TCONS_00289239 | 26.73814051 | 4.740827232 | 0.018113663 |
| TCONS_00289844 | 0.011411725 | -6.453339355 | 0.015550531 |
| TCONS_00298051 | 0.000904667 | -10.11032609 | 0.008877155 |
| TCONS_00298089 | 0.03066811 | -5.027116911 | 0.033823781 |
| TCONS_00298202 | 0.009551152 | -6.710109483 | 0.031012934 |
| TCONS_00299965 | 0.001438303 | -9.441416594 | 0.01822123 |
| TCONS_00306436 | 140.7219093 | 7.136703152 | 0.015330265 |
| TCONS_00306442 | 50.140095 | 5.647892825 | 0.014808576 |
| TCONS_00307062 | 0.002611095 | -8.581129107 | 0.046663318 |
| TCONS_00307798 | 0.015641092 | -5.998514974 | 0.013216434 |
| TCONS_00307799 | 0.004279059 | -7.86849086 | 0.003801398 |
| TCONS_00307803 | 0.006680299 | -7.225871653 | 0.007837889 |
| TCONS_00309473 | 0.018564821 | -5.751284778 | 0.024149989 |
| TCONS_00311838 | 0.038019569 | -4.717114003 | 0.037014692 |
| TCONS_00313407 | 0.008883796 | -6.814607997 | 0.025010005 |
| TCONS_00314218 | 0.017712327 | -5.819102421 | 0.024577177 |
| TCONS_00317126 | 0.019465368 | -5.682946604 | 0.027410234 |
| TCONS_00317132 | 0.023653329 | -5.401812916 | 0.025139089 |
| TCONS_00317134 | 0.00182636 | -9.096812919 | 0.026364715 |
| TCONS_00317784 | 0.005666261 | -7.463387222 | 0.017092152 |
| TCONS_00318939 | 1187.564098 | 10.21378967 | 1.36E-05 |
| TCONS_00319522 | 35.19087547 | 5.137129501 | 0.007128931 |
| TCONS_00319686 | 0.015472653 | -6.014135556 | 0.025649376 |
| TCONS_00320149 | 0.028594724 | -5.128107209 | 0.028549022 |
| TCONS_00320463 | 50.09216957 | 5.646513193 | 0.033066755 |
| TCONS_00320816 | 0.007715382 | -7.018046786 | 0.01898756 |
| TCONS_00324718 | 45.3658591 | 5.503535075 | 0.003006466 |
| TCONS_00329624 | 0.002778614 | -8.491419028 | 0.049229945 |
| TCONS_00331389 | 0.014848597 | -6.073529531 | 0.049309548 |
| TCONS_00332765 | 167.9285454 | 7.391703678 | 0.007967093 |
| TCONS_00333743 | 15.60093269 | 3.963560377 | 0.044391231 |
| TCONS_00334429 | 18.24306061 | 4.189275883 | 0.016682394 |
| TCONS_00334430 | 206.995094 | 7.693452765 | 9.85E-05 |
| TCONS_00334438 | 25.35056755 | 4.663946142 | 0.019146015 |
| TCONS_00335025 | 0.012137948 | -6.364331681 | 0.026795735 |
| TCONS_00335300 | 18.69070784 | 4.224249302 | 0.023424662 |
| TEX41 | 35.11809097 | 5.134142517 | 0.047858599 |
| THAP9-AS1 | 35.86665635 | 5.164571352 | 0.025736871 |
| TMEM161B-AS1 | 0.006584744 | -7.246656869 | 0.008150848 |
| TMEM9B-AS1 | 0.011859958 | -6.397757292 | 0.014709205 |
| TPT1-AS1 | 133.525854 | 7.060975302 | 0.006205731 |
| TTC28-AS1 | 56.87212302 | 5.829649755 | 0.02559824 |
| TUG1 | 18.43381188 | 4.204282528 | 0.021088319 |
| VIM-AS1 | 12.47106284 | 3.640512518 | 0.044384252 |
| WDFY3-AS2 | 150.7338167 | 7.235859308 | 0.012903081 |
| XIST | 324.3935406 | 8.341601282 | 0.000135074 |
| ZBED3-AS1 | 507.4528876 | 8.987130077 | 0.00078572 |
| ZEB1-AS1 | 139.322421 | 7.122283637 | 0.001822491 |
| ZFAS1 | 67.18056289 | 6.069971978 | 0.012519021 |
| ZFPM2-AS1 | 15.13972399 | 3.920266999 | 0.039066297 |
| ZMIZ1-AS1 | 25.74869924 | 4.686427648 | 0.039380614 |
