## Supplemental Data 2 for "Integrated analysis of high-throughput sequencing-based lncRNA-mediated ceRNA network in Hepatic Alveolar Echinococcosis"

Supplementary Table 2 All differentially expressed mRNAs

| DEmRNAs | Fold Change | log2FoldChange | P-value |
| --- | --- | --- | --- |
| VDR | 44.66824457 | 5.481177654 | 0.034277031 |
| XDH | 0.005278654 | -7.565614059 | 0.02672613 |
| LDLR | 0.001535247 | -9.347313565 | 0.018465857 |
| IL1A | 216.5918593 | 7.758835209 | 0.014583759 |
| SPP1 | 59.00144984 | 5.882678501 | 0.012287814 |
| CSF1 | 0.005595064 | -7.481629695 | 0.039247958 |
| DHFR | 0.006998859 | -7.158664465 | 0.035618003 |
| CD44 | 159.0717208 | 7.313533571 | 0.033905159 |
| EVI2A | 35.98043643 | 5.169140781 | 0.02928666 |
| AGMO | 0.003239164 | -8.270162874 | 0.023519917 |
| PRR13 | 30.08030948 | 4.910747505 | 0.03755105 |
| ALG3 | 31.02134913 | 4.955189526 | 0.032996849 |
| RPS4Y1 | 531.7809683 | 9.054688336 | 0.019309166 |
| SPIN4 | 110.1203779 | 6.782937655 | 0.006518374 |
| TFEC | 24.14362035 | 4.59357012 | 0.037874346 |
| NR3C1 | 42.73473354 | 5.417337222 | 0.019322765 |
| NRP1 | 20.66033595 | 4.368791809 | 0.044200846 |
| AMPD3 | 55.27819835 | 5.78863869 | 0.014054628 |
| MCUR1 | 0.006345177 | -7.300123947 | 0.040874679 |
| NR2C1 | 108.6101903 | 6.763015659 | 0.029545955 |
| ZNF226 | 105.695818 | 6.723774486 | 0.039549983 |
| TTPAL | 0.002662853 | -8.55281153 | 0.022054613 |
| THEMIS2 | 112.7078561 | 6.816444269 | 0.007544662 |
| GK5 | 0.006061455 | -7.366120114 | 0.041918652 |
| UBXN11 | 27.5675842 | 4.784900941 | 0.039865364 |
| RPL31 | 25.15856927 | 4.652977976 | 0.041670365 |
| TENM4 | 36.10635457 | 5.174180863 | 0.040088946 |
| MROH1 | 45.49000444 | 5.50747767 | 0.042892741 |
| TMEM192 | 0.006002822 | -7.380143391 | 0.046812004 |
| NBPF11 | 0.005341353 | -7.548579124 | 0.048244197 |
| CENPM | 46.07361946 | 5.525869033 | 0.019396788 |
| SLC8A1 | 335.3777413 | 8.389643132 | 0.009213824 |
| PABPC4L | 40.21973893 | 5.329831812 | 0.036521317 |
| MICAL3 | 81.43645404 | 6.347602839 | 0.021035691 |
| ACCS | 130.0328594 | 7.02273243 | 0.006879528 |
| ITGB2 | 24.52444382 | 4.616148514 | 0.027648878 |
| ZNF717 | 24.20700617 | 4.597352758 | 0.046018321 |
| MPV17L | 0.006620806 | -7.238777447 | 0.043669876 |
| CACNA1C | 156.7053397 | 7.29191053 | 0.01626969 |
| CSGALNACT1 | 96.1413614 | 6.587085328 | 0.030102746 |
| AHCYL2 | 169.0393978 | 7.401215723 | 0.017827798 |
| EPB41L2 | 420.0899308 | 8.714554396 | 0.022272468 |
| LRIG3 | 130.9907647 | 7.03332129 | 0.00582453 |
| TMEM51 | 25.50000816 | 4.672425804 | 0.038350495 |
| SGMS2 | 26.86787237 | 4.747810176 | 0.049051492 |
| DTHD1 | 61.9982461 | 5.954155498 | 0.025163916 |
| SPART | 429.6831405 | 8.747129361 | 0.0012714 |
| STRA6 | 32.60399856 | 5.026977003 | 0.046103252 |
| ANO6 | 24.9446389 | 4.64065788 | 0.043262411 |
| FKBP11 | 94.90877473 | 6.568469572 | 0.011683417 |
| SLC38A4 | 0.001245963 | -9.648522958 | 0.026619702 |
| FGFR2 | 139.5704122 | 7.124849324 | 0.036856658 |
| ADAMDEC1 | 37.66828918 | 5.235278605 | 0.020546049 |
| ZNF200 | 24.85454726 | 4.635437919 | 0.04361442 |
| TCP11L1 | 143.3623302 | 7.163522181 | 0.004316347 |
| ITGAM | 21.03720116 | 4.394870873 | 0.042733372 |
| NFASC | 68.91571861 | 6.106761172 | 0.030253097 |
| CSF2RA | 18.43458309 | 4.204342884 | 0.044188602 |
| TEX10 | 115.7794619 | 6.855235546 | 0.010805372 |
| SYTL2 | 58.30057935 | 5.865438315 | 0.01182417 |
| DCAF11 | 0.003965994 | -7.978101656 | 0.023582298 |
| GPC3 | 78.4114602 | 6.292992621 | 0.021028551 |
| PLCE1 | 49.06531682 | 5.616631671 | 0.027048963 |
| XIAP | 0.000481608 | -11.0198516 | 0.02239537 |
| XPNPEP1 | 57.18352321 | 5.837527605 | 0.036557802 |
| AFF2 | 95.3025582 | 6.574443036 | 0.0244613 |
| SLC38A6 | 41.4435113 | 5.373074335 | 0.0343788 |
| LDHB | 25.8985128 | 4.69479735 | 0.045377369 |
| SLC11A2 | 31.2844813 | 4.967375279 | 0.03275016 |
| SAMD8 | 0.008766174 | -6.833836917 | 0.045920542 |
| PPP2R2A | 66.04584271 | 6.045395848 | 0.015326853 |
| ZNF185 | 126.0392047 | 6.977728746 | 0.0395528 |
| NCK1 | 4520.924017 | 12.14240196 | 0.000119392 |
| TNFRSF17 | 31.91795389 | 4.996296265 | 0.021530859 |
| WLS | 83.56076718 | 6.384753833 | 0.035347535 |
| DDX3X | 0.002963459 | -8.398502152 | 0.024898285 |
| ATF5 | 0.007276721 | -7.102495781 | 0.037201649 |
| YAP1 | 22.06269959 | 4.463537424 | 0.037216999 |
| APTX | 514.6670237 | 9.007495537 | 0.002648994 |
| SSC5D | 64.66270539 | 6.014861964 | 0.041506551 |
| VAPB | 49.35108302 | 5.62500984 | 0.024737527 |
| RBBP7 | 25.93524303 | 4.696841984 | 0.028022849 |
| PI4KB | 635.3813664 | 9.311478971 | 0.025201653 |
| RBM14 | 18.57717258 | 4.215459037 | 0.037672841 |
| CYP4F3 | 0.000709682 | -10.46053966 | 0.024560557 |
| WASF2 | 59.63720352 | 5.898140704 | 0.032064486 |
| ZNF559 | 147.8907342 | 7.208387856 | 0.049540762 |
| CCN4 | 77.69596794 | 6.279767826 | 0.012676373 |
| TM4SF19 | 33.66725715 | 5.073274285 | 0.033614296 |
| SLC16A3 | 124.8899446 | 6.964513514 | 0.014995529 |
| MEIS2 | 71.12860257 | 6.152357915 | 0.016228686 |
| FAM156A | 114.0644594 | 6.833705532 | 0.017302705 |
| TREM1 | 75.29207298 | 6.234426076 | 0.019001495 |
| ZNF410 | 55.12232869 | 5.784564932 | 0.035364275 |
| EPDR1 | 27.71503274 | 4.792596807 | 0.044894607 |
| FYB1 | 24.34442468 | 4.605519501 | 0.041790871 |
| TAF1C | 39.22663621 | 5.29376172 | 0.026858634 |
| TCF4 | 33.23982029 | 5.054840677 | 0.025922356 |
| GFOD2 | 0.004059873 | -7.944349579 | 0.030839949 |
| ATPAF1 | 37.71600854 | 5.2371051 | 0.028714321 |
| DPYSL2 | 53.99983658 | 5.754883136 | 0.017605377 |
| RCAN3 | 160.9991757 | 7.330909492 | 0.037494934 |
| CREB3L2 | 22.85975512 | 4.514738044 | 0.029637228 |
| GMDS | 64.8179767 | 6.018322082 | 0.019200243 |
| TMCO1 | 236.8365739 | 7.887748078 | 0.040887673 |
| BIRC2 | 67.35406477 | 6.073693109 | 0.04147887 |
| RIT1 | 24.9253271 | 4.639540532 | 0.046139673 |
| DDX11 | 77.1684356 | 6.269938954 | 0.018352991 |
| SHTN1 | 0.009833002 | -6.66815241 | 0.045382449 |
| SLC4A7 | 867.9635356 | 9.761490624 | 0.020182377 |
| CLPB | 0.007382515 | -7.081671973 | 0.049636141 |
| BLNK | 1307.490692 | 10.35258496 | 0.014840142 |
| RPS3 | 64.11530608 | 6.002596904 | 0.008686245 |
| MGST1 | 0.001094733 | -9.835204775 | 0.041388896 |
| TMEM254 | 138.2053032 | 7.110669165 | 0.012378774 |
| TRAPPC3 | 136.465078 | 7.092387996 | 0.038684616 |
| PTGS1 | 92.78257035 | 6.535781909 | 0.00839597 |
| DICER1 | 29.3220227 | 4.873912722 | 0.040421844 |
| PLA2G2D | 51.11766247 | 5.67574996 | 0.04081724 |
| CDK5RAP2 | 38.07126422 | 5.250630573 | 0.023482049 |
| ELF2 | 159.5077035 | 7.317482291 | 0.024485048 |
| ZNF83 | 134.3693429 | 7.070060207 | 0.01743896 |
| LILRB4 | 74.06269244 | 6.210675092 | 0.018278236 |
| MDM4 | 0.00153438 | -9.348128419 | 0.015256378 |
| MAPK8 | 818.6577354 | 9.677116606 | 0.023545217 |
| SLC35C2 | 32.99374052 | 5.044120441 | 0.039269829 |
| H6PD | 0.010653198 | -6.552569559 | 0.036238648 |
| OLFM1 | 98.10602345 | 6.616269812 | 0.044639366 |
| MICAL2 | 222.7137668 | 7.799046929 | 0.013838016 |
| CRELD2 | 136.3512403 | 7.091184013 | 0.041426427 |
| VGLL4 | 43.04379083 | 5.427733235 | 0.030215169 |
| POSTN | 49.59421296 | 5.63209988 | 0.01151435 |
| AASDH | 58.56185071 | 5.871889242 | 0.019376393 |
| MLLT3 | 27.95250918 | 4.804905888 | 0.039311124 |
| ZNF79 | 470.0307861 | 8.876611443 | 0.004341327 |
| ANTXR2 | 78.87206206 | 6.301442456 | 0.012062122 |
| SH3GLB2 | 95.67725415 | 6.580104081 | 0.020626598 |
| CDC25B | 40.78806304 | 5.350075092 | 0.047144088 |
| CLIP4 | 31.19152414 | 4.963082145 | 0.023931186 |
| ZNF92 | 159.6627821 | 7.318884245 | 0.004869769 |
| METTL9 | 3199.851264 | 11.64378913 | 0.000214831 |
| DMPK | 209.3351547 | 7.709670801 | 0.005745025 |
| SLC39A7 | 346.1732962 | 8.435350629 | 0.037699999 |
| SLFN12 | 286.0445144 | 8.160095867 | 0.008177398 |
| HELLS | 414.0272747 | 8.693582 | 0.003173943 |
| PPP3CB | 24.7729639 | 4.630694582 | 0.045440753 |
| ZNF385A | 110.8580321 | 6.792569493 | 0.0241041 |
| CCL4L2 | 76.22925207 | 6.252272816 | 0.017889697 |
| DDX60L | 51.48910965 | 5.686195418 | 0.019458252 |
| REL | 0.007934529 | -6.977639752 | 0.03797467 |
| NOP14 | 0.00090851 | -10.10420936 | 0.037711312 |
| SEL1L3 | 22.3942203 | 4.485054532 | 0.043005584 |
| BTBD10 | 37.22626288 | 5.218248887 | 0.015697512 |
| CLEC1A | 53.80819239 | 5.749753937 | 0.027670259 |
| DTX4 | 25.81883531 | 4.690352017 | 0.042560989 |
| CLEC12A | 365.1855106 | 8.512485715 | 0.00323797 |
| MAGOHB | 61.08335392 | 5.932707374 | 0.017731161 |
| TBCB | 164.7995994 | 7.364568925 | 0.009080782 |
| CNOT8 | 285.5564532 | 8.157632178 | 0.042749786 |
| ERMN | 57.52143806 | 5.846027839 | 0.033111921 |
| SLMAP | 313.4678906 | 8.292173861 | 0.00283364 |
| FGD4 | 27.38106292 | 4.775106547 | 0.032709319 |
| SLBP | 142.9679899 | 7.159548358 | 0.006912464 |
| CCDC18 | 55.07047204 | 5.78320707 | 0.020197086 |
| CFAP54 | 60.7191391 | 5.924079431 | 0.012070274 |
| EGLN3 | 975.0436603 | 9.929323011 | 0.018052212 |
| ACSM2A | 0.001867484 | -9.064688193 | 0.036204316 |
| CPEB4 | 16.18378017 | 4.016476723 | 0.037656409 |
| RPS15 | 718.9112023 | 9.489669774 | 0.011553807 |
| HMGB1 | 0.001878776 | -9.055990929 | 0.0200914 |
| TLR2 | 70.020505 | 6.129705562 | 0.01410365 |
| CNPY3 | 104.6856673 | 6.709920124 | 0.022465432 |
| CARNMT1 | 33.48645781 | 5.065505871 | 0.030384535 |
| ADD3 | 17.19372581 | 4.1038103 | 0.045725921 |
| CEP70 | 115.792398 | 6.855396731 | 0.015927585 |
| GATM | 169.8809726 | 7.408380464 | 0.002622487 |
| RETREG2 | 31.60604526 | 4.982128623 | 0.028460282 |
| ARHGDIB | 180.2281988 | 7.493680945 | 0.006023792 |
| HDAC9 | 169.191763 | 7.402515523 | 0.011123357 |
| PNISR | 231.7445743 | 7.856391752 | 0.021065931 |
| CRLS1 | 24.80197237 | 4.63238295 | 0.045891436 |
| TASP1 | 71.81994486 | 6.16631264 | 0.03396716 |
| ZEB1 | 72.02922572 | 6.170510491 | 0.011997302 |
| GABPB2 | 164.6407662 | 7.363177791 | 0.007338216 |
| CTNNA1 | 20.02659293 | 4.323845095 | 0.048596148 |
| CMTR2 | 30.36792159 | 4.924476267 | 0.029276312 |
| PI15 | 76.25385269 | 6.252738326 | 0.04795491 |
| CELF2 | 57.0123574 | 5.833202752 | 0.018006927 |
| ERAP2 | 800.7993849 | 9.645297056 | 0.001459137 |
| CTBP2 | 43.0218671 | 5.426998232 | 0.045554741 |
| SPDL1 | 54.77863112 | 5.775541309 | 0.032796476 |
| AQP1 | 288.3827908 | 8.171841264 | 0.02171074 |
| WDSUB1 | 52.22346217 | 5.7066262 | 0.021189486 |
| SKA2 | 54.11414407 | 5.757933822 | 0.028053267 |
| ZNF131 | 34.97904139 | 5.128418847 | 0.037117394 |
| PPIP5K2 | 31.66007145 | 4.984592606 | 0.044226676 |
| USP28 | 112.7165973 | 6.816556156 | 0.01043437 |
| TFIP11 | 545.9751653 | 9.092691519 | 0.023423425 |
| PDGFRA | 13.80554372 | 3.787175803 | 0.047058211 |
| TRIP12 | 348.6502701 | 8.445636789 | 0.001970178 |
| SLC1A4 | 79.03988437 | 6.304508931 | 0.010991051 |
| KANSL3 | 42.75315103 | 5.41795885 | 0.046780104 |
| HPS4 | 183.1399405 | 7.516802649 | 0.006798584 |
| TOR1AIP2 | 0.003221808 | -8.277913863 | 0.025711931 |
| PBRM1 | 215.0199418 | 7.748326657 | 0.003320782 |
| ARHGEF2 | 47.60405978 | 5.57301271 | 0.020280198 |
| CAP1 | 137.4335098 | 7.102590002 | 0.008546291 |
| WDR60 | 24.3979747 | 4.608689488 | 0.041839164 |
| DGKA | 109.8784431 | 6.779764563 | 0.022259246 |
| IKZF4 | 68.44954384 | 6.096969022 | 0.043977484 |
| TMEM107 | 150.4583979 | 7.233220823 | 0.040343721 |
| TPCN1 | 29.93462784 | 4.903743429 | 0.034463653 |
| INPP4A | 45.64201644 | 5.512290625 | 0.018448344 |
| CFLAR | 62.21749376 | 5.959248376 | 0.006847791 |
| ZNF841 | 28.59566388 | 4.837724495 | 0.025618166 |
| ALG9 | 117.7870049 | 6.880036569 | 0.047541636 |
| PCM1 | 35.76437571 | 5.160451354 | 0.023862226 |
| TACC1 | 42.31494029 | 5.403095226 | 0.023819845 |
| ST18 | 96.03079013 | 6.585425143 | 0.011633948 |
| PLPP5 | 33.30379796 | 5.057614806 | 0.036489043 |
| NDUFAF6 | 187.6931163 | 7.55223193 | 0.047238075 |
| ZBTB25 | 394.2135866 | 8.622833689 | 0.006915578 |
| CDH6 | 31.58589743 | 4.981208658 | 0.035095358 |
| CTTNBP2 | 123.8482345 | 6.952429492 | 0.039606084 |
| EPHA4 | 64.05904975 | 6.001330492 | 0.022303518 |
| HNRNPH1 | 62.96542407 | 5.97648792 | 0.008952615 |
| TPM3 | 513.5743368 | 9.004429303 | 0.007879648 |
| AGRN | 31.13878931 | 4.960640948 | 0.039439992 |
| BAALC | 148.5047181 | 7.214364957 | 0.01716054 |
| CES2 | 0.001101447 | -9.826384333 | 0.009603027 |
| P4HA2 | 104.7693192 | 6.711072488 | 0.024784507 |
| ATP2B4 | 52.29121161 | 5.708496593 | 0.043680055 |
| CD200 | 339.4467791 | 8.407041586 | 0.006864473 |
| GLOD4 | 26.33400377 | 4.718854977 | 0.044138963 |
| KPNA5 | 47.52529325 | 5.570623624 | 0.047583004 |
| MECOM | 70.77979338 | 6.145265645 | 0.012665179 |
| SNX6 | 672.1373899 | 9.392612351 | 0.023196718 |
| SRRM1 | 328.3542873 | 8.359109482 | 0.001469205 |
| BORA | 70.8418628 | 6.146530243 | 0.019531026 |
| SUN1 | 61.81455501 | 5.949874674 | 0.015725367 |
| RABGEF1 | 96.90329623 | 6.598473836 | 0.024651029 |
| NEU3 | 117.5527438 | 6.877164403 | 0.024584309 |
| GOLGA8A | 22.98873183 | 4.522854977 | 0.031260559 |
| CBFB | 532.6500781 | 9.057044262 | 0.027101459 |
| AP1S2 | 113.9238366 | 6.831925828 | 0.006392826 |
| ALDH3A2 | 0.008247658 | -6.921799825 | 0.039711464 |
| ITGB1BP1 | 34.2521735 | 5.098123633 | 0.039944904 |
| CNTRL | 22.02704443 | 4.461204024 | 0.042109991 |
| ELF1 | 102.2818521 | 6.67640638 | 0.04053618 |
| ZNF266 | 408.2874443 | 8.673441392 | 0.003903458 |
| CUL1 | 66.42081516 | 6.053563524 | 0.022347858 |
| HK2 | 29.0977549 | 4.862835938 | 0.03721123 |
| APOC4 | 0.000922801 | -10.08169291 | 0.033656722 |
| CD1A | 32.16052437 | 5.007219025 | 0.043337392 |
| CPT1A | 0.004184953 | -7.900572875 | 0.028252952 |
| DBT | 0.004203853 | -7.894072012 | 0.021666798 |
| ACO1 | 0.006009647 | -7.378503921 | 0.038717441 |
| IVD | 0.007643723 | -7.031508728 | 0.034173413 |
| KCNN4 | 61.15285983 | 5.934348063 | 0.017987256 |
| MEF2C | 138.9082718 | 7.117988703 | 0.041122854 |
| MMP7 | 42.29506809 | 5.40241754 | 0.013191557 |
| MMP12 | 26.82929785 | 4.745737393 | 0.020737075 |
| P2RX1 | 56.3145195 | 5.815435033 | 0.018950426 |
| SLC25A3 | 0.004216099 | -7.889875541 | 0.024771261 |
| PLAU | 21.82270273 | 4.447757885 | 0.048092282 |
| SCN7A | 34.73545561 | 5.118337116 | 0.019881344 |
| CXCL5 | 18.75682405 | 4.229343663 | 0.045572194 |
| ST3GAL1 | 0.003588774 | -8.122293257 | 0.022199044 |
| TCEA3 | 0.002170395 | -8.847826877 | 0.0466993 |
| TLR1 | 25.02149305 | 4.645095973 | 0.046446699 |
| TPT1 | 0.001892148 | -9.045759259 | 0.015752727 |
| UBE2G2 | 0.010748294 | -6.539748549 | 0.043327074 |
| CXCR4 | 29.86376188 | 4.900324005 | 0.024374082 |
| CUL4B | 35.03579033 | 5.130757535 | 0.036179352 |
| ITGA10 | 76.24567836 | 6.252583662 | 0.017824827 |
| TMEFF1 | 31.55583263 | 4.979834786 | 0.044610519 |
| SPINT1 | 39.53741958 | 5.305146811 | 0.030855269 |
| PLPP2 | 96.23254368 | 6.588452958 | 0.043617876 |
| ABCB11 | 0.004017893 | -7.959345087 | 0.041162939 |
| B3GALNT1 | 35.97380652 | 5.168874919 | 0.046864668 |
| FUBP3 | 0.010964997 | -6.5109508 | 0.048209349 |
| HNF4G | 102.1646348 | 6.67475207 | 0.022096323 |
| PRKAR2A | 0.004503469 | -7.794747547 | 0.02991705 |
| BUB1 | 68.98365293 | 6.108182621 | 0.034408551 |
| CSNK1G3 | 33.3089802 | 5.057839279 | 0.030235831 |
| DFFA | 0.007028415 | -7.152584825 | 0.044935934 |
| PABPN1 | 52.3735752 | 5.710767186 | 0.020325041 |
| USP9Y | 2615.199052 | 11.35270504 | 0.007236588 |
| EIF1AY | 159.6386193 | 7.318665896 | 0.033027016 |
| MACROH2A1 | 42.94003125 | 5.424251336 | 0.024946727 |
| DOCK3 | 220.7315794 | 7.786149236 | 0.04653276 |
| OXA1L | 0.006541478 | -7.256167671 | 0.034753601 |
| BCAT1 | 19.02846777 | 4.250087491 | 0.049956043 |
| SMAD5 | 0.003002702 | -8.379522961 | 0.03438654 |
| ZBTB16 | 0.012984681 | -6.267045632 | 0.04829877 |
| B3GALT5 | 114.7803939 | 6.84273242 | 0.01658779 |
| SULT1C4 | 19.35407778 | 4.27456566 | 0.040328289 |
| MALT1 | 0.005614991 | -7.476500526 | 0.043582407 |
| TRPA1 | 28.6079604 | 4.83834474 | 0.042151414 |
| MAFF | 0.00692508 | -7.173953595 | 0.048667873 |
| ZNF112 | 56.3990246 | 5.817598307 | 0.02596052 |
| SPCS1 | 0.007802051 | -7.001930823 | 0.039203956 |
| MRPL42 | 0.012264846 | -6.349327082 | 0.045626408 |
| NUP58 | 0.00224953 | -8.796160993 | 0.022314116 |
| TRIM14 | 0.003660469 | -8.093755903 | 0.029606637 |
| MYH15 | 70.86224064 | 6.146945179 | 0.028083547 |
| ATMIN | 0.010505 | -6.572780046 | 0.043798761 |
| NSL1 | 0.007352277 | -7.087593118 | 0.046506006 |
| RSL1D1 | 0.005856609 | -7.415718593 | 0.037363962 |
| MEMO1 | 0.005322863 | -7.553581896 | 0.045938945 |
| ABHD5 | 0.005940299 | -7.395248747 | 0.042175879 |
| AK3 | 0.004564895 | -7.775202458 | 0.037969499 |
| PCYOX1 | 0.003043029 | -8.36027622 | 0.018166061 |
| C1RL | 0.005481264 | -7.511275672 | 0.025307109 |
| TM7SF3 | 0.010177088 | -6.618531408 | 0.039124873 |
| PGPEP1 | 0.004061324 | -7.943834162 | 0.038924876 |
| OCIAD1 | 0.001285323 | -9.603653067 | 0.043997668 |
| STX17 | 0.006417353 | -7.283806034 | 0.03410023 |
| ADI1 | 0.000842148 | -10.21363863 | 0.019701048 |
| FBXW7 | 29.95610635 | 4.904778212 | 0.049758733 |
| AGPAT5 | 0.005936842 | -7.396088588 | 0.049017759 |
| SYNJ2BP | 0.001428352 | -9.451432331 | 0.028118622 |
| DNAH7 | 149.4054626 | 7.223089087 | 0.013551582 |
| BRWD1 | 98.0624702 | 6.615629199 | 0.009862396 |
| NRIP3 | 21.36633816 | 4.41726777 | 0.037947121 |
| LRRN1 | 28.53896839 | 4.834861281 | 0.039320834 |
| GATAD1 | 0.004931749 | -7.663685045 | 0.043469747 |
| CCNB1IP1 | 0.003614346 | -8.112049513 | 0.027392717 |
| ADAM28 | 65.35096326 | 6.030136596 | 0.021046171 |
| SLC28A3 | 145.6025542 | 7.185891853 | 0.002953525 |
| ALDH8A1 | 0.006623981 | -7.238085787 | 0.039347238 |
| PRKAG2 | 30.2610193 | 4.919388678 | 0.045466496 |
| E2F8 | 51.62738931 | 5.690064741 | 0.034147445 |
| DNAJC22 | 0.002949808 | -8.405163196 | 0.023365184 |
| FBXO17 | 0.006986821 | -7.161148177 | 0.045132624 |
| ESRP2 | 0.002296126 | -8.766582346 | 0.049473002 |
| MED28 | 0.003176404 | -8.298389942 | 0.033822754 |
| BHLHE41 | 25.94254036 | 4.697247854 | 0.036623756 |
| RNF170 | 0.009743379 | -6.681362157 | 0.045381356 |
| TBXAS1 | 325.3881704 | 8.346017992 | 0.006456842 |
| ITCH | 0.00587718 | -7.410660265 | 0.031246985 |
| PHAX | 0.007007818 | -7.156818916 | 0.046480832 |
| PRXL2A | 0.003805338 | -8.0377598 | 0.034952191 |
| FCRLA | 170.756734 | 7.415798664 | 0.016875053 |
| SYAP1 | 0.001429811 | -9.449960054 | 0.016911442 |
| SPPL2A | 0.001595592 | -9.291692341 | 0.017513725 |
| EAF1 | 0.00925959 | -6.754835907 | 0.046790155 |
| PDGFD | 29.25056786 | 4.870392728 | 0.048021533 |
| SURF4 | 0.001096504 | -9.832873188 | 0.00970853 |
| N4BP2L1 | 0.005712219 | -7.451733112 | 0.037572315 |
| PTPRD | 366.4960743 | 8.517653935 | 0.034717387 |
| CCBE1 | 0.007600033 | -7.039778568 | 0.047008256 |
| KIAA1958 | 0.004561034 | -7.776423251 | 0.037654615 |
| PKD1L1 | 133.1962806 | 7.057409986 | 0.016999067 |
| CTHRC1 | 77.49043686 | 6.275946372 | 0.017785323 |
| PKHD1 | 34.8966601 | 5.12501706 | 0.022435813 |
| LIPH | 75.04276976 | 6.229641172 | 0.023942671 |
| WDR36 | 0.003073114 | -8.346082935 | 0.019419049 |
| PRAP1 | 0.007391406 | -7.079935405 | 0.035034844 |
| OSBPL3 | 141.9666796 | 7.149408551 | 0.003631297 |
| YWHAZ | 440.2155866 | 8.782066417 | 0.018065652 |
| SSBP3 | 0.006034173 | -7.372628233 | 0.047086596 |
| TRAF5 | 38.58937493 | 5.27013177 | 0.026028112 |
| ISL2 | 72.39969553 | 6.177911725 | 0.038730047 |
| EVC2 | 88.35198685 | 6.465190673 | 0.009945107 |
| TNFRSF19 | 41.47616914 | 5.374210744 | 0.042481711 |
| TNFAIP8L1 | 0.010256072 | -6.607377849 | 0.047547791 |
| ATP6V0D2 | 87.95628654 | 6.458714791 | 0.007859063 |
| CNKSR3 | 0.005476007 | -7.512659985 | 0.031632788 |
| ZNF610 | 47.40309854 | 5.56690946 | 0.03106361 |
| DTWD2 | 0.009491046 | -6.719217222 | 0.047800927 |
| TCAIM | 0.000926172 | -10.07643293 | 0.033614757 |
| SMIM14 | 0.0024343 | -8.682277335 | 0.021452942 |
| ATP2A3 | 33.40955592 | 5.062188901 | 0.028580033 |
| IL5RA | 51.67534704 | 5.691404267 | 0.024157871 |
| GATC | 0.006662653 | -7.229687607 | 0.049387457 |
| SULT1C2 | 18.60635322 | 4.217723414 | 0.048489958 |
| PPA2 | 0.005135454 | -7.605292535 | 0.036519157 |
| PPM1B | 0.001032366 | -9.919829426 | 0.036979737 |
| ATG4C | 40.64423923 | 5.344978979 | 0.023851172 |
| MTDH | 0.009385426 | -6.735362032 | 0.042680035 |
| LDLRAD4 | 70.41368948 | 6.137784033 | 0.041205937 |
| CCNY | 36.51253128 | 5.190319784 | 0.01546065 |
| LYRM7 | 0.000969555 | -10.01038995 | 0.035333348 |
| DPY19L4 | 0.005041503 | -7.631930326 | 0.04970896 |
| RBL1 | 31.45866236 | 4.975385423 | 0.028434394 |
| KIF1B | 0.000746086 | -10.38837045 | 0.024086477 |
| C15orf48 | 29.22372728 | 4.86906829 | 0.034083805 |
| DERL3 | 30.03097158 | 4.908379246 | 0.036605572 |
| IYD | 0.010928924 | -6.515704805 | 0.038850893 |
| CA12 | 59.86880524 | 5.903732575 | 0.016056944 |
| SYF2 | 424.3905124 | 8.729248594 | 0.006307315 |
| ARHGEF10L | 244.7608315 | 7.935228895 | 0.00305597 |
| CASP8 | 203.9208183 | 7.671865257 | 0.014326255 |
| ABCC5 | 57.83477622 | 5.853865345 | 0.019227613 |
| LPP | 0.000552562 | -10.82157673 | 0.028588536 |
| MBNL1 | 509.5509076 | 8.993082478 | 0.019502795 |
| TMEM45A | 24.17285571 | 4.595316014 | 0.047111038 |
| ZMAT3 | 0.003099071 | -8.333948556 | 0.035093766 |
| CD47 | 33.23605475 | 5.054677234 | 0.015979941 |
| P2RY14 | 29.27416364 | 4.871556049 | 0.047744215 |
| UGT3A1 | 0.001664969 | -9.230288807 | 0.040778339 |
| TMEM267 | 45.79549039 | 5.517133634 | 0.023262015 |
| GTF2H2C | 1561.441829 | 10.60866311 | 0.000267621 |
| ENPP5 | 44.81890118 | 5.486035374 | 0.02601653 |
| SSBP1 | 269.7278138 | 8.075360487 | 0.003454585 |
| TCAF1 | 0.003328692 | -8.230828684 | 0.030576631 |
| PTPN12 | 29.9988323 | 4.90683444 | 0.030451298 |
| DENND3 | 217.5255679 | 7.765041174 | 0.004049287 |
| AGTPBP1 | 364.1120089 | 8.508238513 | 0.002388188 |
| TRDMT1 | 434.4619014 | 8.76308586 | 0.004068984 |
| GATA3 | 68.09918584 | 6.089565645 | 0.037632399 |
| PFKP | 17.40711029 | 4.12160482 | 0.048326692 |
| SVIL | 109.4258134 | 6.773809298 | 0.044265477 |
| CCDC91 | 21.78240219 | 4.44509116 | 0.035755181 |
| RECQL | 28.70804321 | 4.843383092 | 0.033795645 |
| TCTN2 | 41.54366781 | 5.376556689 | 0.039649214 |
| ITGA11 | 77.79797702 | 6.281660736 | 0.017657181 |
| PKM | 455.1997643 | 8.830356 | 0.034157737 |
| UBE2Q2 | 132.5171593 | 7.050035372 | 0.0284618 |
| C17orf75 | 66.16836458 | 6.048069717 | 0.041927727 |
| CHST9 | 122.2479163 | 6.933666065 | 0.012112858 |
| ZNF383 | 50.49208092 | 5.657985231 | 0.040742649 |
| C20orf194 | 67.14440439 | 6.06919527 | 0.014805961 |
| NRIP1 | 0.00427808 | -7.868820967 | 0.041193701 |
| PDXK | 263.749294 | 8.043023422 | 0.00207232 |
| GGA1 | 43.34685897 | 5.437855551 | 0.027317469 |
| DOCK11 | 95.58496679 | 6.57871183 | 0.009180588 |
| GUCY1A1 | 78.84196065 | 6.300891749 | 0.008988707 |
| TSGA10 | 136.1417781 | 7.088966047 | 0.012170067 |
| CCDC88A | 89.63024852 | 6.485913792 | 0.008301575 |
| HNRNPLL | 228.2973644 | 7.834770394 | 0.002455562 |
| NGLY1 | 29.18135328 | 4.866974885 | 0.034553898 |
| RUFY3 | 218.3284733 | 7.770356482 | 0.003793065 |
| AKAP11 | 16.60928998 | 4.053918497 | 0.038014797 |
| CEP57L1 | 45.43079811 | 5.505598745 | 0.040601996 |
| ASPG | 0.013046391 | -6.26020541 | 0.04933325 |
| C5orf46 | 187.678758 | 7.552121561 | 0.005374005 |
| TBK1 | 16.35391917 | 4.03156451 | 0.040845203 |
| CTH | 0.003847914 | -8.021707576 | 0.030935402 |
| USP33 | 135.5079145 | 7.082233306 | 0.005211461 |
| PRKACB | 30.92567598 | 4.950733225 | 0.025291746 |
| AFF4 | 129.2166468 | 7.013648133 | 0.046043228 |
| NIPA2 | 374.3181463 | 8.548121179 | 0.004638131 |
| PQBP1 | 302.516233 | 8.240868749 | 0.009506554 |
| SLC7A2 | 0.000359791 | -11.44055405 | 0.017830314 |
| ASH2L | 33.81676978 | 5.079666953 | 0.040374956 |
| C11orf24 | 209.1579447 | 7.708448988 | 0.031204709 |
| PICALM | 69.25050045 | 6.113752592 | 0.01819885 |
| MTM1 | 28.47219245 | 4.831481684 | 0.04268329 |
| PECAM1 | 20.73079153 | 4.373703297 | 0.031946352 |
| ZYG11B | 0.001230058 | -9.667057454 | 0.042180283 |
| DENND2D | 173.9832826 | 7.442804879 | 0.009936967 |
| PRRX1 | 67.37398239 | 6.074119673 | 0.014715271 |
| MTHFD2 | 58.6000456 | 5.872829882 | 0.013418414 |
| AGFG1 | 369.9148809 | 8.531049528 | 0.034505942 |
| FAM193B | 32.00211194 | 5.000095212 | 0.047651755 |
| PPP1R11 | 40.81777069 | 5.351125485 | 0.041801227 |
| RIPK1 | 55.16899924 | 5.785785905 | 0.017946625 |
| AKAP7 | 306.0055087 | 8.257413814 | 0.04086715 |
| CACNA2D1 | 54.17543709 | 5.759566983 | 0.020870073 |
| ASAP1 | 111.4803776 | 6.800645984 | 0.010131221 |
| OXR1 | 57.49310951 | 5.845317156 | 0.042545147 |
| ZNF462 | 385.0480044 | 8.588894509 | 0.008194871 |
| PIP5K1B | 54.77059017 | 5.775329521 | 0.038046164 |
| PTBP3 | 0.006124277 | -7.351244643 | 0.035456404 |
| STAMBPL1 | 52.32965841 | 5.709556936 | 0.030542163 |
| DENND5B | 0.006137535 | -7.348125042 | 0.040231919 |
| CAPRIN2 | 57.9146308 | 5.855855953 | 0.037836237 |
| MLXIP | 207.4967119 | 7.696944665 | 0.004251354 |
| EXOSC8 | 68.74437463 | 6.103169757 | 0.029972411 |
| IFT88 | 101.1633152 | 6.660542412 | 0.029713311 |
| ZMYM5 | 117.0822938 | 6.871379105 | 0.010473824 |
| DOCK9 | 55.24084495 | 5.787663482 | 0.026429285 |
| KLHDC2 | 0.006065249 | -7.365217379 | 0.040803744 |
| AP4E1 | 40.75636233 | 5.348953386 | 0.0286211 |
| TANC2 | 368.0618833 | 8.523804541 | 0.030980192 |
| B3GNTL1 | 28.9106675 | 4.853530014 | 0.035820493 |
| ABCA7 | 119.5928217 | 6.901986987 | 0.017803869 |
| MAU2 | 26.6253786 | 4.734730135 | 0.045606847 |
| TPM4 | 32.00701286 | 5.000316135 | 0.025916208 |
| RPN2 | 770.9501726 | 9.59049381 | 0.006679532 |
| CHEK2 | 843.4259203 | 9.720117549 | 0.001355605 |
| ENTHD1 | 89.41245221 | 6.48240386 | 0.007787516 |
| TAZ | 31.05182724 | 4.956606261 | 0.03717214 |
| ADAMTSL4 | 58.07894187 | 5.859943264 | 0.025601698 |
| FCRL5 | 673.0462802 | 9.394561901 | 0.000778941 |
| PIP5K1A | 53.9982062 | 5.754839577 | 0.04260062 |
| KANSL1L | 55.25758343 | 5.788100565 | 0.020437949 |
| FAP | 206.3673315 | 7.689070796 | 0.016155512 |
| MGAT5 | 214.7566733 | 7.746559152 | 0.002726395 |
| NEB | 331.0920476 | 8.371088549 | 0.00608318 |
| PDK1 | 41.67452176 | 5.381093738 | 0.030885213 |
| COL6A6 | 635.8021638 | 9.312434116 | 0.001876481 |
| CDV3 | 39.50118546 | 5.303824045 | 0.021934133 |
| MFN1 | 288.0638917 | 8.170245022 | 0.024895939 |
| PHC3 | 82.32200778 | 6.363206264 | 0.01158901 |
| B3GNT5 | 52.00854706 | 5.700676829 | 0.029465337 |
| KLHL6 | 35.26312679 | 5.140088499 | 0.025381087 |
| ANAPC4 | 139.8480419 | 7.127716243 | 0.00784603 |
| RAI14 | 33.24325086 | 5.054989565 | 0.024177398 |
| CPLANE1 | 30.88748933 | 4.948950702 | 0.029132228 |
| TRIO | 175.8472209 | 7.458178725 | 0.004297805 |
| NUP155 | 30.73344523 | 4.941737597 | 0.020727798 |
| LRRC1 | 301.8839208 | 8.237850107 | 0.002964065 |
| TBC1D22B | 92.0312462 | 6.524051859 | 0.023984094 |
| DTNBP1 | 44.58327625 | 5.478430733 | 0.030941098 |
| INTS1 | 30.07816753 | 4.91064477 | 0.02562203 |
| NCAPG2 | 100.8423584 | 6.655957953 | 0.015216762 |
| FAM49B | 44.41564625 | 5.472996078 | 0.022559746 |
| RIC1 | 669.0063405 | 9.385876074 | 0.00352721 |
| SEMA4D | 102.5875145 | 6.680711346 | 0.012199265 |
| PNPLA7 | 83.79148652 | 6.388731764 | 0.028547024 |
| SHC3 | 159.3094171 | 7.31568774 | 0.023976777 |
| FSD1L | 181.5340341 | 7.504096241 | 0.007896485 |
| UAP1L1 | 107.2227439 | 6.74446715 | 0.018463511 |
| ZNF25 | 189.5218369 | 7.566220276 | 0.018874328 |
| DIP2C | 25.017698 | 4.644877141 | 0.046442537 |
| CUBN | 17.43190734 | 4.123658528 | 0.045937583 |
| ITIH5 | 220.4101392 | 7.784046781 | 0.036517058 |
| EXT2 | 30.47541089 | 4.929573767 | 0.033530752 |
| PHF21A | 121.4940386 | 6.924741716 | 0.020451427 |
| TRIM66 | 112.5857747 | 6.814880743 | 0.008771863 |
| PATL2 | 170.8338881 | 7.416450379 | 0.009687913 |
| MINAR1 | 141.1712406 | 7.141302403 | 0.012990111 |
| DMXL2 | 300.2797544 | 8.230163398 | 0.022362025 |
| ZNF710 | 35.30357724 | 5.141742471 | 0.026775247 |
| NPIPA5 | 27.85234585 | 4.799726938 | 0.038829101 |
| NLRC5 | 242.7352731 | 7.923239959 | 0.038206225 |
| PAFAH1B1 | 70.64756299 | 6.14256789 | 0.010044127 |
| NAT9 | 67.00174671 | 6.066126801 | 0.022849999 |
| CDK12 | 193.1623575 | 7.593670166 | 0.043393839 |
| CDC6 | 188.7316366 | 7.560192468 | 0.018643158 |
| SPIRE1 | 56.63367811 | 5.823588325 | 0.016112856 |
| NAPG | 43.67394439 | 5.448700928 | 0.021935019 |
| PIGN | 117.959071 | 6.882142555 | 0.013145452 |
| CCDC102B | 69.5244231 | 6.119447963 | 0.013049017 |
| ZNF565 | 61.22852477 | 5.936132018 | 0.031955664 |
| ZNF550 | 25.07715598 | 4.648301835 | 0.039912479 |
| SAMD4B | 103.5707044 | 6.694472176 | 0.034769881 |
| TGFB1 | 31.35919113 | 4.970816442 | 0.036140824 |
| ZNF229 | 53.38138881 | 5.738264935 | 0.034060566 |
| ZNF737 | 42.66457192 | 5.414966668 | 0.033254255 |
| CIRBP | 49.31707794 | 5.624015417 | 0.032380924 |
| MATK | 22.28571163 | 4.478047125 | 0.044861751 |
| MYO1F | 25.92102236 | 4.696050716 | 0.032926662 |
| ZBP1 | 65.40642975 | 6.031360561 | 0.024968159 |
| PRDM15 | 179.2738611 | 7.486021343 | 0.009428303 |
| TRPM2 | 32.64957822 | 5.028992449 | 0.037043605 |
| MEI1 | 60.194313 | 5.911555286 | 0.013190821 |
| SRRD | 40.45509129 | 5.338249373 | 0.017484411 |
| CELSR1 | 89.37580911 | 6.481812492 | 0.00727028 |
| MORC4 | 1114.003657 | 10.12153825 | 0.000947739 |
| PLXNA3 | 94.60588875 | 6.563858082 | 0.006971881 |
| CENPE | 85.62429227 | 6.419948253 | 0.030022717 |
| ARFIP1 | 2367.584213 | 11.20920003 | 0.00467192 |
| LARP1B | 358.5887942 | 8.486186595 | 0.036838182 |
| MAP9 | 26.45731245 | 4.725594614 | 0.03293924 |
| CAMK2D | 62.25682415 | 5.960160079 | 0.015669088 |
| CCNG2 | 21.41097687 | 4.420278715 | 0.037888349 |
| PUS10 | 62.6416401 | 5.969050081 | 0.028086986 |
| WDR35 | 57.71115602 | 5.850778325 | 0.0276121 |
| PDCD6IP | 50.01381311 | 5.644254697 | 0.046045072 |
| CHMP2B | 33.03984802 | 5.046135146 | 0.027979858 |
| APEH | 155.4748687 | 7.280537588 | 0.010759793 |
| ITPR1 | 31.76550555 | 4.989389075 | 0.031254756 |
| ZNF654 | 38.42437348 | 5.263949831 | 0.046459381 |
| HK3 | 75.13670621 | 6.231445968 | 0.009829348 |
| MCPH1 | 75.15556775 | 6.231808082 | 0.038219842 |
| HSPH1 | 89.23424004 | 6.479525487 | 0.03713667 |
| MYCBP2 | 30.44085139 | 4.927936804 | 0.028433723 |
| GABRR2 | 47.48135293 | 5.569289139 | 0.020636566 |
| FAM135A | 62.36350636 | 5.96263014 | 0.038752689 |
| RRAGD | 40.31328007 | 5.333183267 | 0.019204963 |
| PRDM1 | 22.06816479 | 4.463894753 | 0.028609952 |
| QKI | 219.8443033 | 7.780338339 | 0.02494354 |
| TDRD9 | 33.70014986 | 5.074683102 | 0.039219418 |
| CEP128 | 191.94934 | 7.584581789 | 0.033699811 |
| ZBTB1 | 30.81449823 | 4.945537394 | 0.030763455 |
| ACTR10 | 43.78856424 | 5.452482242 | 0.012666626 |
| METTL3 | 26.41292122 | 4.723171965 | 0.045985947 |
| PAPLN | 136.8495858 | 7.096447258 | 0.029290355 |
| MAPK1IP1L | 0.005269288 | -7.568176358 | 0.034013056 |
| LARP4 | 377.4362556 | 8.560089201 | 0.034051705 |
| ALDH1L2 | 61.42010987 | 5.940639188 | 0.015625472 |
| UHRF1BP1L | 144.3498037 | 7.173425335 | 0.003863135 |
| SFSWAP | 125.6710551 | 6.973508592 | 0.005974002 |
| ARHGAP9 | 27.31587981 | 4.771667986 | 0.029771823 |
| FMNL3 | 47.29568975 | 5.563636806 | 0.020557575 |
| SGMS1 | 111.3151892 | 6.798506655 | 0.006236244 |
| KCNMA1 | 39.34250805 | 5.298017026 | 0.044131362 |
| SHLD2 | 47.97059318 | 5.584078374 | 0.03348956 |
| CSGALNACT2 | 56.32175912 | 5.81562049 | 0.029375549 |
| ZFAND4 | 38.12213716 | 5.252557095 | 0.03703486 |
| WDR47 | 182.6042905 | 7.512576853 | 0.022092145 |
| MCOLN3 | 456.9741011 | 8.835968593 | 0.013824603 |
| CLSPN | 53.88289917 | 5.751755572 | 0.026213426 |
| CDC14A | 306.1538082 | 8.258112819 | 0.012312781 |
| EIF4G3 | 201.7005228 | 7.656071013 | 0.020604 |
| SPOCD1 | 52.57638567 | 5.716343063 | 0.039802064 |
| PHLDB1 | 25.57092819 | 4.676432624 | 0.049269392 |
| TRPC6 | 53.86181563 | 5.751190957 | 0.023244905 |
| ADAMTS6 | 219.4529588 | 7.777767912 | 0.025122875 |
| DMXL1 | 380.3340213 | 8.571123185 | 0.006878189 |
| FAM151B | 75.6842422 | 6.241921051 | 0.020910369 |
| FER | 55.30700312 | 5.789390265 | 0.014951218 |
| WDR41 | 14.99081156 | 3.906006584 | 0.048522977 |
| DEPDC1B | 73.42466546 | 6.198192882 | 0.035104334 |
| LOC102724971 | 1071.589534 | 10.06553668 | 0.011514911 |
| DNAH14 | 131.5750468 | 7.039742098 | 0.028713535 |
| SPRTN | 51.3648137 | 5.682708507 | 0.029560573 |
| CEP170 | 139.1569475 | 7.120569128 | 0.005287616 |
| EMSY | 98.61553727 | 6.623743062 | 0.0470788 |
| NHS | 66.05979027 | 6.045700484 | 0.022473143 |
| RPS6KA3 | 29.12263507 | 4.864068994 | 0.031003664 |
| ARHGAP17 | 91.89189891 | 6.521865776 | 0.015732316 |
| HNRNPR | 45.62189463 | 5.511654456 | 0.038621542 |
| CENPF | 20.46501912 | 4.35508811 | 0.041820588 |
| FHAD1 | 67.48354007 | 6.076463752 | 0.049179762 |
| SASS6 | 21.64858394 | 4.436200755 | 0.036451511 |
| EPS15 | 23.99663673 | 4.584760313 | 0.047751525 |
| POGZ | 99.65135034 | 6.63881745 | 0.041174426 |
| INPP5B | 163.7143762 | 7.355037204 | 0.012898767 |
| PPP1R12B | 0.005022254 | -7.637449169 | 0.038680062 |
| TMEM39B | 42.93007845 | 5.423916904 | 0.039551814 |
| PRPF38B | 128.6326007 | 7.007112516 | 0.047213933 |
| SCYL3 | 45.73848421 | 5.51533665 | 0.021986061 |
| MIER1 | 6891.139911 | 12.75052693 | 5.86E-05 |
| PTPRF | 25.52932536 | 4.674083508 | 0.026130521 |
| RAP1GAP | 229.0600464 | 7.83958203 | 0.047142264 |
| S100A6 | 85.62429227 | 6.419948253 | 0.015033423 |
| SFPQ | 0.004232941 | -7.884124053 | 0.026405177 |
| COP1 | 95.17525297 | 6.572514594 | 0.007128292 |
| PRPF3 | 51.2329306 | 5.678999513 | 0.021160292 |
| CD101 | 80.9711719 | 6.339336453 | 0.039402453 |
| RASAL2 | 24.40177395 | 4.608914127 | 0.048458371 |
| TANK | 0.001524069 | -9.357855918 | 0.048628354 |
| IMMT | 39.40130883 | 5.300171649 | 0.032551801 |
| COL6A3 | 27.99840356 | 4.807272663 | 0.034929545 |
| UBR3 | 121.761225 | 6.927910969 | 0.041200342 |
| CLASP1 | 87.33532156 | 6.448493345 | 0.0120986 |
| BAZ2B | 346.6669956 | 8.437406681 | 0.036009733 |
| GULP1 | 173.6274407 | 7.439851165 | 0.016870243 |
| REV1 | 344.3094467 | 8.427561954 | 0.031045422 |
| SERPINE2 | 60.42563218 | 5.917088757 | 0.014201265 |
| COPS7B | 92.12051528 | 6.525450576 | 0.019301603 |
| RGPD6 | 140.1179261 | 7.13049773 | 0.04375272 |
| VRK2 | 31.74966517 | 4.988669472 | 0.034218379 |
| RBM6 | 38.62984816 | 5.271644102 | 0.020303608 |
| USP19 | 39.25505375 | 5.294806495 | 0.035086299 |
| COL7A1 | 43.70729873 | 5.449802312 | 0.029517086 |
| DNAJC13 | 25.86295756 | 4.692815359 | 0.048851242 |
| NEPRO | 59.19634293 | 5.887436146 | 0.02120033 |
| DNAH1 | 21.00339822 | 4.392550861 | 0.037481785 |
| NKTR | 64.53458853 | 6.012000702 | 0.013935291 |
| ARHGEF3 | 51.21582925 | 5.678517867 | 0.042601768 |
| SETD5 | 141.630695 | 7.145990158 | 0.004413706 |
| CHDH | 0.004463182 | -7.807711627 | 0.037589284 |
| EAF2 | 429.4021169 | 8.746185493 | 0.031271784 |
| POGLUT1 | 114.103193 | 6.834195354 | 0.005965562 |
| SHOX2 | 69.34299622 | 6.11567827 | 0.038707843 |
| LRRC31 | 55.15217157 | 5.785345787 | 0.030287137 |
| B4GALT4 | 23.5347801 | 4.556722468 | 0.038837224 |
| IQCB1 | 66.04113667 | 6.045293046 | 0.013210541 |
| PSMD6 | 77.14504263 | 6.269501547 | 0.01171939 |
| SCLT1 | 19.34370429 | 4.27379219 | 0.040638883 |
| ZNF827 | 19.64049708 | 4.295759538 | 0.039575204 |
| RASGEF1B | 16.71185746 | 4.062800188 | 0.036761788 |
| SPATA5 | 45.62481545 | 5.511746818 | 0.039264113 |
| EXOC1 | 28.8691204 | 4.851455246 | 0.039270331 |
| PDGFC | 73.0592828 | 6.190995686 | 0.014992775 |
| TNIP1 | 199.8565955 | 7.642821374 | 0.04297922 |
| SUB1 | 1486.820226 | 10.5380145 | 0.000819456 |
| STARD4 | 66.87884181 | 6.063477958 | 0.029509886 |
| SLC38A9 | 62.54001239 | 5.9667076 | 0.033491034 |
| EBF1 | 187.3983239 | 7.549964239 | 0.008852379 |
| SSBP2 | 40.96683789 | 5.356384635 | 0.032983436 |
| TNFAIP8 | 124.590556 | 6.961050906 | 0.04772753 |
| UIMC1 | 322.9053891 | 8.334967708 | 0.028481674 |
| SLC22A5 | 35.55079552 | 5.151809938 | 0.035862388 |
| SMN1 | 161.9062133 | 7.339014542 | 0.044467269 |
| ELOVL7 | 30.22627641 | 4.917731358 | 0.035053847 |
| CARD6 | 29.56095939 | 4.885621187 | 0.048871287 |
| HMGN4 | 42.7792933 | 5.418840745 | 0.022263434 |
| TAF8 | 30.61275876 | 4.936061158 | 0.042086526 |
| UBR2 | 97.89059557 | 6.61309836 | 0.015467097 |
| SASH1 | 143.2483509 | 7.162374721 | 0.012891141 |
| SENP6 | 37.97479435 | 5.246970247 | 0.02360315 |
| SLC35B3 | 296.0949405 | 8.209916028 | 0.003858555 |
| REV3L | 137.2985197 | 7.101172261 | 0.002940291 |
| DST | 33.5406757 | 5.067839848 | 0.041793339 |
| PRPF4B | 38.14116837 | 5.253277133 | 0.023470583 |
| ARHGAP18 | 79.07243726 | 6.305102989 | 0.010219232 |
| FIG4 | 50.13209719 | 5.647662683 | 0.017984459 |
| RASA4B | 136.3730175 | 7.091414413 | 0.013628599 |
| SLC25A13 | 271.716173 | 8.08595663 | 0.001894586 |
| PEX1 | 35.01178735 | 5.129768808 | 0.027578885 |
| PIK3CG | 191.0468913 | 7.577782972 | 0.003375091 |
| GSAP | 38.54705902 | 5.268548888 | 0.043418402 |
| MTERF1 | 341.7163709 | 8.416655556 | 0.005668402 |
| LYPLA1 | 307.5996731 | 8.26491016 | 0.003066331 |
| EEF1D | 29.33720354 | 4.874659453 | 0.041857009 |
| PLAG1 | 36.46091803 | 5.188278982 | 0.042129358 |
| SLC20A2 | 59.63438938 | 5.898072625 | 0.047317416 |
| TAF2 | 165.9901086 | 7.374953463 | 0.0052426 |
| SLCO5A1 | 66.33666029 | 6.051734477 | 0.023716893 |
| TRIM55 | 180.8134604 | 7.498358271 | 0.037080776 |
| LRRCC1 | 180.3381737 | 7.494561006 | 0.004316411 |
| RAPGEF1 | 220.9007206 | 7.787254315 | 0.01651606 |
| NOL8 | 32.430943 | 5.019299069 | 0.034207571 |
| ZCCHC7 | 89.41713965 | 6.482479491 | 0.034681822 |
| MFSD14B | 43.45135553 | 5.44132928 | 0.029436371 |
| ATE1 | 272.499186 | 8.09010811 | 0.014555425 |
| ADK | 26.30442644 | 4.717233688 | 0.04511665 |
| MPP7 | 117.9146199 | 6.881598795 | 0.030965591 |
| ANKRD26 | 42.74128338 | 5.417558323 | 0.027569575 |
| IPMK | 1317.737516 | 10.36384731 | 0.000618708 |
| KIFBP | 32.53368042 | 5.023862134 | 0.035034155 |
| POLL | 39.87009614 | 5.317235179 | 0.028208293 |
| KCNIP2 | 125.5453789 | 6.972065116 | 0.028370929 |
| FAM149B1 | 38.44788929 | 5.264832494 | 0.029724954 |
| KIF5B | 48.20575475 | 5.591133479 | 0.047674315 |
| SRGN | 253.1516519 | 7.983858088 | 0.010370131 |
| FRMD4A | 25.81794012 | 4.690301995 | 0.04586426 |
| SFMBT2 | 25.43046835 | 4.668486127 | 0.037966361 |
| UROS | 71.93402216 | 6.168602369 | 0.014127405 |
| OR13A1 | 88.94402668 | 6.474825815 | 0.033334928 |
| TNKS2 | 31.35606744 | 4.970672728 | 0.026738585 |
| RASSF4 | 27.00769761 | 4.755298751 | 0.04356582 |
| FANK1 | 191.6440303 | 7.582285249 | 0.017832853 |
| JAML | 93.95268165 | 6.553862435 | 0.008006683 |
| ATM | 992.5095138 | 9.954937121 | 0.018663711 |
| POU2AF1 | 33.9468789 | 5.085207033 | 0.022996096 |
| IFT46 | 465.6172455 | 8.863000683 | 0.005157412 |
| DYNC2H1 | 159.4635606 | 7.317082977 | 0.042551297 |
| MSANTD2 | 0.006274262 | -7.316338581 | 0.041968588 |
| CASP1 | 65.30517648 | 6.029125448 | 0.023302018 |
| CASP4 | 172.3912579 | 7.429542805 | 0.018989955 |
| MADD | 224.5264767 | 7.810741771 | 0.016912798 |
| LPXN | 36.891447 | 5.205214472 | 0.047332759 |
| OSBPL8 | 174.5915905 | 7.44784026 | 0.004970347 |
| ERC1 | 20.56797732 | 4.36232802 | 0.03779088 |
| NEMP1 | 29.88810506 | 4.901499527 | 0.033052831 |
| FBXW8 | 182.8767602 | 7.51472794 | 0.010042254 |
| ANAPC7 | 94.54657984 | 6.562953365 | 0.010029452 |
| LIMA1 | 61.17193611 | 5.934798033 | 0.03743981 |
| FOXJ2 | 120.9285203 | 6.918010725 | 0.040533474 |
| DDX55 | 105.4641228 | 6.720608491 | 0.013097633 |
| SINHCAF | 56.53065079 | 5.8209614 | 0.024455329 |
| TBC1D15 | 39.5623727 | 5.306057047 | 0.021488652 |
| RERG | 191.4846575 | 7.581084992 | 0.006891538 |
| PCED1B | 32.58679506 | 5.026215563 | 0.049066911 |
| KNTC1 | 166.8979574 | 7.382822488 | 0.005786513 |
| TUBGCP3 | 532.1846474 | 9.055783082 | 0.00346192 |
| N4BP2L2 | 357.7769269 | 8.482916541 | 0.012552642 |
| PDS5B | 35.17359285 | 5.136420803 | 0.041921814 |
| RBM26 | 541.0479342 | 9.079612605 | 0.020227428 |
| TSC22D1 | 99.73836859 | 6.6400767 | 0.015976157 |
| ARHGEF7 | 171.107467 | 7.418758909 | 0.047935873 |
| ZFYVE26 | 211.5965843 | 7.725172529 | 0.017170142 |
| FUT8 | 29.18209575 | 4.867011591 | 0.031304511 |
| AKAP6 | 28.28709291 | 4.822072012 | 0.041742179 |
| TUBGCP5 | 231.7950918 | 7.856706207 | 0.046783569 |
| PLCB2 | 124.2206105 | 6.956760753 | 0.038206102 |
| TRPM7 | 46.53066447 | 5.540109884 | 0.021619661 |
| DTWD1 | 150.1904662 | 7.230649427 | 0.008352485 |
| TJP1 | 160.7694886 | 7.328849822 | 0.024036428 |
| TLE3 | 42.65947986 | 5.41479447 | 0.028410684 |
| ATXN2L | 102.4407153 | 6.678645422 | 0.01972478 |
| DYNC1LI2 | 86.32623311 | 6.431727132 | 0.006869309 |
| GALNS | 42.03172814 | 5.393406869 | 0.039725505 |
| SYNRG | 269.6490286 | 8.074939026 | 0.03115384 |
| OSBPL7 | 43.80763298 | 5.45311036 | 0.038968935 |
| ARSG | 229.4221903 | 7.841861129 | 0.021093134 |
| ABR | 126.384976 | 6.981681163 | 0.020389028 |
| MINK1 | 34.10411928 | 5.091874101 | 0.03084935 |
| RHOT1 | 53.59813878 | 5.744110998 | 0.020198713 |
| DHX33 | 137.9282572 | 7.10777424 | 0.006604968 |
| CDK5RAP3 | 28.51819743 | 4.83381089 | 0.042942346 |
| UNK | 83.26246265 | 6.379594324 | 0.02511573 |
| OSBPL1A | 67.11590927 | 6.068582881 | 0.030314842 |
| DCC | 264.5735912 | 8.047525254 | 0.002742717 |
| DTNA | 168.0594716 | 7.392828043 | 0.006934009 |
| ATP9B | 382.8788907 | 8.580744312 | 0.00431954 |
| MBP | 38.6972271 | 5.274158287 | 0.025969008 |
| NPC1 | 102.4012653 | 6.678089732 | 0.007055221 |
| MIB1 | 60.22329301 | 5.912249692 | 0.029395061 |
| PTPRM | 322.4629976 | 8.332989812 | 0.00183749 |
| CEP76 | 177.8337503 | 7.474385343 | 0.048906755 |
| PIAS2 | 110.7831252 | 6.791594332 | 0.022181172 |
| SEC11C | 817.6944051 | 9.675417959 | 0.015323162 |
| ZNF234 | 250.6304462 | 7.969417871 | 0.003409815 |
| ZNF350 | 69.40196237 | 6.116904551 | 0.037780245 |
| ZNF708 | 263.2663612 | 8.040379383 | 0.002114373 |
| CEP89 | 83.53207839 | 6.38425843 | 0.014353684 |
| STK4 | 192.7703499 | 7.590739357 | 0.003725843 |
| NCF4 | 151.4080473 | 7.242298076 | 0.003713208 |
| NF2 | 228.0649087 | 7.833300673 | 0.040879471 |
| ASCC2 | 0.005695223 | -7.456032034 | 0.041003969 |
| THOC5 | 71.47857554 | 6.159438979 | 0.0483549 |
| HUWE1 | 31.72860687 | 4.987712272 | 0.031361722 |
| CHM | 53.54905523 | 5.742789217 | 0.012784258 |
| BRWD3 | 21.17921232 | 4.40457703 | 0.049673973 |
| CXorf40B | 899.6704652 | 9.813252852 | 0.00123812 |
| MOSPD1 | 109.7309754 | 6.777827024 | 0.032058786 |
| ZNF41 | 27.88950308 | 4.801650324 | 0.044564838 |
| BRCC3 | 156.7813637 | 7.292610269 | 0.010407954 |
| PAM | 263.1938475 | 8.039981954 | 0.009260889 |
| NSD1 | 22.65754665 | 4.50191975 | 0.047816529 |
| PHYKPL | 0.002180316 | -8.84124726 | 0.049343943 |
| ZFYVE16 | 44.2287388 | 5.466912199 | 0.016326514 |
| DOP1A | 64.22495067 | 6.005061972 | 0.02870185 |
| GABBR1 | 30.35432932 | 4.923830392 | 0.045913744 |
| AHI1 | 73.19647947 | 6.193702356 | 0.013232033 |
| RNASET2 | 335.4688667 | 8.390035073 | 0.002746514 |
| NUB1 | 47.46716413 | 5.568857955 | 0.022738079 |
| PTK2 | 45.13094972 | 5.496045233 | 0.041969787 |
| ST6GALNAC4 | 123.6071202 | 6.949618039 | 0.043762504 |
| SNX30 | 300.1093405 | 8.229344412 | 0.029684419 |
| CC2D2B | 261.8805904 | 8.032765326 | 0.043238063 |
| DCAF6 | 20.26767946 | 4.341109012 | 0.043965931 |
| GTF2H1 | 72.73799349 | 6.184637224 | 0.009912766 |
| PPTC7 | 45.51313585 | 5.508211086 | 0.020101946 |
| STK38L | 85.69735584 | 6.421178786 | 0.009532739 |
| TSPAN31 | 107.1035399 | 6.742862354 | 0.008122179 |
| SPATS2 | 209.3710754 | 7.709918337 | 0.00607819 |
| ANKRD10 | 0.007857297 | -6.991751218 | 0.045000467 |
| TDRD3 | 22.30595961 | 4.479357309 | 0.031146955 |
| MARK3 | 32.88424497 | 5.039324641 | 0.038741574 |
| TDP1 | 87.49181654 | 6.451076177 | 0.045914779 |
| WDR78 | 57.49375083 | 5.845333249 | 0.029124515 |
| CEMIP | 173.6341658 | 7.439907043 | 0.003228389 |
| CDC7 | 56.653743 | 5.82409937 | 0.028682894 |
| HMCN1 | 3552.939804 | 11.79479753 | 0.001512212 |
| CDH11 | 52.31572938 | 5.709172871 | 0.029804964 |
| FTO | 366.4771574 | 8.517579467 | 0.002562877 |
| RSPRY1 | 102.5506429 | 6.680192726 | 0.046541938 |
| TMC8 | 36.32925226 | 5.183059767 | 0.03505328 |
| GPATCH8 | 0.006058413 | -7.366844435 | 0.040666806 |
| AMZ2 | 31.38820157 | 4.972150465 | 0.03494239 |
| BCAS3 | 153.2885195 | 7.260105841 | 0.00623946 |
| SRSF11 | 137.6587099 | 7.104952084 | 0.008228826 |
| PTPN2 | 68.39161727 | 6.0957476 | 0.035141395 |
| ZBTB14 | 95.64701266 | 6.579648004 | 0.021219526 |
| ZNF558 | 50.01714198 | 5.644350718 | 0.027823292 |
| ZHX3 | 0.006294145 | -7.311773787 | 0.048047276 |
| USP25 | 122.7380868 | 6.93943919 | 0.034584126 |
| TMEM184B | 76.5368182 | 6.258082021 | 0.039702711 |
| FAM118A | 222.4008224 | 7.797018313 | 0.048923417 |
| OGT | 126.424749 | 6.982135104 | 0.015973116 |
| LOC102724993 | 75.43207748 | 6.237106255 | 0.010975219 |
| TET3 | 121.8719556 | 6.92922237 | 0.040826011 |
| EFEMP1 | 73.31193775 | 6.195976234 | 0.038490285 |
| PRKD3 | 717.0878337 | 9.48600603 | 0.014580457 |
| MSH6 | 93.49985187 | 6.546892174 | 0.012233214 |
| HDLBP | 23.02581642 | 4.525180405 | 0.042715136 |
| ITSN2 | 139.3971614 | 7.123057373 | 0.010303347 |
| KDM3A | 389.7838919 | 8.606530661 | 0.037866422 |
| UGP2 | 97.45597396 | 6.606678719 | 0.004246567 |
| FAM49A | 77.61562212 | 6.278275156 | 0.020379166 |
| CCNT2 | 0.005113162 | -7.611568675 | 0.035704645 |
| ANKRD44 | 164.2203629 | 7.359489218 | 0.028540125 |
| EPHB1 | 347.6628005 | 8.441544898 | 0.009114548 |
| VPS8 | 31.39211066 | 4.972330127 | 0.041912092 |
| MYLK | 50.21864057 | 5.65015107 | 0.026164669 |
| PLOD2 | 62.27440893 | 5.960567518 | 0.013951206 |
| FCGR2B | 479.8541103 | 8.906452041 | 0.009045526 |
| CEP44 | 73.23720403 | 6.194504809 | 0.04884125 |
| ZNF518B | 53.1400575 | 5.731727884 | 0.020373261 |
| LHFPL2 | 32.33117106 | 5.014853859 | 0.022763807 |
