## Supplemental Data 3 for "Integrated analysis of high-throughput sequencing-based lncRNA-mediated ceRNA network in Hepatic Alveolar Echinococcosis"

Supplementary Table 3 The degree of ceRNA network

| degree.layout | Gene Symbol | type |
| --- | --- | --- |
| 27 | hsa-miR-17-5p | miRNA |
| 25 | hsa-miR-20b-5p | miRNA |
| 16 | lnc-PCBP1-AS1 | LncRNA |
| 13 | lnc-ANKRD36B | LncRNA |
| 12 | lnc-HNRNPU | LncRNA |
| 12 | lnc-PRIM2 | LncRNA |
| 11 | hsa-miR-107 | miRNA |
| 10 | hsa-miR-24-3p | miRNA |
| 10 | hsa-miR-212-3p | miRNA |
| 10 | lnc-H19 | LncRNA |
| 9 | hsa-miR-129-5p | miRNA |
| 9 | hsa-miR-27a-3p | miRNA |
| 9 | lnc-WDFY3-AS2 | LncRNA |
| 8 | hsa-miR-23b-3p | miRNA |
| 8 | hsa-miR-125b-5p | miRNA |
| 8 | hsa-miR-4465 | miRNA |
| 8 | lnc-PPP1R9B | LncRNA |
| 7 | hsa-miR-363-3p | miRNA |
| 7 | hsa-miR-338-3p | miRNA |
| 7 | lnc-SNHG12 | LncRNA |
| 6 | hsa-miR-4262 | miRNA |
| 6 | lnc-SNHG5 | LncRNA |
| 6 | hsa-miR-193a-3p | miRNA |
| 6 | hsa-miR-125a-5p | miRNA |
| 6 | lnc-SNHG11 | LncRNA |
| 5 | hsa-miR-22-3p | miRNA |
| 5 | hsa-miR-429 | miRNA |
| 4 | hsa-miR-3666 | miRNA |
| 4 | hsa-miR-4295 | miRNA |
| 4 | hsa-miR-4319 | miRNA |
| 4 | hsa-miR-146b-5p | miRNA |
| 3 | CAPRIN2 | mRNA |
| 3 | ZFYVE26 | mRNA |
| 3 | UBR3 | mRNA |
| 2 | BRWD1 | mRNA |
| 2 | UNK | mRNA |
| 2 | CLIP4 | mRNA |
| 2 | OXR1 | mRNA |
| 2 | FMNL3 | mRNA |
| 2 | TET3 | mRNA |
| 2 | PLAG1 | mRNA |
| 2 | PAFAH1B1 | mRNA |
| 2 | NRIP3 | mRNA |
| 2 | USP28 | mRNA |
| 2 | ENPP5 | mRNA |
| 2 | DPYSL2 | mRNA |
| 2 | SAMD8 | mRNA |
| 2 | XIAP | mRNA |
| 2 | TCF4 | mRNA |
| 2 | TTPAL | mRNA |
| 2 | LDLR | mRNA |
| 2 | PRDM1 | mRNA |
| 2 | ZNF385A | mRNA |
| 2 | DHX33 | mRNA |
| 2 | DICER1 | mRNA |
| 1 | RRAGD | mRNA |
| 1 | MECOM | mRNA |
| 1 | BRWD3 | mRNA |
| 1 | ZEB1 | mRNA |
| 1 | PHF21A | mRNA |
| 1 | ZNF462 | mRNA |
| 1 | HMGB1 | mRNA |
| 1 | EBF1 | mRNA |
| 1 | lnc-MIR17HG | LncRNA |
| 1 | HK2 | mRNA |
| 1 | VDR | mRNA |
| 1 | ZNF410 | mRNA |
| 1 | REV3L | mRNA |
| 1 | GOLGA8A | mRNA |
| 1 | KIF5B | mRNA |
| 1 | LHFPL2 | mRNA |
| 1 | NRP1 | mRNA |
| 1 | ALG9 | mRNA |
| 1 | SMAD5 | mRNA |
| 1 | YAP1 | mRNA |
| 1 | PTPRF | mRNA |
| 1 | SETD5 | mRNA |
| 1 | DYNC1LI2 | mRNA |
| 1 | EMSY | mRNA |
| 1 | MSANTD2 | mRNA |
| 1 | YWHAZ | mRNA |
| 1 | CEP170 | mRNA |
| 1 | TNKS2 | mRNA |
| 1 | EGLN3 | mRNA |
| 1 | PPP1R11 | mRNA |
| 1 | TJP1 | mRNA |
| 1 | CSNK1G3 | mRNA |
| 1 | OGT | mRNA |
