## Supplemental Data 4 for "Integrated analysis of high-throughput sequencing-based lncRNA-mediated ceRNA network in Hepatic Alveolar Echinococcosis"

Supplementary Table 4 Top 10 GO and KEGG enrichment for DEmRNAs in ceRNA network

| Category | ID | Description | Count | P value |
| --- | --- | --- | --- | --- |
| Biological Process | GO:0090263 | positive regulation of canonical Wnt signaling pathway | 5 | 6.27113E-05 |
|  | GO:0051642 | centrosome localization | 3 | 8.0556E-05 |
|  | GO:0061842 | microtubule organizing center localization | 3 | 8.90102E-05 |
|  | GO:0030177 | positive regulation of Wnt signaling pathway | 5 | 0.000160002 |
|  | GO:0034968 | histone lysine methylation | 4 | 0.000314937 |
|  | GO:0050769 | positive regulation of neurogenesis | 7 | 0.00039385 |
|  | GO:0018023 | peptidyl-lysine trimethylation | 3 | 0.000444081 |
|  | GO:0022604 | regulation of cell morphogenesis | 7 | 0.000467014 |
|  | GO:0018022 | peptidyl-lysine methylation | 4 | 0.000514145 |
|  | GO:0016571 | histone methylation | 4 | 0.000694707 |
| Cellular Component | GO:0005874 | microtubule | 7 | 0.000174111 |
|  | GO:0090575 | RNA polymerase II transcription regulator complex | 4 | 0.001015456 |
|  | GO:0030426 | growth cone | 4 | 0.001661076 |
|  | GO:0030427 | site of polarized growth | 4 | 0.001903736 |
|  | GO:0005667 | transcription regulator complex | 5 | 0.005544919 |
|  | GO:0005871 | kinesin complex | 2 | 0.008748263 |
|  | GO:0005875 | microtubule associated complex | 3 | 0.009656627 |
|  | GO:0150034 | distal axon | 4 | 0.010397188 |
|  | GO:1904115 | axon cytoplasm | 2 | 0.010835924 |
|  | GO:0005876 | spindle microtubule | 2 | 0.014335153 |
| Molecular Function | GO:0046974 | histone methyltransferase activity (H3-K9 specific) | 2 | 0.000362835 |
|  | GO:0051010 | microtubule plus-end binding | 2 | 0.001503888 |
|  | GO:0008017 | microtubule binding | 4 | 0.007148878 |
|  | GO:0018024 | histone-lysine N-methyltransferase activity | 2 | 0.007163626 |
|  | GO:0016706 | 2-oxoglutarate-dependent dioxygenase activity | 2 | 0.007808804 |
|  | GO:0070888 | E-box binding | 2 | 0.009174597 |
|  | GO:0042054 | histone methyltransferase activity | 2 | 0.012604493 |
|  | GO:0008201 | heparin binding | 3 | 0.01283321 |
|  | GO:0016279 | protein-lysine N-methyltransferase activity | 2 | 0.013431783 |
|  | GO:0016278 | lysine N-methyltransferase activity | 2 | 0.013854088 |
| KEGG pathways | hsa04520 | Adherens junction | 2 | 0.019400134 |
|  | hsa01524 | Platinum drug resistance | 2 | 0.020440846 |
|  | hsa04931 | Insulin resistance | 2 | 0.042135565 |
|  | hsa04066 | HIF-1 signaling pathway | 2 | 0.042844373 |
|  | hsa05145 | Toxoplasmosis | 2 | 0.044997883 |
